# Mapping the human genetic architecture of COVID-19: an update

**DOI:** 10.1101/2021.11.08.21265944

**Authors:** COVID-19 Host Genetics Initiative, Andrea Ganna

**Affiliations:** Massachusetts general hospital; Broad Institute

## Abstract

The Coronavirus Disease 2019 (COVID-19) pandemic continues to pose a major public health threat especially in countries with low vaccination rates. To better understand the biological underpinnings of SARS-CoV-2 infection and COVID-19 severity we formed the COVID19 Host Genetics Initiative. Here we present GWAS meta-analysis of up to 125,584 cases and over 2.5 million controls across 60 studies from 25 countries, adding 11 new genome-wide significant loci compared to those previously identified. Genes in novel loci include *SFTPD, MUC5B* and *ACE2*, reveal compelling insights regarding disease susceptibility and severity.

## Main text

The Coronavirus Disease 2019 (COVID-19) pandemic continues to pose a major public health threat especially in countries with low vaccination rates. To better understand the biological underpinnings of SARS-CoV-2 infection and COVID-19 severity we formed the COVID19 Host Genetics Initiative. Here we present GWAS meta-analysis of up to 125,584 cases and over 2.5 million controls across 60 studies from 25 countries, adding 11 new genome-wide significant loci compared to those previously identified^1^. Genes in novel loci include *SFTPD, MUC5B* and *ACE2*, reveal compelling insights regarding disease susceptibility and severity.

### Additional genomic regions identified for COVID-19 severity and SARS-CoV-2 infection

Here we present meta-analyses bringing together 60 studies from 25 countries (**Figure 1; Supplementary Table 1**) for three COVID-19 related phenotypes: (1) individuals critically ill with COVID-19 based on either requiring respiratory support in hospital or who died as a consequence of the disease (9,376 cases - of which 3,197 new in this data release – and 1,776,645 controls), (2) cases with moderate or severe COVID-19 defined as those hospitalized due to symptoms associated with the infection (25,027 cases – 11,386 new - and 2,836,272 controls), and (3) all cases with reported SARS-CoV-2 infection regardless of symptoms (125,584 cases – 76,022 new - and 2,575,347 controls). An overview of the study design is provided in **Supplementary Figure 1**. We found a total of 23 genome-wide significant loci (P < 5 × 10^−8^) of which 20 loci remain significant after multiple testing corrections (P < 1.67 × 10^−8^) to account for the number of phenotypes examined (**Figure 2; Supplementary Figure 2; Supplementary Table 2**). We compared the effects of these loci between the previous1 and the current analysis and found that only one locus did not replicate (rs72711165). All the other loci showed the expected increase in statistical significance (Supplementary Figure 3).

**Figure 1.**
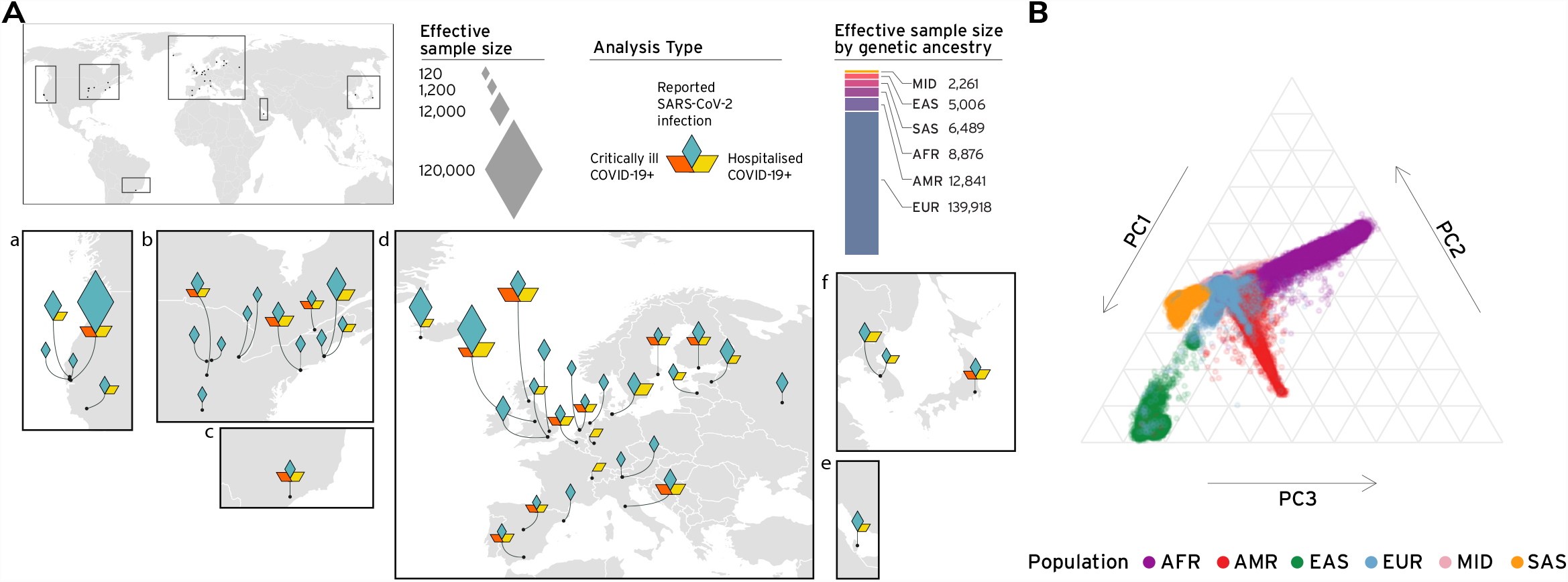
**Panel A: Geographical overview** of the contributing studies to the COVID-19 HGI and composition by major ancestry groups. Populations are defined as Middle Eastern (MID), South Asian (SAS), East Asian (EAS), African (AFR), Admixed American (AMR), European (EUR). **Panel B. Principal components analysis** highlights the population structure and the sample ancestry of the individuals participating to the COVID-19 HGI. This figure is an updated version of the figure published by the COVID-19 Host Genetics Initiative ^1^.

**Figure 2.**
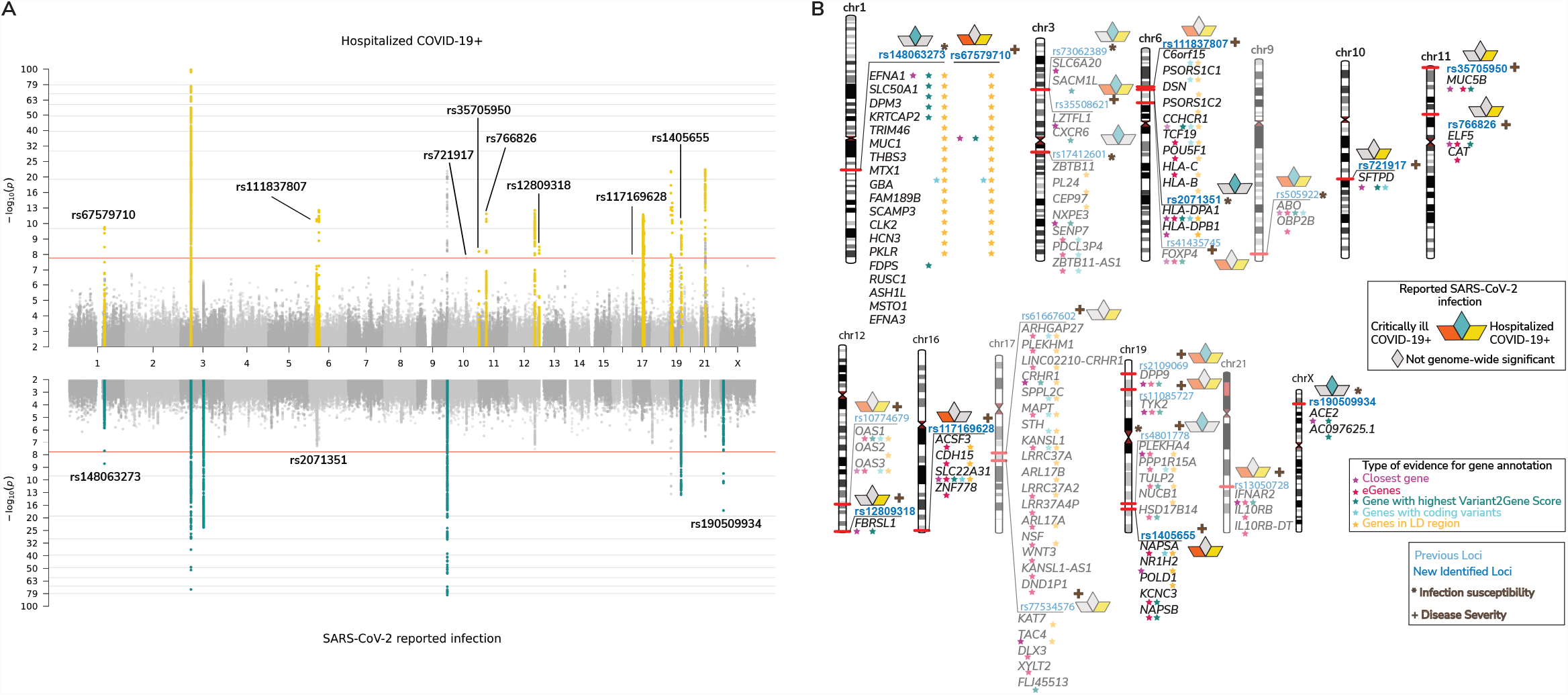
Genome-wide association results for COVID-19. A. Top panel shows results of genome-wide association study of hospitalized COVID-19 (n=25,027 cases and n=2,836,272 controls), and bottom panel the results of reported SARS-CoV-2 infection (n=125,584 cases and n=2,575,347 controls). Loci highlighted in yellow (top panel) represent regions associated with severity of COVID-19 manifestation. Loci highlighted in green (bottom panel) are regions associated with susceptibility to SARS-CoV-2 infection. Lead variants for the loci newly identified in this data release are annotated with their respective rsID. **B**. Results of gene prioritization using different evidence measures of gene annotation. Genes in linkage disequilibrium (LD) region, genes with coding variants and eGenes (fine-mapped cis-eQTL variant PIP > 0.1 in GTEx Lung) are annotated if in LD with a COVID-19 lead variant (r2 > 0.6). V2G: Highest gene prioritized by OpenTargetGenetics’ V2G score. The * indicates SARS-CoV-2 infection susceptibility and + indicates COVID-19 severity. The transparent loci were reported in the previous freeze (data release 5), and loci in bright blue were identified in the current freeze (data release 6). This figure is an updated version of the figure published by the COVID-19 Host Genetics Initiative ^1^.

### Heterogeneity in genetic effects across phenotypes, studies, and ancestry groups

Across the genome-wide significant loci, we observed clear patterns of association to the different phenotypes under study. Thus, we developed a two-class Bayesian model for classifying loci based on the patterns of association across the two better-powered phenotypes (COVID-19 hospitalization and SARS-CoV-2 reported infection). Intuitively, loci that are associated to susceptibility will also be associated to severity as to develop COVID-19, SARS-CoV-2 infection needs to first occur. In contrast, those genetic effects that solely modify the course of illness should be associated to severity to illness and not show any association to reported infection except via preferential ascertainment of hospitalized cases in a cohort (**Supplementary Methods**). We identified 16 loci that are substantially more likely (> 99% posterior probability) to impact the risk of COVID-19 hospitalization and 7 loci that clearly influence susceptibility to SARS-CoV-2 infection (**Supplementary Table 3; Supplementary Figure 4**).

We observed that several loci had a significant heterogeneous effect across studies (6/23 loci with P-value for heterogeneity < 2.2×10^−3^; **Supplementary Table 2**). Thanks to an increased diversity in our study population (**Supplementary Figure 5**) we were able to explore if such heterogeneity was due to effect differences across continental ancestry groups. Only one locus (FOXP4) showed a significantly different effect across ancestries (P-value heterogeneity < 7×10^−5^; **Supplementary Table 4 and Supplementary Figure 6**), though even at this locus all ancestry groups showed a positive effect estimate. This confirms that factors related to between-study heterogeneity (e.g., variable definition of COVID-19 severity due to different thresholds for testing, hospitalization, and patient recruitment) rather than differences across ancestries are a more likely explanation for the observed heterogeneity in the effect sizes across studies.

### Biological insights from novel loci

For the 23 genome-wide significant loci, we explored candidate causal genes and performed a phenome-wide association study to better understand their potential biological mechanisms (**Supplementary Table 2**,**5**,**6**; **Supplementary Figure 7**). Several of these loci with prior and direct connections to lung disease and SARS-CoV-2 infection mechanism are highlighted here.

Several loci involved in COVID-19 severity implicate lung surfactant biology. A missense variant rs721917:A>G (p.Met31Thr) in *SFTPD* (10q22.3) confers risk for hospitalization (OR [95% confidence interval [CI] = 1.06 [1.04, 1.08]; *P* = 1.7 × 10^−8^) and has been previously associated with increased risk of chronic obstructive pulmonary disease^2^ (OR = 1.08; *P* = 2.0 × 10^−8^), and decreased lung function^3^ (FEV1/FVC; β = –0.019; *P* = 2.0 × 10^−15^). *SFTPD* encodes the surfactant protein D (SP-D) that participates in innate immune response, protecting the lungs against inhaled microorganisms. The recombinant fragment of SP-D binds to the S1 spike protein of the SARS-CoV-2 and potentially inhibits binding to ACE2 receptor and SARS-CoV-2 infection^4^. Another missense variant rs117169628:G>A (p.Pro256Leu) in *SLC22A31* (16q24.3) also conferring risk for hospitalization (OR [95% CI] = 1.09 [1.06, 1.13]; *P* = 2.6 × 10^−8^). *SLC22A31* belongs to the family of solute carrier proteins that facilitate transport across membranes^5^ and is co-regulated with other surfactant proteins^6^.

We found a variant rs35705950:G>T located in the promoter of *MUC5B* (11p15.5) to be protective against hospitalization (OR [95% CI] = 0.83 [0.86, 0.93]; *P* = 6.5 × 10^−9^). This well-studied promoter variant increases expression of *MUC5B* in lung in GTEx (*P* = 6.7 × 10^−16^) and is the strongest known variant associated with *increased* risk of developing idiopathic pulmonary fibrosis (IPF)^7,8^, but also improves survival in IPF patients carrying this mutation^9^.

Finally, we identified rs190509934:T>C, located 69 bp upstream of *ACE2* (Xp22.2) to be associated with decreased susceptibility risk (OR [95% CI] = 0.69 [0.63, 0.75]; *P* = 3.6 × 10^−18^). ACE2 is the SARS-CoV-2 receptor and functionally interacts with *SLC6A19* and *SLC6A20* ^10^, one of which also showed a significant association with susceptibility (rs73062389:G>A at *SLC6A20*; OR [95% CI] = 1.18 [1.16, 1.20]; *P* = 2.5 × 10^−74^). Notably, rs190509934 is 10 times more common in South Asians (MAF = 0.027) than in Europeans (MAF = 0.0024), demonstrating the importance of diversity for variant discovery. Recent results have shown that rs190509934:T>C variant lowers *ACE2* expression, which in turn confers protection from SARS-CoV-2 infection^11^.

We applied Mendelian randomization to infer potential causal relationships between COVID-19 related phenotypes and their genetically correlated traits (**Supplementary Methods; Supplementary Tables 7**,**8**,**9; Supplementary Figure 8**). A novel causal association was observed between genetic liability to type II diabetes (T2D) and SARS-CoV-2 reported infection (OR [95% CI] = 1.02 [1.01, 1.03]; P = 1.6 × 10-3), and COVID-19 hospitalization (OR [95% CI] = 1.06 [1.03, 1.1]; P = 1.4 × 10-4). Multivariable MR (MVMR) was used to estimate the direct effect of liability to T2D on COVID-19-related phenotypes that was not mediated via BMI. This analysis indicated that the observed causal association of liability to T2D on COVID-19 phenotypes is mediated by BMI (**Supplementary Table 10**).

## Discussion

We have substantially expanded the genetic analysis of SARS-CoV-2 infection and COVID-19 severity by doubling the case size, identifying 11 novel loci. We developed a new approach to systematically assign the 23 discovered loci to either disease susceptibility (7 loci) or disease severity (16 loci). While distinguishing the two phenotypes is challenging because progression to a severe form of the disease requires susceptibility to infection in the first place, it is now evident that the genetic mechanisms involved in these two aspects of the disease can be differentiated. Among the new loci associated with disease susceptibility, *ACE2* represents an expected, yet interesting finding. *MUC5B, SFTPD*, and *SLC22A31* are the three most interesting novel loci associated with COVID-19 severity. Their relationship with lung function and lung diseases is consistent with loci previously associated with disease severity. The SPs secreted by alveolar cells, representing an emerging biological mechanism, maintains healthy lung function and facilitates the clearance of pathogens^12^. The protective effect of the *MUC5B* variant is unexpected given the otherwise risk-increasing, concordant effect between IPF and COVID-19 observed for other variants^8^. Nonetheless, this result aligns with the *MUC5B* promoter variant association that shows a twofold higher survival rate among IPF patients ^9^. In mice, *Muc5b* appears essential for effective mucociliary clearance and for controlling infection^13^ supporting therapies to control mucin secretion to be of potential benefit in COVID-19 patients.

Expanding genomic research to include participants from around the world enabled us to test if the effect of COVID-19related genetic variants was markedly different across ancestry groups. We didn’t detect obvious heterogeneity between ancestry groups, and we attribute the observed heterogeneity in the effect of COVID-19 related genetic variants to the diverse inclusion criteria across studies in terms of COVID-19 severity. However, we also note that ascertainment differences across studies might mask true underlying differences in effect sizes between ancestry groups.

The novel biological insights gained by this expansion of the COVID-19 Host Genetic Initiative showed that increasing sample size and diversity remain a fruitful activity to better understand the human genetic architecture of COVID-19.

## Data Availability

All data produced are available online at https://www.covid19hg.org

## Data Availability

Summary statistics generated by COVID-19 HGI are available at https://www.covid19hg.org/results/r6/. The analyses described here utilize the freeze 6 data. COVID-19 HGI continues to regularly release new data freezes. Summary statistics for non-European ancestry samples are not currently available due to the small individual sample sizes of these groups, but results for 23 loci lead variants are reported in Supplementary Table 3. Individual level data can be requested directly from contributing studies, listed in Supplementary Table 1. We used publicly available data from GTEx (https://gtexportal.org/home/), the Neale lab (http://www.nealelab.is/uk-biobank/), Finucane lab (https://www.finucanelab.org), FinnGen Freeze 4 cohort (https://www.finngen.fi/en/access_results), and eQTL catalogue release 3 (http://www.ebi.ac.uk/eqtl/).

## Code Availability

The code for summary statistics lift-over, the projection PCA pipeline including precomputed loadings and meta-analyses are available on GitHub (https://github.com/covid19-hg/) and the code for the Mendelian randomization and genetic correlation pipeline is available on GitHub at https://github.com/marcoralab/MRcovid. Code for implementing the MVMR analysis is available at: https://github.com/marcoralab/multivariate_MR; and subtype analyses at: https://github.com/mjpirinen/covid19-hgi_subtypes

## Author contributions

Detailed author contributions are integrated in the authorship list.

**COVID-19 Host Genetics Initiative**

**Leadership**

**Leadership**

Gita A. Pathak^1^,

Juha Karjalainen^2^,

Christine Stevens^3^,

Benjamin M Neale^4^,

Mark Daly^2,3,5^,

Andrea Ganna^2,3,5^

**Writing group**

**Writing group lead**

Gita A. Pathak^1^,

Shea J. Andrews^6^,

Masahiro Kanai^3^,

Mattia Cordioli^7^,

Andrea Ganna^2,3,5^

**Analysis group**

**Manuscript analyses team lead**

Juha Karjalainen^2^

**Manuscript analyses team members: phewas**

Gita A. Pathak^1^,

Renato Polimanti^1^

**Manuscript analyses team members: mendelian randomization**

Shea J. Andrews^6^,

Nadia Harerimana^8^

**Manuscript analyses team members: methods development**

Mattia Cordioli^7^,

Matti Pirinen^7^

**Manuscript analyses team members: PC projection, gene prioritization**

Masahiro Kanai^3^

**Project management group**

**Project management lead**

Christine Stevens^3^,

Rachel G. Liao^3^

**Project managment support**

Karolina Chwialkowska^9^,

Amy Trankiem^3^,

Mary K. Balaconis^3^

**Website development**

**Website development**

Huy Nguyen^3^,

Matthew Solomonson^3^

**Scientific communication group**

**Scientific communication lead**

Kumar Veerapen^3^,

Brooke Wolford^10^

**AncestryDNA COVID-19 Research Study**

**Analysis Team Lead**

Genevieve Roberts^11^

**Data Collection Lead**

Danny Park^11^

**Admin Team Lead**

Catherine A. Ball^11^

**Analysis Team Member**

Marie Coignet^11^,

Shannon McCurdy^11^,

Spencer Knight^11^,

Raghavendran Partha^11^,

Brooke Rhead^11^

**Data Collection Member**

Miao Zhang^11^,

Nathan Berkowitz^11^,

Michael Gaddis^11^,

Keith Noto^11^,

Luong Ruiz^11^,

Milos Pavlovic^11^

**Admin Team Member**

Eurie L. Hong^11^,

Kristin Rand^11^,

Ahna Girshick^11^,

Harendra Guturu^11^,

Asher Haug Baltzell^11^

**BelCovid**

**Analysis Team Lead**

Mari E.K. Niemi^12^

**Data Collection Lead**

Souad Rahmouni^13^,

Julien Guntz^14^

**Admin Team Lead**

Yves Beguin^15^

**Analysis Team Member**

Mattia Cordioli^16^,

Sara Pigazzini^12^,

Lindokuhle Nkambule^17,18,19^

**Data Collection Member**

Michel Georges^13^,

Michel Moutschen^20,21^,

Benoit Misset^20,21^,

Gilles Darcis^20,21^,

Julien Guiot^20,21^,

Samira Azarzar^20,21^,

Stéphanie Gofflot^15^,

Sabine Claassen^14^,

Olivier Malaise^20^,

Pascale Huynen^20^,

Christelle Meuris^22^,

Marie Thys^22^,

Jessica Jacques^22^,

Philippe Léonard^22^,

Frederic Frippiat^22^,

Jean-Baptiste Giot^22^,

Anne-Sophie Sauvage^22^,

Christian Von Frenckell^22^,

Yasmine Belhaj^13^,

Bernard Lambermont^22^

**Biobanque Quebec COVID19**

**Analysis Team Lead**

Tomoko Nakanishi^23,24,25,26^

**Data Collection Lead**

David R. Morrison^24^

**Admin Team Lead**

Vincent Mooser^27^,

J. Brent Richards^24,28,29^

**Analysis Team Member**

Guillaume Butler-Laporte^24,30^,

Vincenzo Forgetta^24^,

Rui Li^27^

**Data Collection Member**

Biswarup Ghosh^24^,

Laetitia Laurent^24^,

Alexandre Belisle^27^,

Danielle Henry^24^,

Tala Abdullah^24^,

Olumide Adeleye^24^,

Noor Mamlouk^24^,

Nofar Kimchi^24^,

Zaman Afrasiabi^24^,

Nardin Rezk^24^,

Branka Vulesevic^24^,

Meriem Bouab^24^,

Charlotte Guzman^24^,

Louis Petitjean^24^,

Chris Tselios^24^,

Xiaoqing Xue^24^,

Jonathan Afilalo^24^,

Marc Afilalo^31,32^,

Maureen Oliveira^33^,

Bluma Brenner^34^,

Nathalie Brassard^35^,

Madeleine Durand^36,37^

**Admin Team Member**

Erwin Schurr^38^,

Pierre Lepage^27^,

Jiannis Ragoussis^27^,

Daniel Auld^27^,

Michaël Chassé^37,39^,

Daniel E. Kaufmann^37,40^,

G. Mark Lathrop^27^,

Darin Adra^24^

**CCHC COVID-19 GAWS**

**Analysis Team Lead**

Caroline Hayward^41^,

Joseph T Glessner^42,43^,

Douglas M Shaw^44^

**Data Collection Lead**

Archie Campbell^45^,

Marcela Morris^46^

**Admin Team Lead**

Hakon Hakonarson^42,43^,

David J Porteous^47^,

Jennifer Below^44^

**Analysis Team Member**

Anne Richmond^41^,

Xiao Chang^42^,

Hannah Polikowski^44^,

Petty E Lauren^44^,

Hung-Hsin Chen^44^,

Zhu Wanying^44^

**Data Collection Member**

Chloe Fawns-Ritchie^48^

**Admin Team Member**

Kari North^49^,

Joseph B McCormick^46^

**CHOP_CAG**

**Data Collection Member**

Xiao Chang^50^,

Joseph R Glessner^51,52^,

Hakon Hakonarson^51,53,54^

**Colorado Center for Personalized Medicine (CCPM)**

**Analysis Team Lead**

Christopher R Gignoux^55^

**Data Collection Lead**

Stephen J Wicks^55^,

Kristy Crooks^55^

**Admin Team Lead**

Kathleen C Barnes^55^

**Analysis Team Member**

Michelle Daya^55^,

Jonathan Shortt^55^,

Nicholas Rafaels^55^,

Sameer Chavan^55^

**Coronagenes**

**Analysis Team Lead**

Paul RHJ Timmers^56,57^,

James F Wilson^56,57^,

Albert Tenesa^56,58^

**Admin Team Lead**

Shona M Kerr^56^

**Analysis Team Member**

Kenton D’Mellow^58^

**Egypt hgCOVID hub**

**Analysis Team Lead**

Mari E.K. Niemi^12^

**Data Collection Lead**

Doaa Shahin^59^,

Yasser M. El-Sherbiny^59,60^

**Admin Team Lead**

Kathrin Aprile von Hohenstaufen^61^,

Ali Sobh^62^,

Madonna M. Eltoukhy^63^

**Analysis Team Member**

Mattia Cordioli^7^,

Lindokuhle Nkambul^17,64^

**Data Collection Member**

Tamer A. Elhadidy^65^,

Mohamed S. Abd Elghafar^66^,

Jehan J. El-Jawhari^59,60^,

Attia A.S. Mohamed^63^,

Marwa H. Elnagdy^67^,

Amr Samir^68^,

Mahmoud Abdel-Aziz^69^,

Walid T. Khafaga^70^,

Walaa M. El-Lawaty^71^,

Mohamed S. Torky^71^,

Mohamed R. El-shanshory^72^

**Admin Team Member**

Amr M. Yassen^73^,

Mohamed A.F. Hegazy^68^,

Kamal Okasha^74^,

Mohammed A. Eid^75^,

Hanteera S. Moahmed^71^

**EraCORE**

**Analysis Team Lead**

Carolina Medina-Gomez^76^

**Data Collection Lead**

M. Arfan Ikram^77^

**Admin Team Lead**

Andre G Uitterlinden^76,77^

**Estonian Biobank**

**Analysis Team Lead**

Reedik Mägi^78^

**Data Collection Lead**

Lili Milani^78^

**Admin Team Lead**

Andres Metspalu^78^

**Analysis Team Member**

Triin Laisk^78^,

Kristi Läll^78^,

Maarja Lepamets^78^

**Data Collection Member**

Tõnu Esko^78^,

Ene Reimann^78^,

Paul Naaber^79^,

Edward Laane^80,81^,

Jaana Pesukova^80^,

Pärt Peterson^82^,

Kai Kisand^83^,

Jekaterina Tabri^84^,

Raili Allos^84^,

Kati Hensen^84^,

Joel Starkopf^85^,

Inge Ringmets^86^,

Anu Tamm^87^,

Anne Kallaste^87^

**Admin Team Member**

Helene Alavere^78^,

Kristjan Metsalu^78^,

Mairo Puusepp^78^

**EXCEED**

**Analysis Team Lead**

Chiara Batini^88^

**Data Collection Lead**

Martin D Tobin^88,89^

**Admin Team Lead**

Laura D Venn^88^

**Analysis Team Member**

Paul H Lee^88^,

Nick Shrine^88^,

Alexander T Williams^88^

**Data Collection Member**

Anna L Guyatt^88^,

Catherine John^88^,

Richard J Packer^88^,

Altaf Ali^88^,

Robert C Free^90^,

Xueyang Wang^88^,

Louise V Wain^88,89^,

Edward J Hollox^91^

**Admin Team Member**

Catherine E Bee^88^,

Emma L Adams^88^

**FinnGen**

**Admin Team Lead**

FinnGen^92^

**Analysis Team Member**

Samuli Ripatti^3,93,94,95^,

Sanni Ruotsalainen^93^

**Data Collection Member**

Kati Kristiansson^96^,

Sami Koskelainen^96^,

Markus Perola^96,97^,

Kati Donner^7^,

Katja Kivinen^7^,

Aarno Palotie^7^

**Admin Team Member**

Mari Kaunisto^7^

**Functional Host Genomics in Infectious Diseases (FHoGID)**

**Analysis Team Lead**

Carlo Rivolta^98,99^

**Data Collection Lead**

Pierre-Yves Bochud^100^,

Stéphanie Bibert^101^,

Noémie Boillat^100^,

Semira Gonseth Nussle^102^,

Werner Albrich^103^

**Analysis Team Member**

Mathieu Quinodoz^98,99^,

Dhryata Kamdar^98,99^

**Data Collection Member**

Noémie Suh^104^,

Dionysios Neofytos^105^,

Véronique Erard^106^,

Cathy Voide^107^,

FHoGID^108^,

RegCOVID^109^,

P-PredictUs^110^,

SeroCOVID^111^,

CRiPSI^112^

**GCAT. Genomes For Life**

**Analysis Team Lead**

Rafael de Cid^113^

**Data Collection Lead**

Anna Carreras^113^,

Victor Moreno^114^,

Manolis Kogevinas^115,116,117,118^

**Analysis Team Member**

Iván Galván-Femenía^113^,

Natalia Blay^113^,

Xavier Farré^113^,

Lauro Sumoy^113^

**Data Collection Member**

Beatriz Cortés^113^,

Josep Maria Mercader^119^,

Marta Guindo-Martinez^120^,

David Torrents^120^,

Judith Garcia-Aymerich^115,117,118^,

Gemma Castaño-Vinyals^115,116,117,118^,

Carlota Dobaño^115,118^

**GEN-COVID Multicenter Study**

**Analysis Team Lead**

Marco Gori^121,122^,

Mari E.K. Niemi^12^

**Data Collection Lead**

Alessandra Renieri^123,124,125^,

Francesca Mari^123,124,125^,

Mario Umberto Mondelli^126,127^,

Francesco Castelli^128^,

Massimo Vaghi^129^,

Stefano Rusconi^130,131^,

Francesca Montagnani^132,133^,

Elena Bargagli^134^,

Federico Franchi^135^,

Maria Antonietta Mazzei^136^,

Luca Cantarini^137^,

Danilo Tacconi^138^,

Marco Feri^139^,

Raffaele Scala^140^,

Genni Spargi^141^,

Cesira Nencioni^142^,

Maria Bandini^143^,

Gian Piero Caldarelli^144^,

Anna Canaccini^145^,

Agostino Ognibene^146^,

Antonella D’Arminio Monforte^147^,

Massimo Girardis^148^,

Andrea Antinori^149^,

Daniela Francisci^150,151^,

Elisabetta Schiaroli^150,151^,

Pier Giorgio Scotton^152^,

Sandro Panese^153^,

Renzo Scaggiante^154^,

Matteo Della Monica^155^,

Mario Capasso^156,157,158^,

Giuseppe Fiorentino^159^,

Marco Castori^160^,

Filippo Aucella^161^,

Antonio Di Biagio^162^,

Luca Masucci^163,164^,

Serafina Valente^165^,

Marco Mandalà^166^,

Patrizia Zucchi^167^,

Ferdinando Giannattasio^168^,

Domenico A. Coviello^169^,

Cristina Mussini^170^,

Luisa Tavecchia^171^,

Lia Crotti^172,173,174,175^,

Marco Rizzi^176^,

Maria Teresa La Rovere^177^,

Simona Sarzi-Braga^178^,

Maurizio Bussotti^179^,

Sabrina Ravaglia^180^,

Rosangela Artuso^181^,

Antonio Perrella^182^,

Davide Romani^183^,

Paola Bergomi^184^,

Emanuele Catena^184^,

Antonella Vincenti^185^,

Claudio Ferri^186^,

Davide Grassi^186^,

Gloria Pessina^187^,

Mario Tumbarello^132,188^,

Massimo Di Pietro^189^,

Ravaglia Sabrina^190^,

Sauro Luchi^191^

**Admin Team Lead**

Simone Furini^125^,

Simona Dei^192^

**Analysis Team Member**

Elisa Benetti^125^,

Nicola Picchiotti^121,193^,

Maurizio Sanarico^194^,

Stefano Ceri^195^,

Pietro Pinoli^195^,

Francesco Raimondi^196^,

Filippo Biscarini^197^,

Alessandra Stella^198^,

Kristina Zguro^199^,

Katia Capitani^132,200^,

Mattia Cordioli^16^,

Sara Pigazzini^12^,

Mattia Cordioli^16^,

Sara Pigazzini^12^,

Lindokuhle Nkambule^17,64^,

Marco Tanfoni^121^

**Data Collection Member**

Chiara Fallerini^123,125^,

Sergio Daga^123,125^,

Margherita Baldassarri^123,125^,

Francesca Fava^123,124,125^,

Elisa Frullanti^123,125^,

Floriana Valentino^123,125^,

Gabriella Doddato^123,125^,

Annarita Giliberti^123,125^,

Rossella Tita^201^,

Sara Amitrano^201^,

Mirella Bruttini^123,132,201^,

Susanna Croci^123,125^,

Ilaria Meloni^123,125^,

Maria Antonietta Mencarelli^201^,

Caterina Lo Rizzo^201^,

Anna Maria Pinto^201^,

Giada Beligni^123,125^,

Andrea Tommasi^202^,

Laura Di Sarno^123,125^,

Maria Palmieri^123,125^,

Miriam Lucia Carriero^123,125^,

Diana Alaverdian^123,125^,

Stefano Busani^148^,

Raffaele Bruno^126,127^,

Marco Vecchia^203^,

Mary Ann Belli^171^,

Stefania Mantovani^203^,

Serena Ludovisi^204^,

Eugenia Quiros-Roldan^128^,

Melania Degli Antoni^128^,

Isabella Zanella^205,206^,

Matteo Siano^131^,

Arianna Emiliozzi^149^,

Massimiliano Fabbiani^133^,

Barbara Rossetti^133^,

Laura Bergantini^134^,

Miriana D’Alessandro^134^,

Paolo Cameli^134^,

David Bennett^134^,

Federico Anedda^135^,

Simona Marcantonio^135^,

Sabino Scolletta^135^,

Susanna Guerrini^136^,

Edoardo Conticini^137^,

Bruno Frediani^137^,

Chiara Spertilli^138^,

Alice Donati^139^,

Luca Guidelli^140^,

Marta Corridi^141^,

Leonardo Croci^142^,

Paolo Piacentini^143^,

Elena Desanctis^143^,

Silvia Cappelli^143^,

Agnese Verzuri^145^,

Valentina Anemoli^145^,

Alessandro Pancrazzi^146^,

Maria Lorubbio^146^,

Federica Gaia Miraglia^147^,

Sophie Venturelli^148^,

Andrea Cossarizza^207^,

Alessandra Vergori^149^,

Arianna Gabrieli^131^,

Agostino Riva^130,131^,

Francesco Paciosi^151^,

Francesca Andretta^152^,

Francesca Gatti^154^,

Saverio Giuseppe Parisi^208^,

Stefano Baratti^208^,

Carmelo Piscopo^155^,

Roberta Russo^156,157^,

Immacolata Andolfo^156,157^,

Achille Iolascon^156,157^,

Massimo Carella^160^,

Giuseppe Merla^156,209^,

Gabriella Maria Squeo^209^,

Pamela Raggi^210^,

Carmen Marciano^210^,

Rita Perna^210^,

Matteo Bassetti^162,211^,

Maurizio Sanguinetti^163,164^,

Alessia Giorli^166^,

Lorenzo Salerni^166^,

Pierpaolo Parravicini^167^,

Elisabetta Menatti^212^,

Tullio Trotta^168^,

Gabriella Coiro^168^,

Fabio Lena^213^,

Enrico Martinelli^214^,

Sandro Mancarella^171^,

Chiara Gabbi^194^,

Franco Maggiolo^176^,

Diego Ripamonti^176^,

Tiziana Bachetti^215^,

Claudia Suardi^216^,

Gianfranco Parati^173,217^,

Giordano Bottà^218^,

Paolo Di Domenico^218^,

Ilaria Rancan^219^,

Francesco Bianchi^132,182^,

Riccardo Colombo^184^,

Chiara Barbieri^220^,

Donatella Acquilini^221^,

Elena Andreucci^181^,

Francesco Paciosi^151^,

Francesco Vladimiro Segala^222^,

Giusy Tiseo^220^,

Marco Falcone^220^,

Mirjam Lista^132,223^,

Monica Poscente^187^,

Oreste De Vivo^165^,

Paola Petrocelli^224^,

Alessandra Guarnaccia^163^,

Silvia Baroni^225^

**Generation Scotland**

**Analysis Team Lead**

Caroline Hayward^226^

**Data Collection Lead**

David J Porteous^227^

**Admin Team Lead**

Chloe Fawns-Ritchie^228^

**Analysis Team Member**

Anne Richmond^226^

**Data Collection Member**

Archie Campbell^227^

**Genes & Health**

**Analysis Team Lead**

David A van Heel^229^

**Data Collection Lead**

Karen A Hunt^229^

**Admin Team Lead**

Richard C Trembath^230^

**Analysis Team Member**

Qin Qin Huang^231^,

Hilary C Martin^231^

**Data Collection Member**

Dan Mason^232^,

Bhavi Trivedi^233^,

John Wright^232^

**Admin Team Member**

Sarah Finer^234^,

Genes & Health Research Team^235^,

Christopher J Griffiths^236^

**Genes for Good**

**Analysis Team Lead**

Albert V Smith^237^

**Data Collection Member**

Andrew P Boughton^237^,

Kevin W. Li^237^,

Jonathon LeFaive^237^,

Aubrey Annis^237^

**Genome-wide assessment of the gene variants associated with severe COVID-19 phenotype in Iran**

**Analysis Team Lead**

Mari E.K. Niemi^12^,

Ahmadreza Niavarani^238^

**Data Collection Lead**

Rasoul Aliannejad^239^

**Analysis Team Member**

Mattia Cordioli^7^,

Lindokuhle Nkambul^17,64^,

Bahareh Sharififard^238^

**Data Collection Member**

Ali Amirsavadkouhi^240^,

Zeinab Naderpour^239^,

Hengameh Ansari Tadi^241^,

Afshar Etemadi Aleagha^242^,

Saeideh Ahmadi^243^,

Seyed Behrooz Mohseni Moghaddam^244^,

Alireza Adamsara^245^,

Morteza Saeedi^246^,

Hamed Abdollahi^247^,

Abdolmajid Hosseini^248^

**Host genetic factors in COVID-19 patients in relation to disease susceptibility, disease severity and pharmacogenomics**

**Analysis Team Lead**

Pajaree Chariyavilaskul^249,250^

**Data Collection Lead**

Watsamon Jantarabenjakul^251,252^

**Admin Team Lead**

Nattiya Hirankarn^253,254^

**Analysis Team Member**

Monpat Chamnanphon^255,256^,

Thitima B. Suttichet^255^,

Vorasuk Shotelersuk^257,258^,

Monnat Pongpanich^259,260^,

Chureerat Phokaew^258,261,262^,

Wanna Chetruengchai^258,262^

**Data Collection Member**

Opass Putchareon^251,263^,

Pattama Torvorapanit^251,263^,

Thanyawee Puthanakit^252,264^,

Pintip Suchartlikitwong^264,265^

**Admin Team Member**

Voraphoj Nilaratanakul^266,267^,

Pimpayao Sodsai^253,254^

**HUNT**

**Analysis Team Lead**

Ben M Brumpton^268,269,270^

**Data Collection Lead**

Kristian Hveem^268,269^,

Cristen Willer^271,272,273^

**Analysis Team Member**

Brooke Wolford^271,272,273^,

Wei Zhou^274,275^

**Data Collection Member** Tormod Rogne^276,277,278^,

Erik Solligard^276,278^,

Bjørn Olav Åsvold^268,269,270^

**Lifelines**

**Analysis Team Lead**

Lude Franke^279^

**Data Collection Lead**

Marike Boezen^280^

**Analysis Team Member**

Patrick Deelen^281^,

Annique Claringbould^279^,

Esteban Lopera^279^,

Robert Warmerdam^279^,

Judith. M. Vonk^282^,

Irene van Blokland^279^

**Data Collection Member**

Pauline Lanting^283^,

Anil P. S. Ori^284,285^

**Mass General Brigham - Host Vulnerability to COVID-19**

**Analysis Team Lead**

Yen-Chen Anne Feng^286^,

Josep Mercader^287,288^

**Data Collection Lead**

Scott T Weiss^289^,

Elizabeth W. Karlson^290^,

Jordan W. Smoller^291^,

Shawn N Murphy^292^,

James B. Meigs^293^,

Ann E. Woolley^290^

**Admin Team Lead**

Robert C. Green^294^

**Data Collection Member**

Emma F Perez^295^

**Michigan Genomics Initiative (MGI)**

**Analysis Team Lead**

Brooke Wolford^273^

**Admin Team Lead**

Sebastian Zöllner^237^

**Analysis Team Member**

Jiongming Wang^237^,

Andrew Beck^237^

**Mount Sinai Health System COVID-19 Genomics Initiative**

**Analysis Team Lead**

Laura G. Sloofman^296,297,298^

**Data Collection Lead**

Steven Ascolillo^299^,

Robert P. Sebra^300,301^,

Brett L. Collins^302^,

Tess Levy^302^

**Admin Team Lead**

Joseph D. Buxbaum^302^,

Stuart C. Sealfon^298^

**Analysis Team Member**

Shea J. Andrews^6^,

Daniel M. Jordan^303,304^,

Ryan C. Thompson^305,306,307^,

Kyle Gettler^308^,

Kumardeep Chaudhary^304,309^,

Gillian M. Belbin^310^,

Michael Preuss^311,312^,

Clive Hoggart^298,313^,

Sam Choi^314^,

Slayton J. Underwood^298,315^

**Data Collection Member**

Irene Salib^300^,

Bari Britvan^302^,

Katherine Keller^302^,

Lara Tang^302^,

Michael Peruggia^302^,

Liam L. Hiester^302^,

Kristi Niblo^302^,

Alexandra Aksentijevich^302^,

Alexander Labkowsky^302^,

Avromie Karp^302^,

Menachem Zlatopolsky^302^,

Marissa Zyndorf^300^

**Admin Team Member**

Alexander W. Charney^316^,

Noam D. Beckmann^299^,

Eric E. Schadt^300,301^,

Noura S. Abul-Husn^310^,

Judy H. Cho^304,309^,

Yuval Itan^304,309^,

Eimear E. Kenny^310^,

Ruth J.F. Loos^311,312,317^,

Girish N. Nadkarni^311,318,319,320,321^,

Ron Do^304,309^,

Paul O’Reilly^314^,

Laura M. Huckins^322^

**MyCode Health Initiative**

**Analysis Team Lead**

Manuel A.R. Ferreira^323^,

Goncalo R. Abecasis^323^

**Data Collection Lead**

Joseph B. Leader^324^,

Michael N. Cantor^323^

**Admin Team Lead**

Anne E Justice^325^,

Dave J. Carey^326^

**Analysis Team Member**

Geetha Chittoor^325^,

Navya Shilpa Josyula^325^,

Jack A. Kosmicki^323^,

Julie E Horowitz^323^,

Aris Baras^323^

**Data Collection Member**

Matthew C. Gass^324^,

Ashish Yadav^323^

**Admin Team Member**

Tooraj Mirshahi^326^

**Netherlands Twin Register**

**Analysis Team Lead**

Jouke Jan Hottenga^327^

**Data Collection Lead**

Meike Bartels^327^

**Admin Team Lead**

Eco (E.J.C.) de geus^327^

**Analysis Team Member**

Michel (M.G.) Nivard^327^

**Penn Medicine Biobank**

**Analysis Team Lead**

Anurag Verma^328^,

Marylyn D. Ritchie^328^

**Admin Team Lead**

Daniel Rader^328^

**Analysis Team Member**

Binglan Li^329^,

Shefali S Verma^328^,

Anastasia Lucas^328^,

Yuki Bradford^328^

**Saudi Human Genome Program-COVID19 : Host Genomic markers predicting the severity and suitability to COVID-19 in highly consanguineous population**

**Analysis Team Lead**

Malak Abedalthagafi^330^,

Manal Alaamery^331^

**Data Collection Lead**

Abdulraheem Alshareef^332^,

Mona Sawaji^333^

**Admin Team Lead**

Salam Massadeh^331^,

Abdulaziz AlMalik^334^

**Analysis Team Member**

Saleh Alqahtani^335^,

Dona Baraka^336^,

Fawz Al Harthi^330^,

Ebtehal Alsolm^330^,

Leen Abu Safieh^330^,

Albandary M Alowayn^330^,

Fatimah Alqubaishi^330^,

Amal Al Mutairi^330^,

Serghei Mangul^337^

**Data Collection Member**

Mansour Almutairi^331^,

Nora Aljawini^338^,

Nour Albesher^338^,

Yaseen M Arabi^339^,

Ebrahim S Mahmoud^339^,

Amin K Khattab^340^,

Roaa T Halawani^340^,

Ziab Z Alahmadey^340^,

Jehad K Albakri^340^,

Walaa A Felemban^340^,

Bandar A Suliman^332^,

Rana Hasanato^336^,

Laila Al-Awdah^341^,

Jahad Alghamdi^342^,

Deema AlZahrani^343^,

Sameera AlJohani^344^,

Hani Al-Afghani^345^,

Nouf AlDhawi^343^,

Hadeel AlBardis^330^,

Sarah Alkwai^338^,

Moneera Alswailm^338^,

Faisal Almalki^343^,

Maha Albeladi^343^,

Iman Almohammed^338^,

Eman Barhoush^346^,

Anoud Albader^343^

**Admin Team Member**

Sara Alotaibi^330^,

Bader Alghamdi^347^,

Junghyun Jung^348^,

Mohammad S fawzy^330^

**Data collection Member**

May Alrashed^349^

**The genetic predisposition to severe COVID-19**

**Analysis Team Lead**

Mari E.K. Niemi^12^

**Data Collection Lead**

Hugo Zeberg^350,351^

**Analysis Team Member**

Mattia Cordioli^16^,

Sara Pigazzini^12^,

Lindo Nkambul^3,17,352^

**Data Collection Member**

Robert Frithiof^353^,

Michael Hultström^353,354^,

Miklos Lipcsey^353,355^,

Nicolas Tardif^356^,

Olav Rooyackers^356^,

Jonathan Grip^356^,

Tomislav Maricic^351^

**The Norwegian Mother, Father and Child Cohort Study**

**Analysis Team Lead**

øyvind Helgeland^357^

**Data Collection Lead**

Per Magnus^358^,

Lill-Iren S Trogstad^359^

**Analysis Team Member**

Yunsung Lee^358^

**Admin Team Member**

Jennifer R Harris^357^

**TwinsUK**

**Analysis Team Lead**

Massimo Mangino^360,361^

**Data Collection Lead**

Tim D Spector^360^

**Data Collection Member**

Duncan Emma^360^

**UK 100**,**000 Genomes Project (Genomics England)**

**Analysis Team Lead**

Loukas Moutsianas^362,363^

**Data Collection Lead**

Mark J Caulfield^362,363,364^,

Richard H Scott^362,365^

**Analysis Team Member**

Athanasios Kousathanas^366^,

Dorota Pasko^366^,

Susan Walker^366^,

Alex Stuckey^366^,

Christopher A Odhams^366^,

Daniel Rhodes^366^

**Data Collection Member**

Tom Fowler^366^,

Augusto Rendon^362,367^,

Georgia Chan^366^,

Prabhu Arumugam^366^

**UK Biobank**

**Analysis Team Lead**

Tomoko Nakanishi^368^,

Konrad J. Karczewski^5,19^,

Alicia R. Martin^5,19^,

Daniel J Wilson^369^,

Chris C A Spencer^370^

**Data Collection Lead**

Derrick W Crook^371^,

David H Wyllie^371,372^,

Anne Marie O’Connell^373^

**Admin Team Lead**

J. Brent Richards^24,28,29^

**Analysis Team Member**

Guillaume Butler-Laporte^24,30^,

Vincenzo Forgetta^24^,

Elizabeth G. Atkinson^5,19^,

Masahiro Kanai^5,19,374^,

Kristin Tsuo^5,19,375^,

Nikolas Baya^5,19^,

Patrick Turley^5,19^,

Rahul Gupta^5,19^,

Raymond K. Walters^5,19^,

Duncan S. Palmer^5,19^,

Gopal Sarma^5,19^,

Matthew Solomonson^5,19^,

Nathan Cheng^5,19^,

Wenhan Lu^5,19^,

Claire Churchhouse^5,19^,

Jacqueline I. Goldstein^5,19^,

Daniel King^5,19^,

Wei Zhou^5,19^,

Cotton Seed^5,19^,

Mark J. Daly^2,3,5^,

Benjamin M. Neale^5,19^,

Hilary Finucane^5,19^,

Sam Bryant^3^,

F. Kyle Satterstrom^5,19^,

Gavin Band^376^,

Sarah G Earle^369^,

Shang-Kuan Lin^369^,

Nicolas Arning^369^,

Nils Koelling^370^

**Data Collection Member**

Jacob Armstrong^369^,

Justine K Rudkin^369^

**Admin Team Member**

Shawneequa Callier^377^,

Sam Bryant^5,19^,

Caroline Cusick^19^

**UK Blood Donors Cohort**

**Analysis Team Lead**

Nicole Soranzo^378,379,380^,

Jing Hua Zhao^381^

**Data Collection Lead**

John Danesh^381,382,383,384,385^,

Emanuele Di Angelantonio^381,382,383,384^

**Analysis Team Member**

Adam S. Butterworth^381,382,383,384^

**VA Million Veteran Program (MVP)**

**Analysis Team Lead**

Yan V Sun^386,387^,

Jennifer E Huffman^388^

**Data Collection Lead**

Kelly Cho^389^

**Admin Team Lead**

Christopher J O’Donnell^388^,

Phil Tsao^390,391^,

J. Michael Gaziano^389^

**Analysis Team Member**

Gina Peloso^388,392^

**Data Collection Member**

Yuk-Lam Ho^389^

**Vanda COVID**

**Analysis Team Lead**

Sandra P Smieszek^393^

**Admin Team Lead**

Christos Polymeropoulos^393^,

Vasilios Polymeropoulos^393^,

Mihael H Polymeropoulos^393^

**Analysis Team Member**

Bartlomiej P Przychodzen^393^

**Variability in immune response genes and severity of SARS-CoV-2 infection (INMUNGEN-CoV2 study)**

**Analysis Team Lead**

Israel Fernandez-Cadenas^394^

**Data Collection Lead**

Anna M Planas^395,396^

**Analysis Team Member**

Jordi Perez-Tur^397,398,399^,

Laia Llucià-Carol^394,400^,

Natalia Cullell^394,401^,

Elena Muiño^394^,

Jara Cárcel-Márquez^394^,

Marta L DeDiego^402^,

Lara Lloret Iglesias^403^

**Data Collection Member**

Alex Soriano^404^,

Veronica Rico^405^,

Daiana Agüero^405^,

Josep L Bedini^405^,

Francisco Lozano^406^,

Carlos Domingo^405^,

Veronica Robles^405^,

Francisca Ruiz-Jaén^407^,

Leonardo Márquez^408^,

Juan Gomez^409^,

Eliecer Coto^409^,

Guillermo M Albaiceta^409^,

Marta García-Clemente^409^,

David Dalmau^410^,

Maria J Arranz^410^,

Beatriz Dietl^410^,

Alex Serra-Llovich^410^,

Pere Soler^411^,

Roger Colobrán^411^,

Andrea Martín-Nalda^411^,

Alba Parra Martínez^411^,

David Bernardo^412^,

Silvia Rojo^413^,

Aida Fiz-López^412^,

Elisa Arribas^412^,

Paloma de la Cal-Sabater^412^,

Tomás Segura^414^,

Esther González-Villa^415^,

Gemma Serrano-Heras^416^,

Joan Martí-Fàbregas^417^,

Elena Jiménez-Xarrié^417^,

Alicia de Felipe Mimbrera^418^,

Jaime Masjuan^418^,

Sebastian García-Madrona^418^,

Anna Domínguez-Mayoral^419^,

Joan Montaner Villalonga^419^,

Paloma Menéndez-Valladares^419^

**Women’s Genome Health Study (WGHS), i**.**e. the genomics subset of Women’s Health Study (WHS)**

**Analysis Team Lead**

Daniel I. Chasman^420,421^

**Data Collection Lead**

Howard D. Sesso^420,421^,

JoAnn E. Manson^420,421^

**Admin Team Lead**

Julie E. Buring^420,421^,

Paul M Ridker^420,421^

**Analysis Team Member**

Giulianini Franco^420^

**COVID-19 HGI corresponding authors**

**COVID-19 HGI corresponding authors**

Benjamin M Neale^4^,

Mark Daly^2,3,5^,

Andrea Ganna^2,3,5^

1. Yale University, New Haven, CT, USA
2. Institute for Molecular Medicine Finland (FIMM), Univerisity of Helsinki, Helsinki, Finland
3. Broad Institute of MIT and Harvard, Cambridge, MA, USA
4. Massachusetts General Hospital, Broad Institute of MIT and Harvard, Cambridge, MA, USA
5. Analytic and Translational Genetics Unit, Massachusetts General Hospital, Boston, MA, USA
6. Icahn School of Medicine at Mount Sinai, New York, NY, USA
7. Institute for Molecular Medicine Finland (FIMM), University of Helsinki, Helsinki, Finland
8. Icahn School of Medicine at Mount Sinai, Genetics and Genomic Sciences
9. Centre for Bioinformatics and Data Analysis, Medical University of Bialystok, Bialystok, Poland
10. University of Michigan, Ann Arbor, MI, USA
11. Ancestry, Lehi, UT, USA
12. Institute for Molecular Medicine Finland (FIMM), Helsinki, Finland
13. University of Liege, GIGA-Institute, Liège, Belgium
14. CHC Mont-Légia, Liège, Belgium
15. 5BHUL (Liège Biobank), CHU of Liège, Liège, Belgium
16. Institute for Molecular Medicine Finland, University of Helsinki, Helsinki, Finland
17. Analytic & Translational Genetics Unit, Massachusetts General Hospital, Boston, MA, USA
18. Stanley Center for Psychiatric Research, Broad Institute of MIT and Harvard, Cambridge, MA, USA
19. Program in Medical and Population Genetics, Broad Institute of MIT and Harvard, Cambridge, MA, USA
20. CHU of Liege, Liège, Belgium
21. University of Liege, Liège, Belgium
22. CHU of liege, Liège, Belgium
23. Department of Human Genetics, McGill University, Montréal, Québec, Canada
24. Lady Davis Institute, Jewish General Hospital, McGill University, Montréal, Québec, Canada
25. Kyoto-McGill International Collaborative School in Genomic Medicine, Graduate School of Medicine, Kyoto University, Kyoto, Japan
26. Research Fellow, Japan Society for the Promotion of Science
27. McGill Genome Centre and Department of Human Genetics, McGill University, Montréal, Québec, Canada
28. Department of Human Genetics, Epidemiology, Biostatistics and Occupational Health, McGill University, Montréal, Québec, Canada
29. Department of Twin Research, King’s College London, London, United Kingdom
30. Department of Epidemiology, Biostatistics and Occupational Health, McGill University, Montréal, Québec, Canada
31. Department of Emergency Medicine, McGill University, Montréal, Québec, Canada
32. Emergency Department, Jewish General Hospital, McGill University, Montréal, Québec, Canada
33. McGill AIDS Centre, Department of Microbiology and Immunology, Lady Davis Institute for Medical Research, Jewish General Hospital, McGill University, Montréal, Québec, Canada
34. McGill Centre for Viral Diseases, Lady Davis Institute, Department of Infectious Disease, Jewish General Hospital, Montréal, Québec, Canada
35. Research Centre of the Centre Hospitalier de l’Université de Montréal, Montréal, Canada
36. Department of Medicine,Research Centre of the Centre Hospitalier de l’Université de Montréal, Montréal, Canada
37. Department of Medicine, Université de Montréal, Montréal, Canada
38. Department of Medicine and Human Genetics, McGill University, Montréal, Québec, Canada
39. Department of Intensive Care, Research Centre of the Centre Hospitalier de l’Université de Montréal, Montréal, Canada
40. Division of Infectious Diseases, Research Centre of the Centre Hospitalier de l’Université de Montréal, Montréal, Canada
41. MRC Human Genetics Unit, Institute of Genetics and Cancer, University of Edinburgh, Western General Hospital, Edinburgh, UK EH4 2XU
42. Center for Applied Genomics, Children’s Hospital of Philadelphia, Philadelphia, PA, USA
43. Department of Pediatrics, Perelman School of Medicine, University of Pennsylvania, Philadelphia, PA, USA
44. Vanderbilt University Medical Center
45. Centre for Genomic and Experimental Medicine, Institute of Genetics and Cancer, University of Edinburgh, Western General Hospital, Edinburgh, UK, EH4 2XU, Usher Institute, University of Edinburgh, Nine, Edinburgh Bioquarter, 9 Little France Road, Edinburgh, UK, EH16 4UX
46. University of Texas Health
47. Centre for Genomic and Experimental Medicine, Institute of Genetics and Cancer, University of Edinburgh, Western General Hospital, Edinburgh, UK, EH4 2XU
48. Department of Psychology, University of Edinburgh, Edinburgh, UK, EH8 9JZ, Centre for Genomic and Experimental Medicine, Institute of Genetics and Cancer, University of Edinburgh, Western General Hospital, Edinburgh, UK, EH4 2XU
49. University of North Carolina at Chapel Hill
50. Center for Applied Genomics, The Children’s Hospital of Philadelphia, Philadelphia, PA, USA
51. Center for Applied Genomics, The Children’s Hospital of Philadelphia, Philadelphia, PA, USA
52. Division of Human Genetics, Department of Pediatrics, The Perelman School of Medicine, University of Pennsylvania, Philadelphia, PA, USA
53. Divisions of Human Genetics and Pulmonary Medicine, Department of Pediatrics, The Perelman School of Medicine, University of Pennsylvania, Philadelphia, PA, USA
54. Faculty of Medicine, University of Iceland, Reykjavik, Iceland
55. University of Colorado - Anschutz Medical Campus, Aurora, CO, USA
56. MRC Human Genetics Unit, Institute of Genetics and Cancer, University of Edinburgh, Western General Hospital, Edinburgh, Scotland
57. Centre for Global Health Research, Usher Institute, University of Edinburgh, Teviot Place, Edinburgh, Scotland
58. The Roslin Institute, The Royal (Dick) School of Veterinary Studies, University of Edinburgh, Edinburgh, UK
59. Department of Clinical Pathology, Faculty of Medicine, Mansoura University, Mansoura, Egypt.
60. Department of Biosciences, School of Science and Technology, Nottingham Trent University, Nottingham, UK
61. Genolier Innovation Network and Hub, Swiss Medical Network, Genolier Healthcare Campus, Route du Muids 3, 1272 Genolier, Switzerland.
62. Department of Pediatrics, Faculty of Medicine, Mansoura University, Mansoura, Egypt.
63. Department of Clinical Pathology, Faculty of Medicine, Tanta University, Tanta, Egypt.
64. Stanley Center for Psychiatric Research & Program in Medical and Population Genetics, Broad Institute of MIT and Harvard, Cambridge, MA, USA
65. Chest Department, Faculty of Medicine, Mansoura University, Mansoura, Egypt.
66. Anesthesia, Surgical Intensive Care & Pain Management Department, Faculty of Medicine, Tanta University, Tanta, Egypt.
67. Department of Medical Biochemistry, Faculty of Medicine, Mansoura University, Mansoura, Egypt.
68. Department of Surgery, Faculty of Medicine, Mansoura University, Mansoura, Egypt.
69. Department of Tropical Medicine, Faculty of Medicine, Mansoura University, Mansoura, Egypt.
70. pediatric and neonatology, Kafr Elzayat General Hospital, Kafr El-Zayat, Egypt.
71. Chest Department, Faculty of Medicine, Tanta University, Tanta, Egypt.
72. Pediatrics Department, Faculty of Medicine, Tanta University, Tanta, Egypt.
73. Department of Anaethesia and Critical Care, Faculty of Medicine, Mansoura University, Mansoura, Egypt.
74. Department of Internal Medicine, Faculty of Medicine, Tanta University, Tanta, Egypt.
75. Faculty of Science, Tanta University, Tanta, Egypt.
76. Department of Internal Medicine
77. Department of Epidemiology
78. Estonian Genome Centre, Institute of Genomics, University of Tartu, Tartu, Estonia
79. SYNLAB Estonia, University of Tartu, Tartu, Estonia
80. Kuressaare Hospital, Kuressaare, Estonia
81. University of Tartu, Tartu, Estonia
82. Institute of Biomedicine and Translational Medicine, University of Tartu
83. Institute of Biomedicine and Translational Medicine, University of Tartu, Tartu, Estonia
84. West Tallinn Central Hospital, Tallinn, Estonia
85. University of Tartu, Tartu University Hospital, Tartu, Estonia
86. Estonian Health Insurance Fund, Tallinn, Estonia
87. Tartu University Hospital, Tartu, Estonia
88. Department of Health Sciences, University of Leicester, Leicester, UK
89. Leicester NIHR Biomedical Research Centre, Leicester, UK
90. Department of Respiratory Sciences, University of Leicester, UK
91. Department of Genetics and Genome Biology, University of Leicester
92. FinnGen, Helsinki, Finland
93. Institute for Molecular Medicine Finland (FIMM), HiLIFE, University of Helsinki, Finland
94. Public Health, Faculty of Medicine, University of Helsinki, Finland 95.
95. Finnish Institute for Health and Welfare (THL), Helsinki, Finland
96. University of Helsinki, Faculty of Medicine, Clinical and Molecular Metabolism Research Program, Helsinki, Finland
97. Institute of Molecular and Clinical Ophthalmology Basel (IOB), Basel, Switzerland
98. Department of Ophthalmology, University of Basel, Basel, Switzerland
99. Infectious Diseases Service, Department of Medicine, University Hospital and University of Lausanne, Lausanne, Switzerland
100. Infectious Diseases Service, Department of Medicine, University Hospital, University of Lausanne, Lausanne, Switzerland
101. Centre for Primary Care and Public Health, University of Lausanne, Lausanne, Switzerland.
102. Division of Infectious Diseases and Hospital Epidemiology, Cantonal Hospital St Gallen, St Gallen, Switzerland.
103. Division of Intensive Care, Geneva University Hospitals and the University of Geneva Faculty of Medicine, Geneva, Switzerland.
104. Infectious Disease Service, Department of Internal Medicine, Geneva University Hospital, Geneva, Switzerland.
105. Clinique de Médecine et spécialités, Infectiologie, HFR-Fribourg, Fribourg, Switzerland
106. Infectious Diseases Division, University Hospital Centre of the canton of Vaud, hospital of Valais, Sion, Switzerland
107. Functional Host Genomics of Infectious Diseases, University Hospital and University of Lausanne, Lausanne, Switzerland
108. Registry COVID, University Hospital and University of Lausanne, Lausanne, Switzerland
109. Pneumonia prediction using lung ultrasound, University Hospital and University of Lausanne, Lausanne, Switzerland
110. Center for Primary Care and Public Health (Unisanté), University of Lausanne, Lausanne, Switzerland
111. Covid-19 Risk Prediction in Swiss ICUs-Trial, Division of Infectious Diseases and Hospital Epidemiology, Cantonal Hospital St Gallen, St Gallen, Switzerland.
112. GCAT-Genomes for Life, Germans Trias i Pujol Health Sciences Research Institute (IGTP), Crta. de Can Ruti, Cami de les Escoles s/n.08916 Badalona, Catalonia
113. Catalan Institute of Oncology,Bellvitge Biomedical Research Institute, Consortium for Biomedical Research in Epidemiology and Public Health and University of Barcelona, Barcelona, Spain
114. ISGlobal, Barcelona, Spain
115. IMIM (Hospital del Mar Medical Research Institute), Barcelona, Spain
116. Universitat Pompeu Fabra (UPF), Barcelona, Spain
117. CIBER Epidemiología y Salud Pública (CIBERESP), Madrid, Spain
118. Barcelona Supercomputing Center - Centro Nacional de Supercomputación (BSC-CNS).Life & Medical Sciences *currently at Programs in Metabolism and Medical and Population Genetics, Broad Institute of MIT and Harvard, Cambridge, MA, USA and Diabetes Unit and Center for Genomic Medicine, Massachusetts General Hospital, Boston, MA, USA. Harvard Medical School, Boston, Massachusetts, USA
119. Barcelona Supercomputing Center - Centro Nacional de Supercomputación (BSC-CNS).Life & Medical Sciences, Barcelona, Spain
120. University of Siena, DIISM-SAILAB, Siena, Italy
121. Université Côte d’Azur, Inria, CNRS, I3S, Maasai, Nice, France
122. Medical Genetics, University of Siena, Italy
123. Genetica Medica, Azienda Ospedaliero-Universitaria Senese, Italy
124. Med Biotech Hub and Competence Center, Department of Medical Biotechnologies, University of Siena, Siena, Italy
125. Division of Infectious Diseases and Immunology, Department of Medical Sciences and Infectious Diseases, Fondazione IRCCS Policlinico San Matteo, Pavia, Italy
126. Department of Internal Medicine and Therapeutics, University of Pavia, Italy
127. Department of Infectious and Tropical Diseases, University of Brescia and ASST Spedali Civili Hospital, Brescia, Italy
128. Chirurgia Vascolare, Ospedale Maggiore di Crema, Crema, Italy
129. III Infectious Diseases Unit, ASST-FBF-Sacco, Milan, Italy
130. Department of Biomedical and Clinical Sciences Luigi Sacco, University of Milan, Milan, Italy
131. Med Biotech Hub and Competence Center, Department of Medical Biotechnologies, University of Siena, Italy
132. Dept of Specialized and Internal Medicine, Tropical and Infectious Diseases Unit, Azienda Ospedaliera Universitaria Senese, Siena, Italy
133. Unit of Respiratory Diseases and Lung Transplantation, Department of Internal and Specialist Medicine, University of Siena, Siena, Italy
134. Dept of Emergency and Urgency, Medicine, Surgery and Neurosciences, Unit of Intensive Care Medicine, Siena University Hospital, Siena, Italy
135. Department of Medical, Surgical and Neuro Sciences and Radiological Sciences, Unit of Diagnostic Imaging, University of Siena, Siena, Italy
136. Rheumatology Unit, Department of Medicine, Surgery and Neurosciences, University of Siena, Policlinico Le Scotte, Siena, Italy
137. Department of Specialized and Internal Medicine, Infectious Diseases Unit, San Donato Hospital Arezzo, Arezzo, Italy
138. Dept of Emergency, Anesthesia Unit, San Donato Hospital, Arezzo, Italy
139. Department of Specialized and Internal Medicine, Pneumology Unit and UTIP, San Donato Hospital, Arezzo, Italy
140. Department of Emergency, Anesthesia Unit, Misericordia Hospital, Grosseto, Italy
141. Department of Specialized and Internal Medicine, Infectious Diseases Unit, Misericordia Hospital, Grosseto, Italy
142. Department of Preventive Medicine, Azienda USL Toscana Sud Est, Arezzo, Italy
143. Clinical Chemical Analysis Laboratory, Misericordia Hospital, Grosseto, Italy
144. Territorial Scientific Technician Department, Azienda USL Toscana Sud Est, Arezzo, Italy
145. Clinical Chemical Analysis Laboratory, San Donato Hospital, Arezzo, Italy
146. Department of Health Sciences, Clinic of Infectious Diseases, ASST Santi Paolo e Carlo, University of Milan, Milan, Italy
147. Department of Anesthesia and Intensive Care, University of Modena and Reggio Emilia, Modena, Italy
148. HIV/AIDS Department, National Institute for Infectious Diseases, IRCCS, Lazzaro Spallanzani, Rome, Italy
149. Infectious Diseases Clinic, Department of Medicine 2, Azienda Ospedaliera di Perugia and University of Perugia, Santa Maria Hospital, Perugia, Italy
150. Infectious Diseases Clinic, “Santa Maria” Hospital, University of Perugia, Perugia, Italy
151. Department of Infectious Diseases, Treviso Hospital, Treviso, Italy
152. Clinical Infectious Diseases, Mestre Hospital, Venezia, Italy
153. Infectious Diseases Clinic, ULSS1, Belluno, Italy
154. Medical Genetics and Laboratory of Medical Genetics Unit, A.O.R.N. “Antonio Cardarelli”, Naples, Italy
155. Department of Molecular Medicine and Medical Biotechnology, University of Naples Federico II, Naples, Italy
156. CEINGE Biotecnologie Avanzate, Naples, Italy
157. IRCCS SDN, Naples, Italy
158. Unit of Respiratory Physiopathology, AORN dei Colli, Monaldi Hospital, Naples, Italy
159. Division of Medical Genetics, Fondazione IRCCS Casa Sollievo della Sofferenza Hospital, San Giovanni Rotondo, Italy
160. Department of Medical Sciences, Fondazione IRCCS Casa Sollievo della Sofferenza Hospital, San Giovanni Rotondo, Italy
161. Infectious Diseases Clinic, Policlinico San Martino Hospital, IRCCS for Cancer Research, Genova, Italy
162. Microbiology, Fondazione Policlinico Universitario Agostino Gemelli IRCCS, Catholic University of Medicine, Rome, Italy
163. Department of Laboratory Sciences and Infectious Diseases, Fondazione Policlinico Universitario A. Gemelli IRCCS, Rome, Italy
164. Department of Cardiovascular Diseases, University of Siena, Siena, Italy
165. Otolaryngology Unit, University of Siena, Siena, Italy
166. Department of Internal Medicine, ASST Valtellina e Alto Lario, Sondrio, Italy
167. First Aid Department, Luigi Curto Hospital, Polla, Salerno, Italy
168. U.O.C. Laboratorio di Genetica Umana, IRCCS Istituto G. Gaslini, Genova, Italy
169. Infectious Diseases Clinics, University of Modena and Reggio Emilia, Modena, Italy
170. U.O.C. Medicina, ASST Nord Milano, Ospedale Bassini, Cinisello Balsamo (MI), Italy
171. Department of Cardiovascular, Neural and Metabolic Sciences, Istituto Auxologico Italiano, IRCCS, San Luca Hospital, Milan, Italy
172. Department of Medicine and Surgery, University of Milano-Bicocca, Milan, Italy
173. Istituto Auxologico Italiano, IRCCS, Center for Cardiac Arrhythmias of Genetic Origin, Milan, Italy
174. Istituto Auxologico Italiano, IRCCS, Laboratory of Cardiovascular Genetics, Milan, Italy
175. Unit of Infectious Diseases, ASST Papa Giovanni XXIII Hospital, Bergamo, Italy
176. Department of Cardiology,Istituti Clinici Scientifici Maugeri IRCCS, Institute of Montescano, Pavia, Italy
177. Istituti Clinici Scientifici Maugeri, IRCCS, Department of Cardiac Rehabilitation, Institute of Tradate (VA), Pavia, Italy
178. Cardiac Rehabilitation Unit, Fondazione Salvatore Maugeri, IRCCS, Scientific Institute of Milan, Milan, Italy
179. IRCCS C. Mondino Foundation, Pavia, Italy
180. Medical Genetics Unit, Meyer Children’s University Hospital, Florence, Italy
181. Department of Medicine, Pneumology Unit, Misericordia Hospital, Grosseto, Italy
182. Department of Preventive Medicine, Azienda USL Toscana Sud Est, Tuscany, Italy
183. Department of Anesthesia and Intensive Care Unit, ASST Fatebenefratelli Sacco, Luigi Sacco Hospital, Polo Universitario, University of Milan, Milan, Italy
184. Infectious Disease Unit, Hospital of Massa, Massa, Italy.
185. Department of Clinical Medicine, Public Health, Life and Environment Sciences, University of L’Aquila, L’Aquila, Italy
186. UOSD Laboratorio di Genetica Medica - ASL Viterbo, San Lorenzo, Italy
187. Department of Medical Sciences, Infectious and Tropical Diseases Unit, Azienda Ospedaliera Universitaria Senese, Siena, Italy
188. Unit of Infectious Diseases, S.M. Annunziata Hospital, Florence, Italy.
189. IRCCS Mondino Foundation, Pavia, Italy.
190. Infectious Disease Unit, Hospital of Lucca, Lucca, Italy
191. Health Management, Azienda USL Toscana Sudest, Tuscany, Italy
192. Department of Mathematics, University of Pavia, Pavia, Italy
193. Independent Researcher, Milan, Italy
194. Department of Electronics, Information and Bioengineering (DEIB), Politecnico di Milano, Milano, Italy
195. Scuola Normale Superiore, Pisa, Italy
196. CNR-Consiglio Nazionale delle Ricerche, Istituto di Biologia e Biotecnologia Agraria (IBBA), Milano, Italy
197. CNR-Consiglio Nazionale delle Ricerche, Istituto di Biologia e Biotecnologia Agraria (IBBA), Milano, Italy
198. Med Biotech Hub and Competence Center, Department of Medical Biotechnologies, University of Siena,Siena, Italy Italy
199. Core Research Laboratory, ISPRO, Florence, Italy
200. Genetica Medica, Azienda Ospedaliero-Universitaria Senese, Siena, Italy
201. Infectious Diseases Clinic, Department of Medicine 2, Azienda Ospedaliera di Perugia and University of Perugia, Santa Maria Hospital
202. Division of Infectious Diseases and Immunology, Fondazione IRCCS Policlinico San Matteo, Pavia, Italy
203. Fondazione IRCCS Ca’ Granda Ospedale Maggiore Policlinico
204. Department of Molecular and Translational Medicine, University of Brescia, Italy
205. Clinical Chemistry Laboratory, Cytogenetics and Molecular Genetics Section, Diagnostic Department, ASST Spedali Civili di Brescia, Brescia, Italy
206. Department of Medical and Surgical Sciences for Children and Adults, University of Modena and Reggio Emilia, Modena, Italy
207. Department of Molecular Medicine, University of Padova, Padua, Italy
208. Laboratory of Regulatory and Functional Genomics, Fondazione IRCCS Casa Sollievo della Sofferenza, San Giovanni Rotondo (Foggia), Italy
209. Clinical Trial Office, Fondazione IRCCS Casa Sollievo della Sofferenza Hospital, San Giovanni Rotondo, Italy
210. Department of Health Sciences, University of Genova, Genova, Italy
211. Oncologia Medica e Ufficio Flussi Sondrio, Sondrio, Italy
212. Local Health Unit-Pharmaceutical Department of Grosseto, Toscana Sud Est Local Health Unit, Grosseto, Italy
213. Department of Respiratory Diseases, Azienda Ospedaliera di Cremona, Cremona, Italy
214. Direzione Scientifica, Istituti Clinici Scientifici Maugeri IRCCS, Pavia, Italy
215. Fondazione per la ricerca Ospedale di Bergamo, Bergamo, Italy
216. Department of Cardiovascular, Neural and Metabolic Sciences,Istituto Auxologico Italiano, IRCCS, San Luca Hospital, Milan, Italy
217. Allelica Inc, New York, NY, USA
218. Dept of Specialized and Internal Medicine, Tropical and Infectious Diseases Unit, Azienda Ospedaliera Universitaria Senese, Siena, Italy
219. Department of Clinical and Experimental Medicine, Infectious Diseases Unit, University of Pisa, Pisa, Italy.
220. Infectious Disease Unit, Santo Stefano Hospital, AUSL Toscana Centro, Prato, Italy.
221. Clinic of Infectious Diseases, Catholic University of the Sacred Heart, Rome, Italy.
222. Medical Genetics, University of Siena, Siena, Italy
223. Infectious Disease Unit, Hospital of Lucca, Lucca Italy.
224. Department of Diagnostic and Laboratory Medicine, Institute of Biochemistry and Clinical Biochemistry, Fondazione Policlinico Universitario A. Gemelli IRCCS, Catholic University of the Sacred Heart, Rome, Italy.
225. MRC Human Genetics Unit, IGC,University of Edinburgh.EH4 2XU,UK
226. Medical Genetics Section, Centre for Genomic and Experimental Medicine, IGC, University of Edinburgh, Edinburgh, UK, Generation Scotland, Centre for Genomic and Experimental Medicine, IGC, University of Edinburgh, Edinburgh, UK
227. Department of Psychology, University of Edinburgh, Edinburgh, UK, Medical Genetics Section, Centre for Genomic and Experimental Medicine, IGC, University of Edinburgh, Edinburgh, UK, Generation Scotland, Centre for Genomic and Experimental Medicine, IGC, University of Edinburgh, Edinburgh, UK
228. Blizard Institute, Queen Mary University of London, London, United Kingdom
229. School of Basic and Medical Biosciences, Faculty of Life Sciences and Medicine, King’s College London, London, United Kingdom
230. Medical and Population Genomics, Wellcome Sanger Institute, Hinxton, UK
231. Bradford Institute for Health Research, Bradford Teaching Hospitals National Health Service (NHS) Foundation Trust, Bradford, UK
232. Blizard Institute, 4 Newark Street, Queen Mary University of London
233. Institute of Population Health Sciences, 4 Newark Street, Queen Mary University of London, London, United Kingdom
234. Genes & Health, Blizard Institute, Queen Mary University of London, E1 2AT, London, United Kingdom
235. Institute of Population Health Sciences, 4 Newark Street, Queen Mary University of London
236. Department of Biostatistics, University of Michigan, Ann Arbor, MI, USA
237. Digestive Oncology Research Center, Digestive Disease Research Institute, Shariati Hospital, Tehran University of Medical Sciences, Tehran, Iran
238. Department of Pulmonology, School of Medicine, Shariati Hospital, Tehran University of Medical Sciences, Tehran, Iran
239. Department of Critical Care Medicine, Noorafshar Hospital, Tehran, Iran
240. Department of Emergency Intensive Care Unit, School of Medicine, Shariati Hospital, Tehran University of Medical Sciences, Tehran, Iran
241. Department of Anesthesiology, School of Medicine, Amir Alam Hospital, Tehran University of Medical Sciences, Tehran, Iran
242. Department of Pulmonology, School of Medicine, Tehran University of Medical Sciences, Tehran, Iran
243. Department of Pathology, Parseh Pathobiology and Genetics Laboratory, Tehran, Iran
244. Department of Microbiology, Health and Family Research Center, NIOC Hospital, Tehran, Iran
245. Department of Emergency Medicine, School of Medicine, Shariati Hospital, Tehran University of Medical Sciences, Tehran, Iran
246. Department of Anesthesiology, School of Medicine, Tehran University of Medical Sciences, Tehran, Iran
247. Department of Pathology, Faculty of Medicine, Tehran Azad University, Tehran, Iran
248. Clinical Pharmacokinetics and Pharmacogenomics Research Unit, Faculty of Medicine, Chulalongkorn University, Bangkok Thailand
249. Department of Pharmacology, Faculty of Medicine, Chulalongkorn University, Bangkok Thailand
250. Thai Red Cross Emerging Infectious Diseases Clinical Centre, King Chulalongkorn Memorial Hospital, Bangkok, Thailand
251. Department of Pediatrics, Faculty of Medicine, Chulalongkorn University, Bangkok, Thailand.
252. Immunology Division, Department of Microbiology, Faculty of Medicine, Chulalongkorn University, Bangkok, Thailand
253. Center of Excellence in Immunology and Immune-mediated Diseases, Department of Microbiology, Faculty of Medicine, Chulalongkorn University, Bangkok, Thailand.
254. Clinical Pharmacokinetics and Pharmacogenomics Research Unit, Faculty of Medicine, Chulalongkorn University, Bangkok, Thailand
255. Department of Pathology, Faculty of Medicine, Nakornnayok, Srinakharinwirot University, Thailand
256. Center of Excellence for Medical Genomics, Medical Genomics Cluster, Department of Pediatrics, Faculty of Medicine, Chulalongkorn University, Bangkok, Thailand.
257. Excellence Center for Genomics and Precision Medicine, King Chulalongkorn Memorial Hospital, The Thai Red Cross Society, Bangkok, Thailand.
258. Department of Mathematics and Computer Science, Faculty of Science, Chulalongkorn University, Bangkok, Thailand
259. Omics Sciences and Bioinfomatics Center, Faculty of Science, Chulalongkorn University, Bangkok, Thailand
260. Research Affairs, Faculty of Medicine, Chulalongkorn University, Bangkok, Thailand
261. Center of Excellence for Medical Genomics, Medical Genomics Cluster, Faculty of Medicine, Chulalongkorn University, Bangkok, Thailand
262. Division of Infectious Disease, Department of Medicine, Faculty of Medicine, Chulalongkorn University, Bangkok, Thailand.
263. Center of Excellence in Pediatric Infectious Diseases and Vaccines, Chulalongkorn University, Bangkok, Thailand
264. Department of Microbiology, Faculty of Medicine, Chulalongkorn University, Bangkok, Thailand
265. Division of Infectious Diseases, Department of Medicine, Faculty of Medicine, Chulalongkorn University, Bangkok, Thailand
266. Healthcare-associated Infection Research Group STAR (Special Task Force for Activating Research), Chulalongkorn University, Bangkok, Thailand
267. K.G. Jebsen Center for Genetic Epidemiology, Department of Public Health and Nursing, NTNU, Norwegian University of Science and Technology, Trondheim, 7030, Norway
268. HUNT Research Center, Department of Public Health and Nursing, NTNU, Norwegian University of Science and Technology, Levanger, 7600, Norway
269. Clinic of Medicine, St. Olavs Hospital, Trondheim University Hospital, Trondheim, 7030, Norway
270. Division of Cardiovascular Medicine, Department of Internal Medicine, University of Michigan, Ann Arbor, MI, USA
271. Department of Computational Medicine and Bioinformatics, University of Michigan, Ann Arbor, MI, USA
272. Department of Human Genetics, University of Michigan, Ann Arbor, MI, USA
273. Analytic and Translational Genetics Unit, Massachusetts General Hospital, Boston, Massachusetts, USA
274. Program in Medical and Population Genetics, Broad Institute of Harvard and MIT, Cambridge, Massachusetts, USA
275. Gemini Center for Sepsis Research, Department of Circulation and Medical Imaging, NTNU, Norwegian University of Science and Technology, Trondheim, Norway
276. Department of Chronic Disease Epidemiology and Center for Perinatal, Pediatric and Environmental Epidemiology, Yale School of Public Health, New Haven, CT, USA
277. Clinic of Anaesthesia and Intensive Care, St. Olavs Hospital, Trondheim University Hospital, Trondheim, Norway
278. Department of Genetics, University Medical Centre Groningen, University of Groningen, Groningen, Netherlands
279. Department of Epidemiology, University Medical Centre Groningen, University of Groningen, Groningen, Netherlands
280. Department of Genetics, University Medical Centre Groningen, University of Groningen / Department of Genetics, University Medical Centre Utrecht, P.O. Box 85500, 3508 GA, Utrecht, The Netherlands
281. Department of Epidemiology, University of Groningen, University Medical Center Groningen, Groningen, The Netherlands
282. University of Groningen, University Medical Center Groningen, Department of Genetics, Groningen, The Netherlands
283. Department of Genetics, University Medical Center Groningen, Groningen, The Netherlands
284. Department of Psychiatry, University Medical Center Groningen, Groningen, The Netherlands
285. Center for Genomic Medicine, Massachusetts General Hospital, Boston, MA, USA
286. Programs in Metabolism and Medical and Population Genetics, Broad Institute of MIT and Harvard, Cambridge, MA, USA
287. Diabetes Unit and Center for Genomic Medicine, Massachusetts General Hospital, Boston, MA, USA. Harvard Medical School, Boston, Massachusetts, USA
288. Channing Division of Network Medicine, Department of Medicine, Brigham and Women’s Hospital, Boston, MA, USA
289. Brigham and Women’s Hospital, Boston, MA, USA
290. Psychiatric and Neurodevelopmental Genetics Unit, Center for Genomic Medicine, Massachusetts General Hospital, Boston, MA
291. Department of Neurology, Massachusetts General Hospital, Boston, MA, USA
292. Division of General Internal Medicine, Massachusetts General Hospital and Dpeartment of Medicine, Harvard Medical School and Program in Medical and Population Genetics, Broad Institute, Boston, MA, USA
293. Division of Genetics, Department of Medicine, Brigham and Women’s Hospital, Broad Institute of MIT and Harvard, Harvard Medical School, Boston, MA, USA
294. Division of Genetics, Department of Medicine, Brigham and Women’s Hospital, Boston, MA, USA
295. Seaver Autism Center for Research and Treatment
296. Department of Psychiatry
297. Icahn School of Medicine at Mount Sinai, New York, NY 10029, USA
298. Mount Sinai Clinical Intelligence Center, Department of Genetics and Genomic Sciences, Icahn School of Medicine at Mount Sinai, New York, NY, 10029, USA
299. Department of Genetics and Genomic Sciences, Icahn School of Medicine at Mount Sinai, New York, NY, 10029, USA
300. Sema4, a Mount Sinai venture, Stamford CT, 06902, USA
301. Seaver Autism Center for Research and Treatment, Department of Psychiatry, Icahn School of Medicine at Mount Sinai, New York, NY 10029, USA
302. Mount Sinai Clnical Intelligence Center, Charles Bronfman Institute for Personalized Medicine
303. Department of Genetics & Genomic Sciences, Icahn School of Medicine at Mount Sinai, New York, NY, 10029, USA
304. Mount Sinai Clinical Intelligence Center, Department of Genetics and Genomic Sciences, Icahn School of Medicine at Mount Sinai, New York, NY 10029, USA
305. Icahn Institute of Data Science and Genomics Technology, New York, NY 10029, USA
306. Mount Sinai Clinical Intelligence Center, New York, NY 10029, USA
307. Department of Genetics and Genomic Sciences, Icahn School of Medicine at Mount Sinai, New York, NY, USA
308. Charles Bronfman Institute for Personalized Medicine, Icahn School of Medicine at Mount Sinai, New York, NY, 10029, USA
309. Institute for Genomic Health, Icahn School of Medicine at Mount Sinai, New York, NY, USA
310. The Charles Bronfman Institute for Personalized Medicine, Icahn School of Medicine at Mount Sinai, New York, NY 10029, USA
311. The Mindich Child Health and Development Institute, Icahn School of Medicine at Mount Sinai, New York, NY 10029, USA
312. Pamela Sklar Division of Psychiatric Genomics, Department of Psychiatry, Department of Genetic and Genomic Sciences
313. Pamela Sklar Division of Psychiatric Genomics, Department of Psychiatry, Department of Genetic and Genomic Sciences, Icahn School of Medicine at Mount Sinai, New York, NY 10029, USA
314. Seaver Autism Center for Research and Treatment, Department of Psychiatry
315. Mount Sinai Clinical Intelligence Center, Department of Psychiatry, Department of Genetic and Genomic Sciences, Icahn School of Medicine at Mount Sinai, NY, 10029, USA
316. Department of Environmental Medicine and Public Health, Icahn School of Medicine at Mount Sinai, New York, NY 10029, USA
317. Mount Sinai Clinical Intelligence Center
318. The Hasso Plattner Institute of Digital Health at Mount Sinai
319. BioMe Phenomics Center, Icahn School of Medicine at Mount Sinai, New York, NY 10029, USA
320. Department of Medicine, Icahn School of Medicine at Mount Sinai, New York, NY 10029, USA
321. Pamela Sklar Division of Psychiatric Genomics, Seaver Autism Center for Research and Treatment, Department of Psychiatry, Department of Genetic and Genomic Sciences, Icahn School of Medicine at Mount Sinai, New York, NY 10029, USA
322. Regeneron Genetics Center, Tarrytown, NY, USA
323. Phenomic Analytics & Clinical Data Core, Geisinger Health System, Danville, PA, USA
324. Department of Population Health Sciences, Geisinger Health System, Danville, PA, USA
325. Department of Molecular and Functional Genomics, Geisinger Health System, Danville, PA, USA
326. Vrije Universiteit Amsterdam, Amsterdam, UK
327. Department of Genetics, University of Pennsylvania Perelman School of Medicine, Philadelphia, PA, USA
328. Department of Biomedical Data Science, Stanford University, Stanford, CA, USA
329. Genomics Research Department, Saudi Human Genome Project, King Fahad Medical City and King Abdulaziz City for Science and Technology, Riyadh, Saudi Arabia
330. Developmental Medicine Department, King Abdullah International Medical Research Center, King Saud Bin Abdulaziz University for Health Sciences, Ministry of National Guard-Health Affairs, Riyadh, Kingdom of Saudi Arabia and Saudi Human Genome Project (SHGP), King Abdulaziz City for Science and Technology (KACST), Satellite Lab at King Abdulaziz Medical City (KAMC), Ministry of National Guard Health Affairs (MNG-HA), Riyadh, Kingdom of Saudi Arabia.
331. College of Applied Medical Sciences, Taibah University, Madina, Saudi Arabia
332. Developmental Medicine Department, King Abdullah International Medical Research Center, King Saud Bin Abdulaziz University for Health Sciences, Ministry of National Guard Health Affairs, Riyadh, Saudi Arabia
333. Life Science and environmental institute, King Abdulaziz City for Science and Technology, Riyadh, Saudi Arabia
334. The Liver Transplant Unit, King Faisal Specialist Hospital and Research Centre, Riyadh, Saudi Arabia. The Division of Gastroenterology and Hepatology, Johns Hopkins University
335. Department of Pathology, College of Medicine, King Saud University, Riyadh, Saudi Arabia
336. Titus Family Department of Clinical Pharmacy, USC School of Pharmacy University of Southern California
337. KACST-BWH Centre of Excellence for Biomedicine, Joint Centers of Excellence Program, King Abdulaziz City for Science and Technology (KACST), Riyadh, Kingdom of Saudi Arabia
338. Ministry of the National Guard Health Affairs, King Abdullah International Medical Research Center and King Saud Bin Abdulaziz University for Health Sciences, Riyadh, Kingdom of Saudi Arabia
339. Ohud Hospital, Ministry of Health, Madinah,Saudi Arabia
340. Pediatric Infectious Diseases, Children’s Specialized Hospital, King Fahad Medical City, Riyadh, Saudi Arabia
341. The Saudi Biobank, King Abdullah International Medical Research Center, King Saud bin Abdulaziz University for Health Sciences, Ministry of National Guard Health Affairs, Riyadh, Saudi Arabia
342. Developmental Medicine Department, King Abdullah International Medical Research Center, King Saud Bin Abdulaziz University for Health Sciences, King AbdulAziz Medical City, Ministry of National Guard Health Affairs, Riyadh, Kingdom of Saudi Arabia
343. Department of Pathology and Laboratory Medicine, King Abdulaziz Medical City, Ministry of National Guard Health Affairs, King Saud Bin Abdulaziz University for Health Sciences and King Abdullah International Medical Research Center, Riyadh, Saudi Arabia
344. Laboratory Department, Security Forces Hospital, General Directorate of Medical Services, Ministry of Interior
345. King Abdulaziz City for Science and Technology (KACST), Riyadh, Kingdom of Saudi Arabia
346. Department of Developmental Medicine, King Abdullah International Medical Research Center, King Saud Bin Abdulaziz University for Health Sciences, King Abdulaziz Medical City, Ministry of National Guard Health Affairs, Riyadh, Saudi Arabia
347. Titus Family Department of Clinical Pharmacy, USC School of Pharmacy
348. Department of Clinical Laboratory Sciences, College of Applied Medical Sciences, King Saud University, Riyadh, Saudi Arabia.
349. Department of Neuroscience, Karolinska Institutet, Stockholm, Sweden
350. Max Planck Institute for Evolutionary Anthropology, Leipzig, Germany
351. Stanley Center for Psychiatric Research & Program in Medical and Population Genetics
352. Anaesthesiology and Intensive Care Medicine, Department of Surgical Sciences, Uppsala University, Uppsala, Sweden
353. Integrative Physiology, Department of Medical Cell Biology, Uppsala University, Uppsala, Sweden
354. Hedenstierna Laboratory, CIRRUS, Anaesthesiology and Intensive Care Medicine, Department of Surgical Sciences, Uppsala University, Uppsala, Sweden
355. Division Anesthesiology and Intensive Care, CLINTEC, Karolinska Institutet, Stockholm, Sweden
356. Department of Genetics and Bioinformatics, Norwegian Institute of Public Health, Oslo, Norway.
357. Centre for Fertility and Health, Norwegian Institute of Public Health, Oslo, Norway.
358. Department of Method Development and Analytics, Norwegian Institute of Public Health, Oslo, Norway.
359. King’s College London, Department of Twin Research and Genetic Epidemiology, London, UK
360. NIHR Biomedical Research Centre at Guy’s and St Thomas’ Foundation Trust, London, UK
361. Genomics England
362. Queen Mary University, London, United Kingdom
363. William Harvey Research Institute, Barts and the London School of Medicine and Dentistry, Queen Mary University of London, London EC1M 6BQ, UK
364. UCL Great Ormond Street Institute of Child Health, London, United Kingdom
365. Genomics England, London, United Kingdom
366. University of Cambridge, London, United Kingdom
367. Department of Human Genetics, McGill University, Montréal, Québec, Canada. Lady Davis Institute, Jewish General Hospital, McGill University, Montréal, Québec, Canada. Kyoto-McGill International Collaborative School in Genomic Medicine, Graduate School of Medicine, Kyoto University, Kyoto, Japan. Research Fellow, Japan Society for the Promotion of Science
368. Big Data Institute, Nuffield Department of Population Health, University of Oxford, Li Ka Shing Centre for Health Information and Discovery, Old Road Campus, Oxford, OX3 7LF, United Kingdom
369. Genomics PLC, King Charles House, Park End Street, Oxford, OX1 1JD, United Kingdom
370. Nuffield Department of Medicine, Experimental Medicine Division, University of Oxford, John Radcliffe Hospital, Oxford, OX3 9DU, United Kingdom
371. Public Health England, Field Service, Addenbrooke’s Hospital, Cambridge, CB2 0QQ, United Kingdom
372. Public Health England, Data and Analytical Services, National Infection Service, London, NW9 5EQ, United Kingdom
373. Program in Bioinformatics and Integrative Genomics, Harvard Medical School, Boston, MA, USA
374. Program in Biological and Biomedical Sciences, Harvard Medical School, Boston, MA, USA
375. Wellcome Centre for Human Genetics, University of Oxford, Roosevelt Drive, Oxford, OX3 7BN, United Kingdom
376. Department of Clinical Research and Leadership, George Washington University, Washington, DC, USA
377. Department of Human Genetics, The Wellcome Sanger Institute, Wellcome Genome Campus, Hinxton, Cambridge, CB10 1HH, UK
378. The National Institute for Health Research Blood and Transplant Unit in Donor Health and Genomics, University of Cambridge, Strangeways Research Laboratory, Wort’s Causeway, Cambridge, CB1 8RN, UK
379. Department of Haematology, University of Cambridge, Cambridge Biomedical Campus, Long Road, Cambridge, CB2 0PT, UK
380. British Heart Foundation Cardiovascular Epidemiology Unit, Department of Public Health and Primary Care, University of Cambridge, Cambridge, UK
381. British Heart Foundation Centre of Research Excellence, University of Cambridge, Cambridge, UK
382. National Institute for Health Research Blood and Transplant Research Unit in Donor Health and Genomics, University of Cambridge, Cambridge, UK
383. Health Data Research UK Cambridge, Wellcome Genome Campus and University of Cambridge, Cambridge, UK
384. Department of Human Genetics, Wellcome Sanger Institute, Hinxton, UK
385. Department of Epidemiology, Emory University Rollins School of Public Health
386. Atlanta CA Health Care System, North Druid Hills, GA, USA
387. Center for Population Genomics, MAVERIC, VA Boston Healthcare System, Boston, MA, USA
388. MAVERIC, VA Boston Healthcare System, Boston, MA, USA
389. Stanford University
390. Palo Alto VA Healthcare System, Stanford, CA, USA
391. Department of Biostatistics, Boston Univeristy School of Public Health
392. Vanda Pharmaceuticals Inc.
393. Stroke Pharmacogenomics and Genetics, Biomedical Research Institute Sant Pau, Sant Pau Hospital, Barcelona, Spain
394. Institute for Biomedical Researhc of Barcelona (IIBB), National Spanish Research Council (CSIC)
395. Institut d’Investigacions Biomèdiques August Pi i Sunyer (IDIBAPS), Barcelona, Spain
396. Institute of Biomedicine of Valencia (IBV), CSIC, València, Spain
397. Network Center for Biomedical Research on Neurodegenerative Diseases (CIBERNED), València, Spain
398. Neurology and Genetic Mixed Unit, La Fe Helath Research Institute, València, Spain
399. Institute for Biomedical Research of Barcelona (IIBB), National Spanish Research Council (CSIC)
400. Department of Neurology, Hospital Universitari MútuaTerrassa, Fundació Docència i Recerca MútuaTerrassa, Terrassa, Spain
401. Department of Molecular and Cell Biology, Centro Nacional de Biotecnología (CNB-CSIC), Campus Universidad Autónoma de Madrid, 28049 Madrid, Spain
402. Instituto de Física de Cantabria (IFCA-CSIC)
403. Hospital Clínic, IDIBAPS, Barcelona, Spain
404. Hospital Clínic, Barcelona, Spain
405. Hospital Clínic, IDIBAPS, School of Medicine, University of Barcelona Barcelona, Spain
406. IDIBAPS, Barcelona, Spain
407. IIBB-CSIC, Barcelona, Spain
408. Servicio de Salud del Principado de Asturias, Oviedo, Spain
409. Hospital Mutua de Terrassa, Terrassa, Spain
410. Hospital Valle Hebrón, Barcelona, Spain
411. Instituto de Biomedicina y Genética Molecular (IBGM), CSIC-Universidad de Valladolid, Spain
412. Hospital Clínico Universitario de Valladolid (SACYL)
413. Department of Neurology, University Hospital of Albacete.
414. Department of Neurology
415. Research Unit, University Hospital of Albacete
416. Department of Neurology, Biomedical Research Institute Sant Pau (IIB Sant Pau), Hospital de la Santa Creu i Sant Pau, Barcelona (Spain)
417. Hospital Universitario Ramón Y Cajal, IRYCIS, Madrid, Spain
418. Institute de Biomedicine of Seville, IBiS/Hospital Universitario Virgen del Rocío/CSIC/University of Seville & Department of Neurology, Hospital Universitario Virgen Macarena, Seville.
419. Brigham and Women’s Hospital, Boston, USA
420. Harvard Medical School, Boston, MA

