## SupplementaryMethodsandFigures for "Mapping the human genetic architecture of COVID-19: an update"

**METHODS**

Ethical statements for each contributing study are given in Supplementary Table 1.

**Probabilistic assignment of variants into susceptibility vs. severity effects**

We consider an assignment of variants reported in **Supplementary Table 3** into two groups based on whether they seem to affect susceptibility to corona virus infection (INF) or severity of the infection (SEV). For this analysis, we meta-analyzed GWAS summary statistics from such versions of B2 (hospitalized for COVID vs. population) and C2 (infected vs. population) GWAS that only included those studies that had contributed some data to B2. Therefore, all studies included in this analysis had made some effort to distinguish from all the infected individuals those individuals who additionally were hospitalized. The sample sizes of these GWAS were 23,988 cases / 2,834,885 controls for B2 and 114,516 cases / 2,138,237 controls for C2. This corresponds to an effective samples size of 84,292 for the hospitalized analysis and 404,416 for the reported infection analysis.

We assume that all hospitalized cases (B2 cases) were included among the infected cases (C2 cases) of the corresponding study and that, for each study, the controls of B2 and C2 had a maximum overlap possible given the control counts in the two data sets.

Next, we explain how we defined the statistical models that represent INF and SEV, and how we compared these models at each SNP.

Intuitively, INF represents a variant that associates with susceptibility of infection but has no effect on the severity of infection. The allele frequency of such a variant is similar among the hospitalized cases (B2 cases) as it is among all infected (C2 cases). Thus, under the INF model, we assume similar effect size between C2 and B2, i.e., $\beta_{C2}\approx\beta_{B2}$.

Model SEV represents a variant that affects severity of infection $(\left| \beta_{B2} \right|>0$) but not susceptibility to infection. If the cases of our susceptibility scan C2 were a random subset of infected individuals, then under the SEV model, $\beta_{C2}\approx0$. However, since our C2 cases are strongly enriched for severe cases, we expect that in our data also ${|\beta}_{C2}|>0$ even when a variant is affecting severity of infection but not susceptibility to infection. We expect that, for such variants, the effect size in C2 is proportional to its effect in B2, i.e., $\beta_{C2}\approx w_{SEV}\beta_{B2}$, where the constant of proportionality, *w_SEV_* < 1, depends on the proportion of all C2 cases that are also B2 cases. If we imagine C2 analysis as a fixed-effect meta-analysis between B2 and an (imaginary) “non-severe infection vs. population” analysis that had no sample overlap with B2 analysis, then $w_{SEV}={n_{B2}^{(eff)}}/{n_{C2}^{(eff)}}$, where $n_{i}^{(eff)}=R_{i}S_{i}/N_{i}$ is the effective sample size of study *i* with *R_i_* the number of controls, *S_i_* the number of cases and *N_i_* = *R_i_* + *S_i_*. In our data, ${n_{B2}^{(eff)}}/{n_{C2}^{(eff)}}\approx0.208,$where the effective sample sizes are computed by summing the effective sample sizes over individual studies of B2 and C2 analyses. Even when controls of B2 analysis and the imaginary “non-severe infection vs. population” analysis overlapped completely, the value of *w_SEV_* would change little in these data. After accounting for the overlap in controls, we estimated a value of *w_SEV_* = 0.20 that we used in the analyses described below.

To derive correlation *r_B2C2_* between the effect size estimators of B2 and C2 analyses due to overlapping samples, we used formula

$$r_{B2C2}=\frac{\sum_{k=1}^{K} \sqrt{n_{B2,k}^{(eff)} n_{C2,k}^{(eff)}}r_{B2C2,k}}{\sqrt{\sum_{k=1}^{K} n_{B2,k}^{(eff)}\sum_{k=1}^{K} n_{C2,k}^{(eff)}}}$$

where subscript *k* refers to individual studies. The correlation *r_B2C2,k_* between B2 and C2 for study *k* is computed as in Bhattacharjee et al.^1^:

$r_{B2C2,k}=\sqrt{n_{B2,k}^{(eff)} n_{C2,k}^{(eff)}}\left( \frac{S_{B2C2,k}}{S_{B2,k}S_{C2,k}}+\frac{R_{B2C2,k}}{R_{B2,k}R_{C2,k}} \right)$

where *S_B2C2,k_* is the number of shared B2 and C2 cases in study k and similarly *R_B2C2,k_* is the number of shared controls. By applying this to the data, we estimated *r_B2C2_* = 0.45.

With these estimates of *w_SEV_* = 0.2 and *r_B2C2_* = 0.45*,* and with the observed data at any one SNP containing effect estimates $(\hat{\beta}_{B2},\hat{\beta}_{C2})$ and their standard errors $\left( s_{B2}, s_{C2} \right)$, we derive the two models, INF and SEV, as follows.

Prior distribution for effects is zero-centered bivariate normal distribution

$\binom{\beta_{B2}}{\beta_{C2}}\sim N\left( \binom{0}{0},\Theta_{i} \right),$ with

$\Theta_{INF}=\tau^{2}\left( \begin{matrix} 1 & {1-\eta}_{INF} \\ 1-\eta_{INF} & 1 \end{matrix} \right)$ and

$\Theta_{SEV}=\tau^{2}\left( \begin{matrix} 1 & (1-\eta_{SEV})w_{SEV} \\ (1-\eta_{SEV})w_{SEV} & w_{SEV}^{2} \end{matrix} \right)$.

We have used value $\tau=0.1$ to define the expected effect sizes of the B2 analysis (implying roughly that 95% of the true effect sizes of the risk variants have odds-ratio below 1.2). By tuning the parameters $\eta_{INF}$ and $\eta_{SEV}$, we can define how much deviation real effects can have from the theoretical relationships $\beta_{C2}=\beta_{B2}$ and $\beta_{C2}=w_{SEV}\beta_{B2}$ corresponding to models INF and SEV, respectively. We have set these values in such a way that, under both models, the mean (Euclidean) distance between the effect size and the corresponding line is 0.0025 and 95% of the effects are within 0.006 units from the line. This happens when $\eta_{INF}=0.001$ and $\eta_{SEV}=0.013$.

The likelihood for the observed data under both models is Gaussian

$$\binom{\hat{\beta}_{B2}}{\hat{\beta}_{C2}}\sim N\left( \binom{\beta_{B2}}{\beta_{C2}},\Sigma\right), \text{where} \Sigma=\left( \begin{matrix} s_{B2}^{2} & s_{B2}s_{C2}r_{B2C2} \\ s_{B2}s_{C2}r_{B2C2} & s_{C2}^{2} \end{matrix} \right).$$

It follows from Trochet et al.^2^ that we can analytically integrate the likelihood with respect to the prior distributions and the resulting marginal likelihood for each model is proportional to a Gaussian density function evaluated at the observed effect size estimates as

$\Pr\left( \text{DATA} \right|\text{Model} i) \propto f_{N}\left( \binom{\hat{\beta}_{B2}}{\hat{\beta}_{C2}}; \binom{0}{0},\Theta_{i}+\Sigma\right)$.

We set equal prior probability on each model (i.e. 50% on INF and 50% on SEV), and consequently the posterior probabilities of the models will be proportional to their marginal likelihoods. These posterior probabilities are reported in Supplementary Table 3.

A limitation of this approach is that it classifies every variant between the two fixed models INF and SEV without considering a possibility that the variant might not fit either of these two models very well. We have chosen this approach since, based on the data shown in **Supplementary Figure 4**, a large majority of the variants are well aligned with either INF or SEV model. Consequently, we kept this model comparison simple and only between INF and SEV rather than complicated it by inclusion of some additional models that would have little support from the observed data.

**Gene prioritization and Phenome wide association study (PheWAS)**

The variant annotation and PheWAS were performed as per methods described in Niemi et. al 2021^3^.

**Genetic Correlations and Mendelian Randomization**

The genetic correlation, heritability estimates, and Mendelian Randomization were performed as per methods described in Niemi et. al 2021^3^.

SNP heritability for all three COVID-19 related phenotypes was low (<1% on the observed scale; Supplementary Table 7). To understand which traits are genetically correlated and/or potentially causally related to the three phenotypes, we first estimated genetic correlations with 38 traits (Supplementary Table 8). In addition to what was previously reported, we found positive genetic correlations with two risk factors (depression and insomnia symptoms), two biomarkers (C-reactive protein (CRP) and 25-hydroxyvitamin D), and two disease liability traits (asthma and heart failure).

We next applied two-sample Mendelian Randomization (MR) to infer potential causal relationships between COVID-19 related phenotypes and their genetically correlated traits. Four traits (BMI, type II diabetes, red blood cell count, and height) showed evidence of causal associations after correcting for multiple testing and were robust to potential violations of the underlying assumptions of MR (Supplementary Table 9).

Multivariable Mendelian Randomization (MVMR) was used to estimate the direct effects of body mass index (BMI) and type II diabetes (T2D) on the risk of SARS-CoV-2 phenotypes, by including both exposures within the same model. We selected all independent (r^2^ = 0.001; kb = 10000) genome-wide significant (p < 5x10^-8^) SNPs associated with BMI and type II diabetes; and using the full list of SNPs associated with both BMI and T2D, performed a second clumping procedure (r^2^ = 0.001; kb = 10000) to obtain independent SNPs. After clumping the full list of SNPs from both BMI and T2D and restricting to SNPs found in the SARS-CoV-2 GWASs, a total of 721 SNPs were available for multivariable MR (516 were associated with BMI only, 201 were associated with T2D only, and 4 SNPs overlap between both GWAS). MVMR was performed using the “MVMR” package [1]. The sensitivity analyses conducted using “MVMR” require estimates of the pairwise covariances between each instrument and each exposure, as such, we used the phenotypic correlation between BMI and T2D and summary data to generate estimates of the covariances. Phenotypic correlations between BMI and T2D were estimated from the LDSC regression intercept (r^2^ = 0.13) ^4^. Next, we calculated conditional F-statistics to evaluate the presence of weak instruments. The conditional F-statistic for both BMI and T2D were >10, indicating that the selected instruments were strongly associated with their corresponding exposure (Supplementary Table 10b) ^5,6^. We then assessed multivariable instrument pleiotropy using the modified Cochran’s Q-statistic that accounts for potential weak instrument bias, with evidence of heterogeneity indicative of a violation in the escalation restriction assumption in MR ^5,6^ . For each of the SARS-CoV-2 phenotypes MVMR models there was evidence of heterogeneity, suggesting the causal estimates from IVW-MVMR may be biased due to the presence of horizontal pleiotropy (Supplementary Table 10c). Inverse weighted multivariable MR model was used to estimate the direct effect of BMI and T2D upon each of the SARS-CoV-2 phenotypes (Supplementary Table 10). When MVMR assumptions are violated, as indicated by the presence of weak instruments or horizontal pleiotropy, it is possible to obtain more robust causal estimates Q- statistic to minimization (Supplementary Table 10).

Code for implementing the MVMR analysis is available at: <https://github.com/marcoralab/multivariate_MR>

**SUPPLEMENTARY FIGURES**

**Supplementary Figure 1**

**Analytical summary of the COVID-19 HGI meta-analysis**. Using the analytical plan set by the COVID-19 HGI, each individual study runs their analyses and uploads the results to the Initiative, who then runs the meta-analysis. There are three main analyses that each study can contribute summary statistics to: critically ill COVID-19, hospitalized COVID-19 and reported SARS-CoV-2 infection. The phenotypic criteria used to define cases are listed in the dark grey boxes, along with the numbers of cases (N) included in the final all ancestries meta-analysis. Controls were defined in the same way across all three analyses: as everybody that is not a case e.g. population controls (light grey box). Sensitivity analyses, not reported in this Figure, also used mild/asymptomatic COVID-19 cases as controls. Sample number (N) of controls differed between the analyses due to the difference in number of studies contributing data to these.

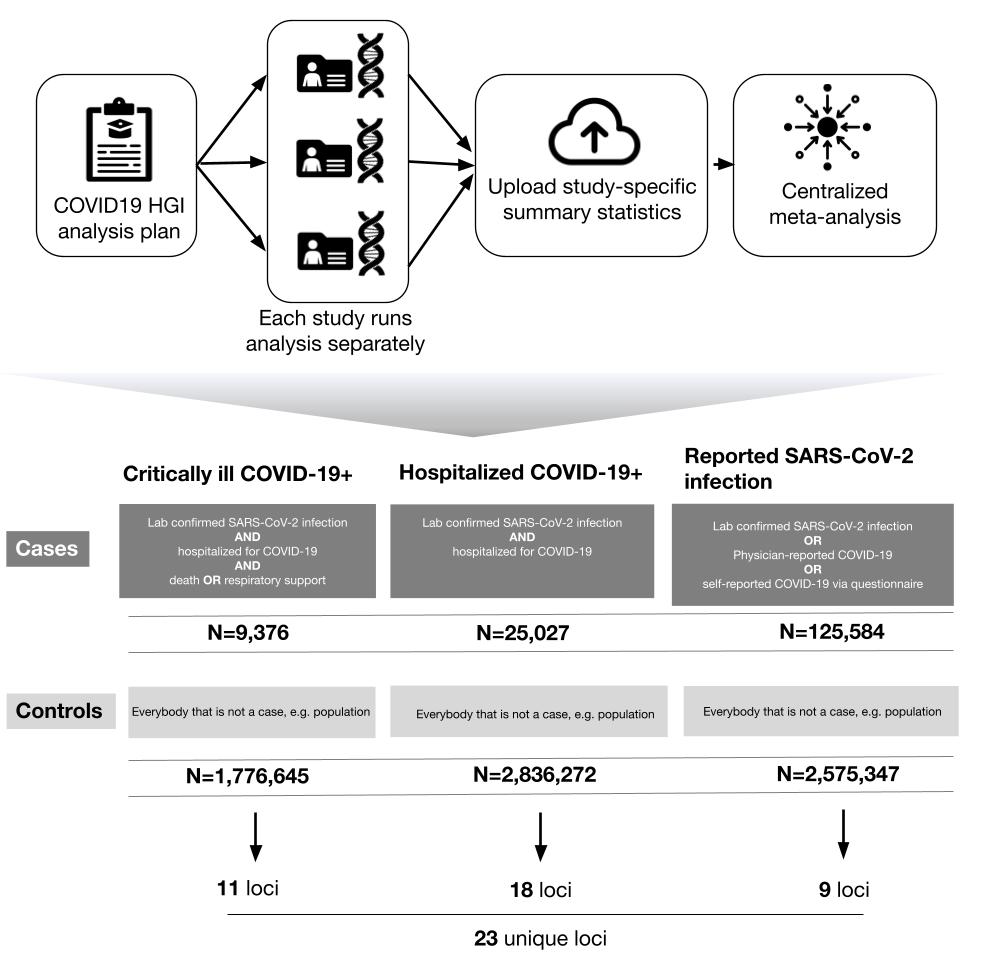

**Supplementary Figure 2**

**Genome-wide association results for COVID-19 critical illness (Release 6).** Results of genome-wide association study of critically ill COVID-19 cases vs population controls (n=9,376 cases and n=1,776,645 controls). Critically ill COVID-19 cases defined as those who required respiratory support in hospital or who were deceased due to the disease.

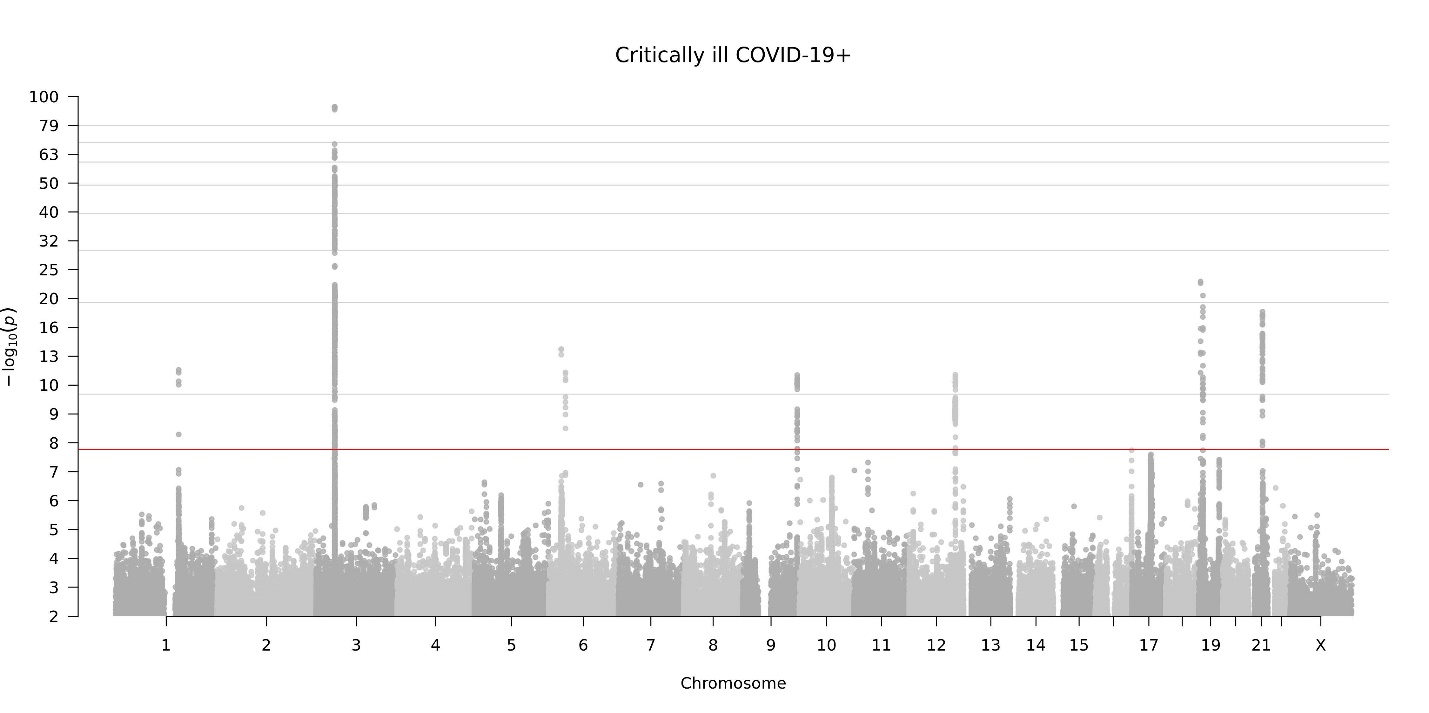

**Supplementary Figure 3**

**Comparison of associations between release 5 and release 6.** a. 𝜒2 statistics (y-axis) and effective sample size (x-axis) in release 5 (dots labelled as R5) and release 6 (dots labelled as R6), for each variant reported in release 6 and rs72711165. b. slope of the line represented in panel a, that is, the change in 𝜒2 statistics for each additional unit in the effective sample size, for each variant reported in release 6 and rs72711165. c. effect size absolute value in release 5 (x-axis) and release 6 (y-axis) and standard error, for each variant reported in release 6 and rs72711165.

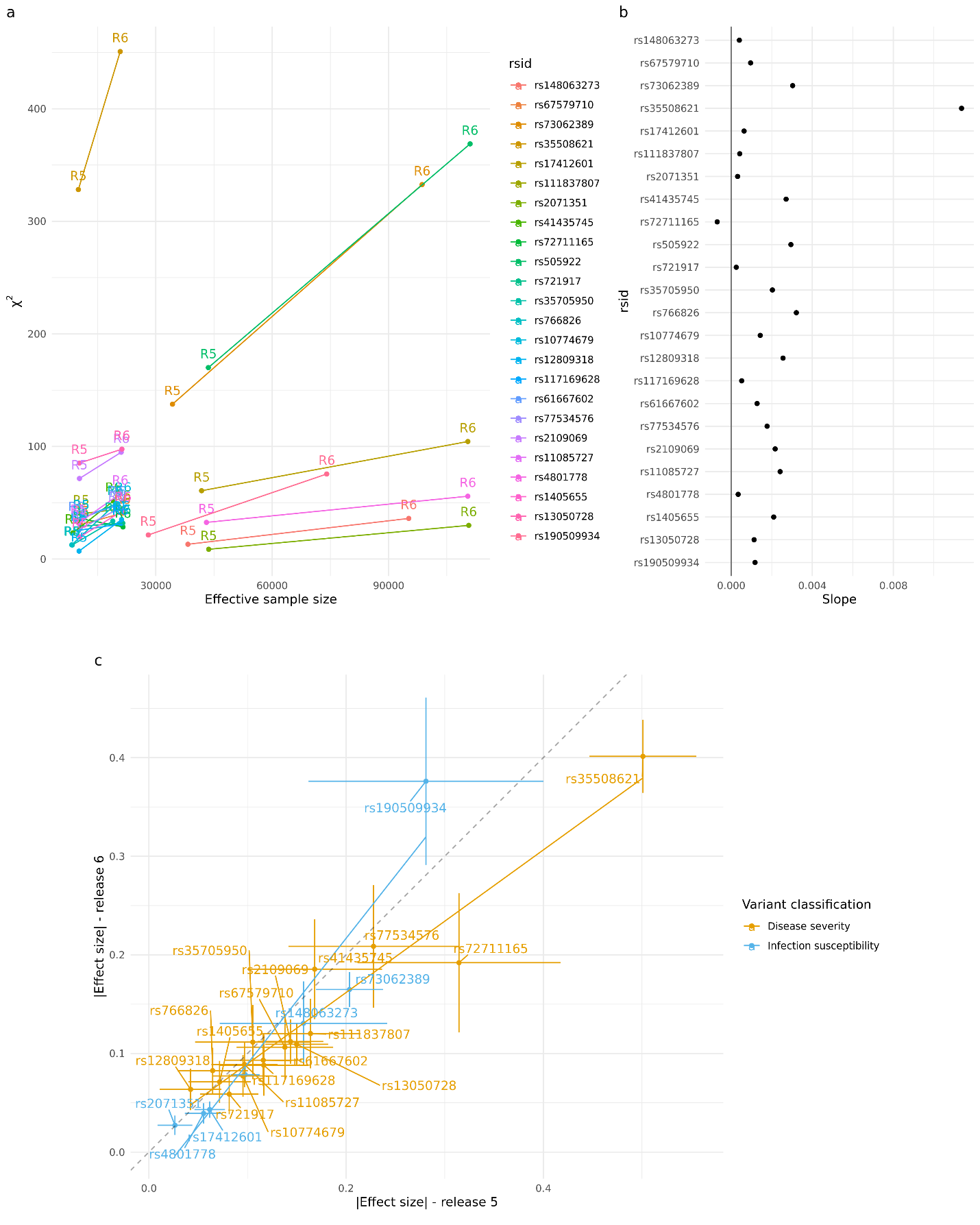

**Supplementary Figure 4**

**The estimated log odds ratios for 23 lead variants shown for B2 GWAS (hospitalized vs. controls) on x-axis and C2 GWAS (infected vs. controls) on y-axis.** The assignment to variants affecting susceptibility to infection (INF) or disease severity (SEV) is shown by colors and corresponds to Supplementary Table 3. The two lines show the expected relationship between effect sizes of variants affecting susceptibility to infection (line y = x) or disease severity (line y = 0.2 x). In left panel, the variants have been annotated by rsids, and in right panel, 95% confidence intervals are shown.

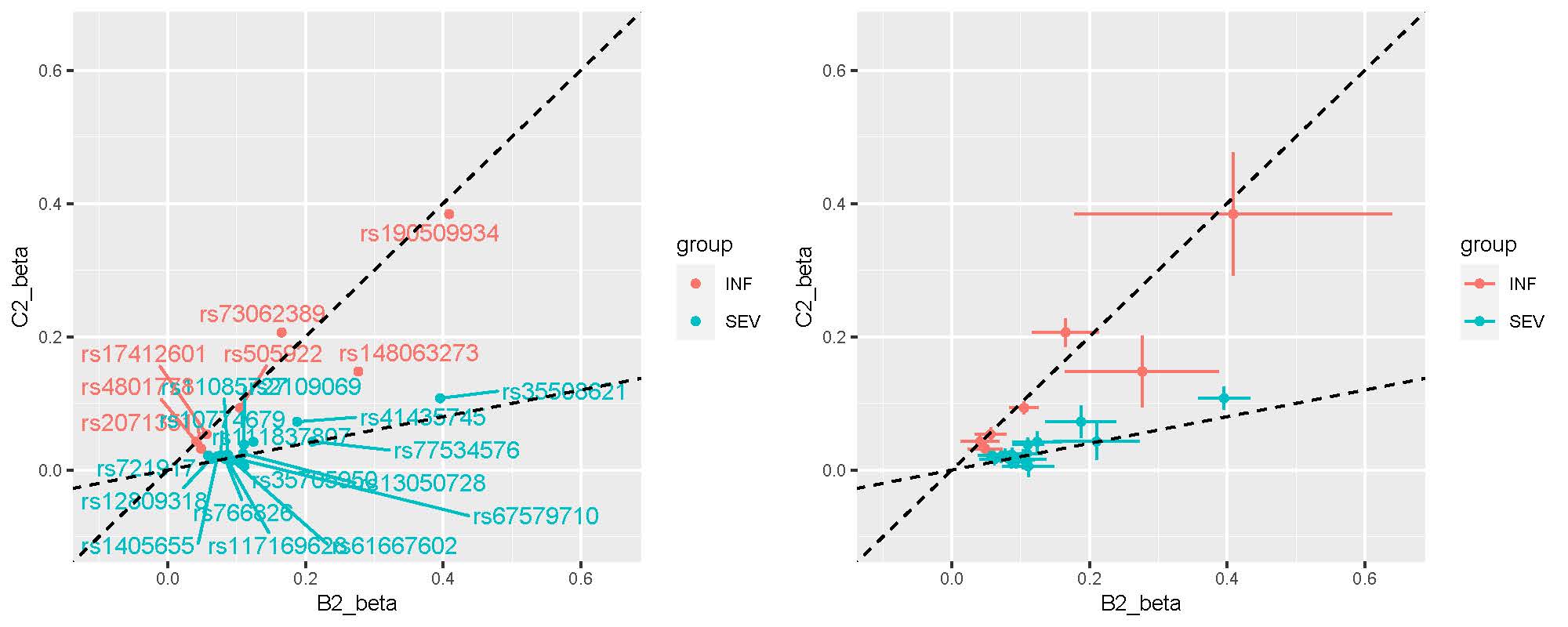

**Supplementary Figure 5**

**Projection of contributed samples from participating studies into the same PC space**. We asked participating studies to perform PC projection using the 1000 Genomes Project and Human Genome Diversity Project as a reference, with a common set of variants. For each panel (except for the reference), colored points correspond to contributed samples from each cohort, whereas gray points correspond to the 1000 Genomes reference samples. Color represents a genetic population that each cohort specified. Since 23andme, genomicsengland100kgp, and MVP only submitted PCA images, we overlaid their submitted transparent images using the same coordinates, instead of directly plotting them.

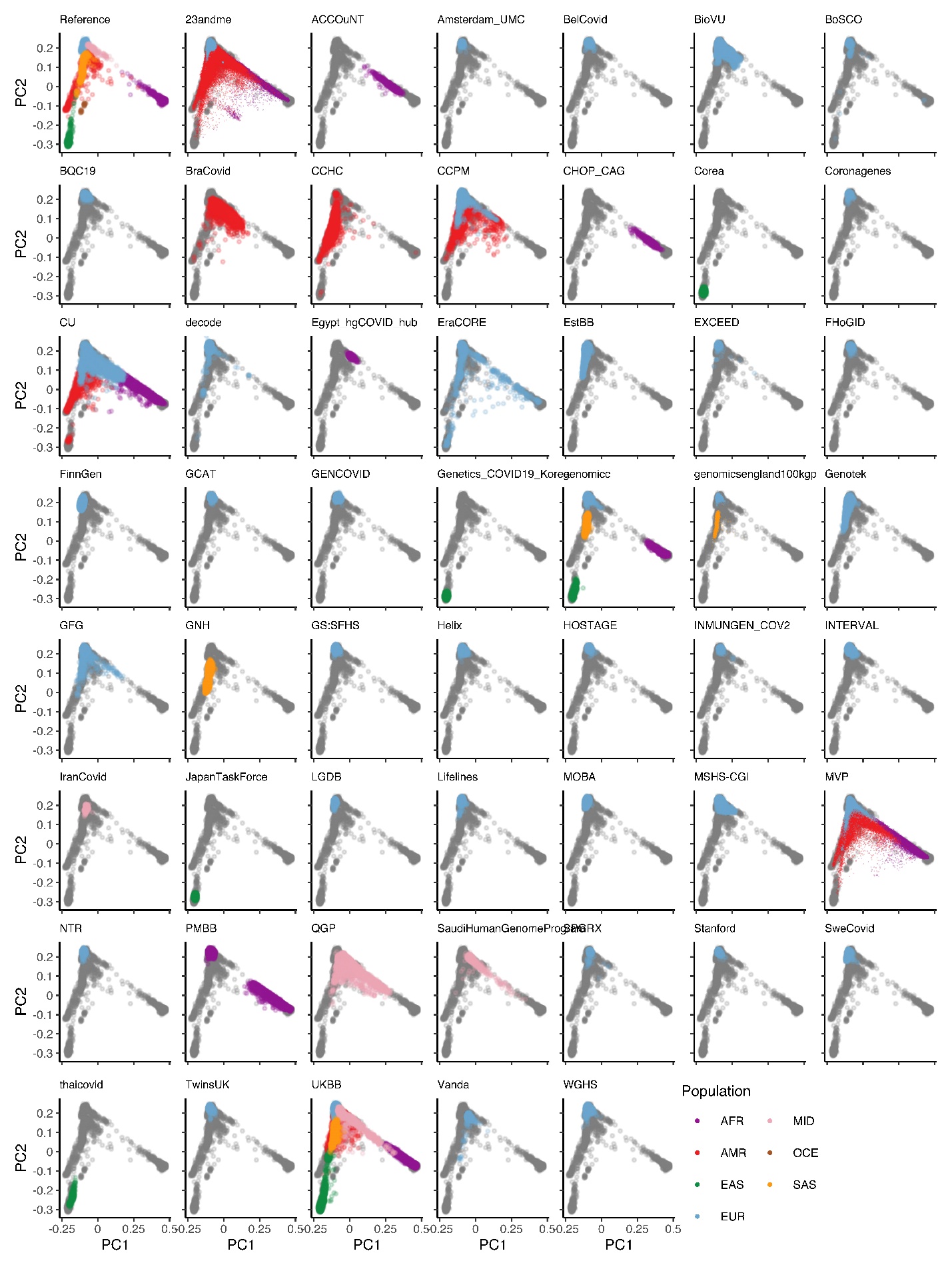

**Supplementary Figure 6**

**Forest plots for highly heterogeneous loci**. Forest plots for 6/23 loci that showed a significant heterogeneous effect across studies. For each of the loci, effect sizes and confidence intervals are reported for each contributing study. Studies are grouped by ancestry, with summary effects reported for each ancestry subgroup. The summary effect size across all studies is also reported in the bottom panel for each locus.

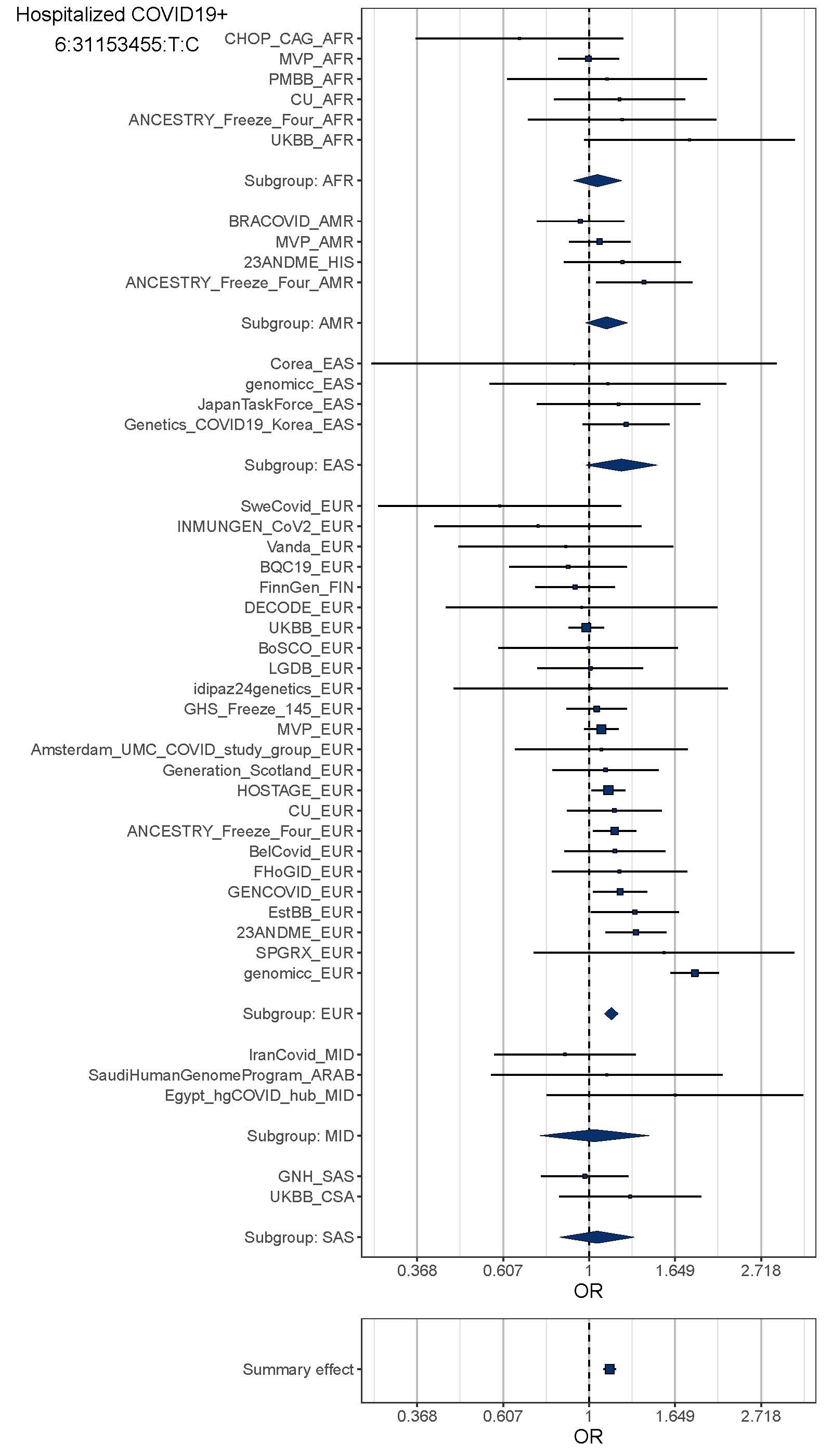

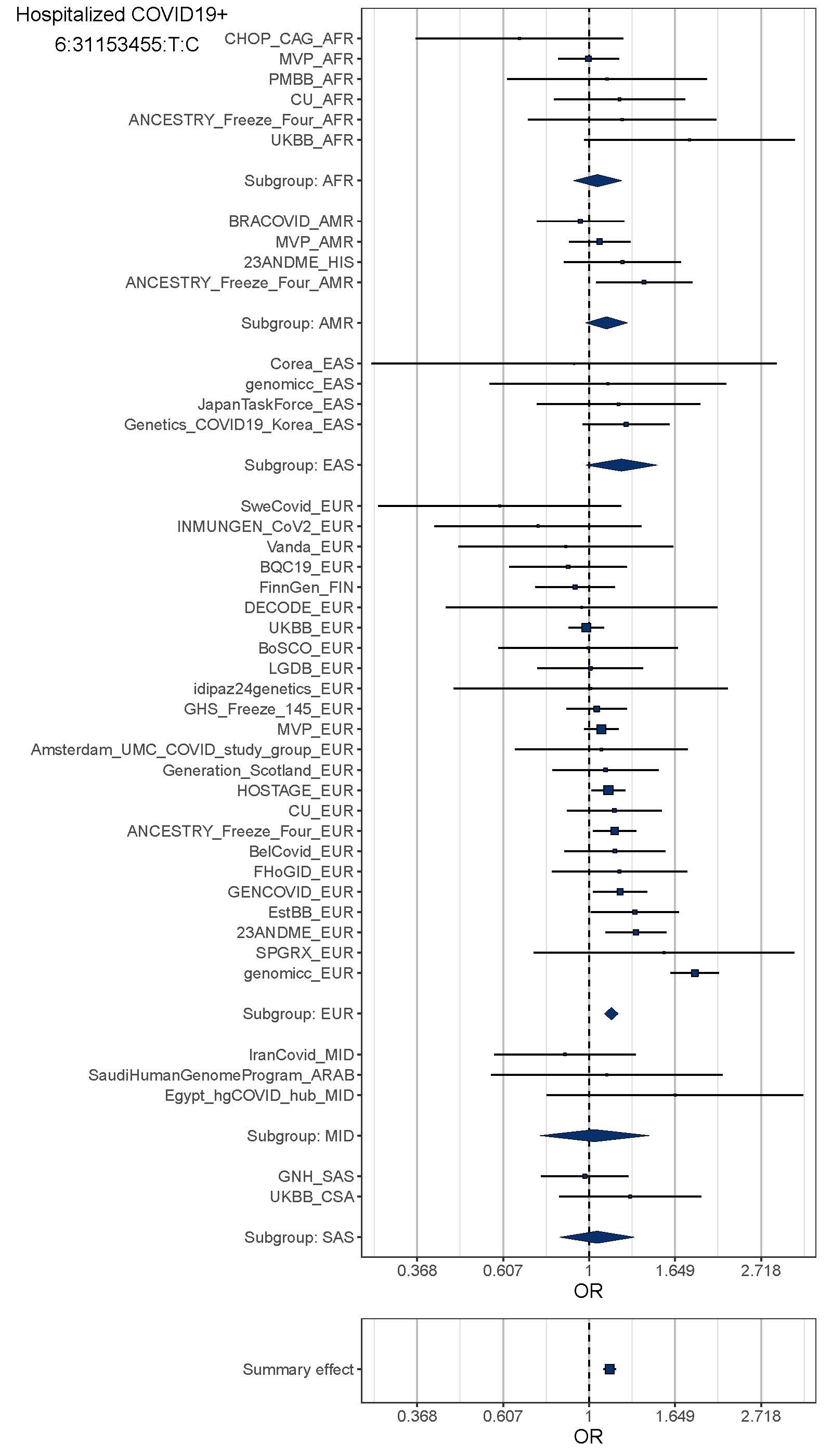

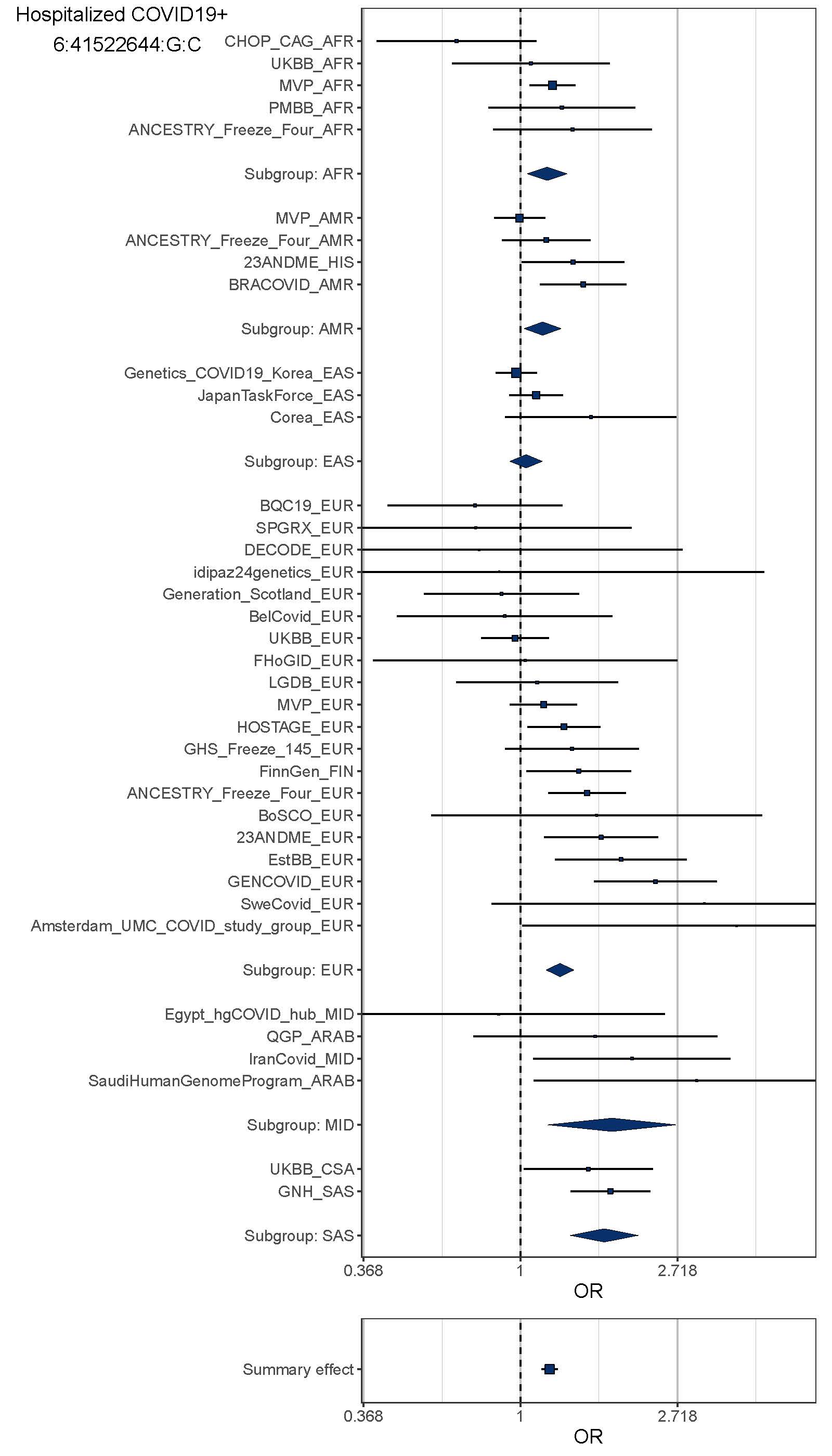

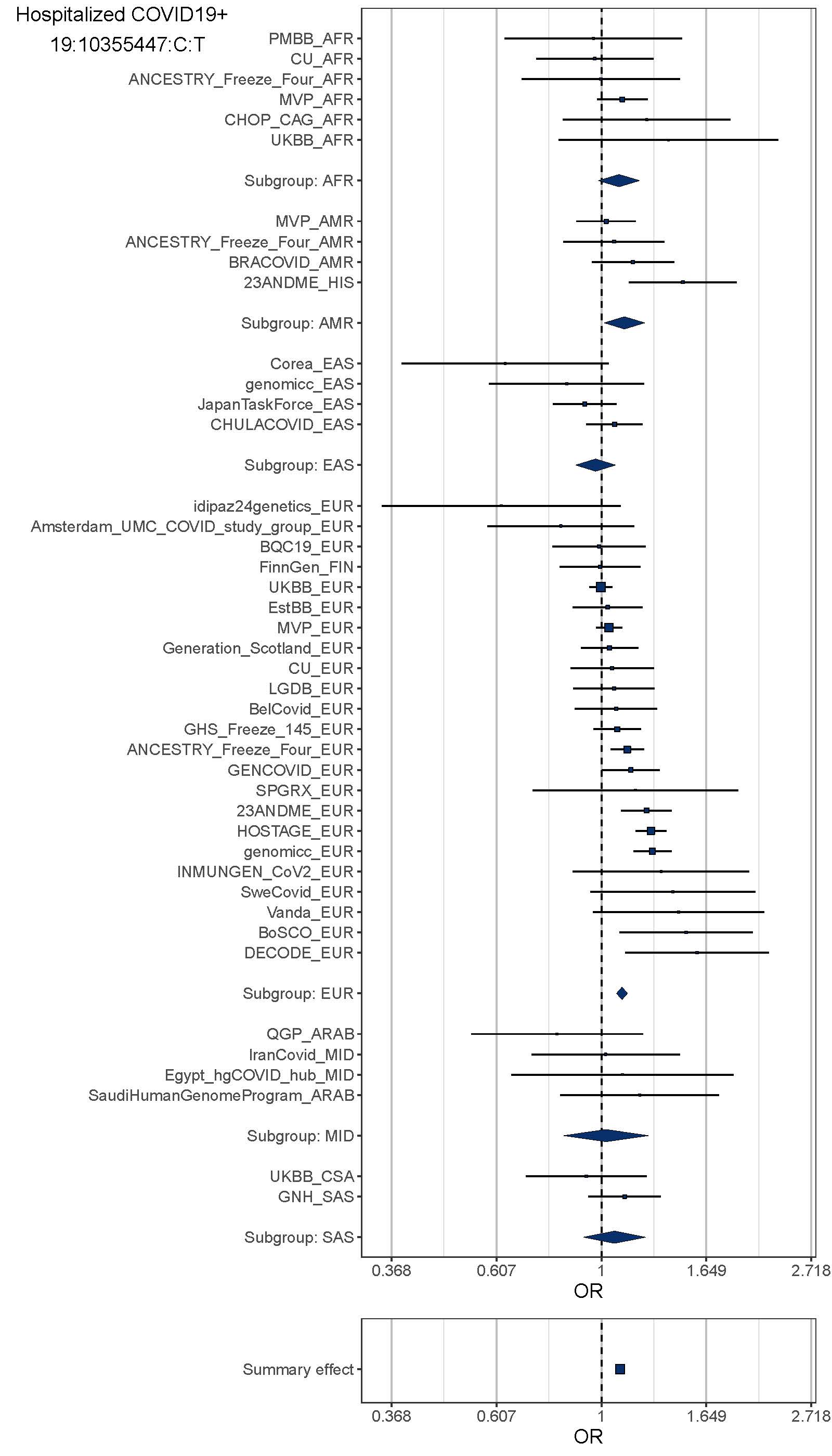

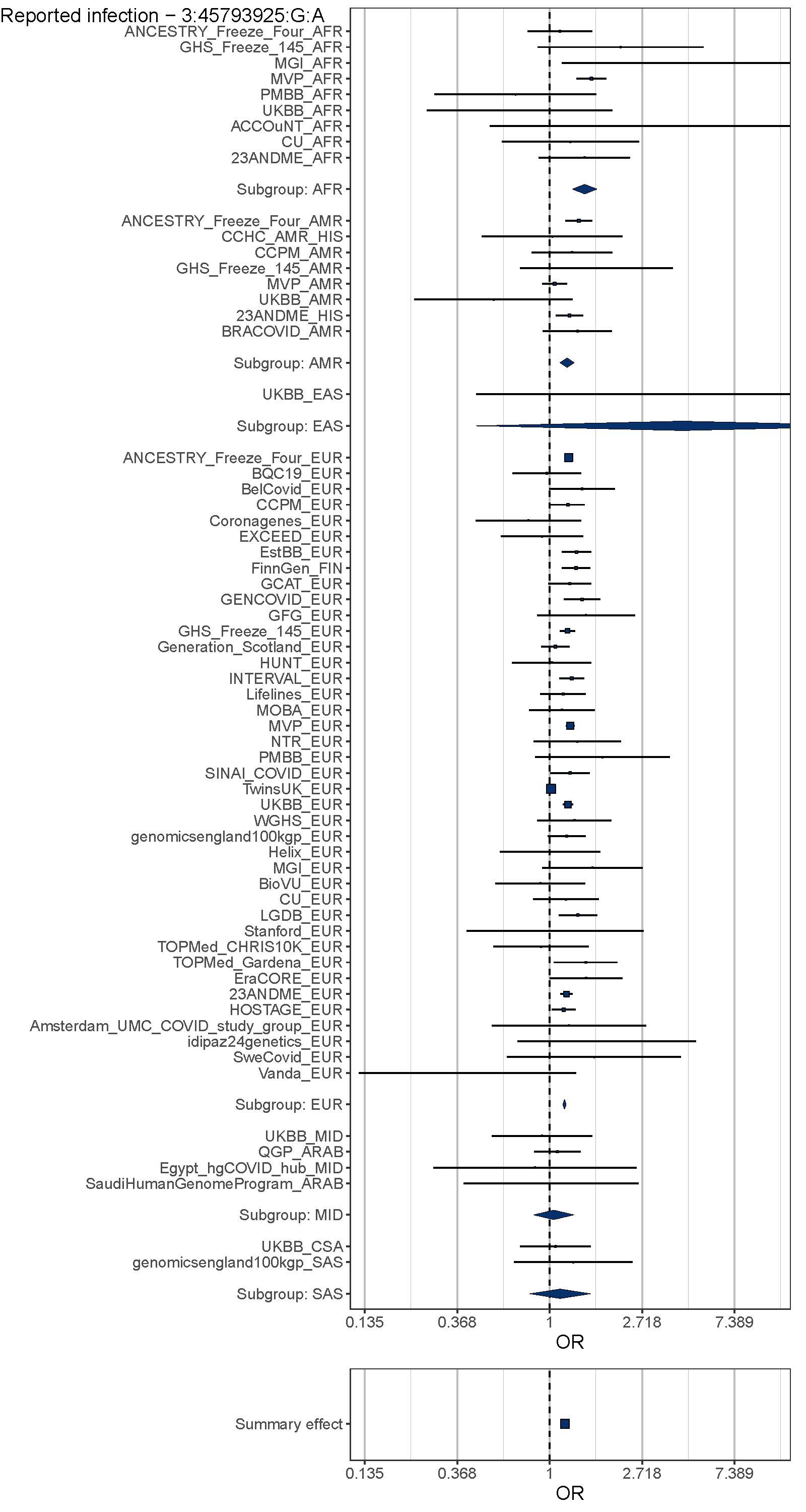

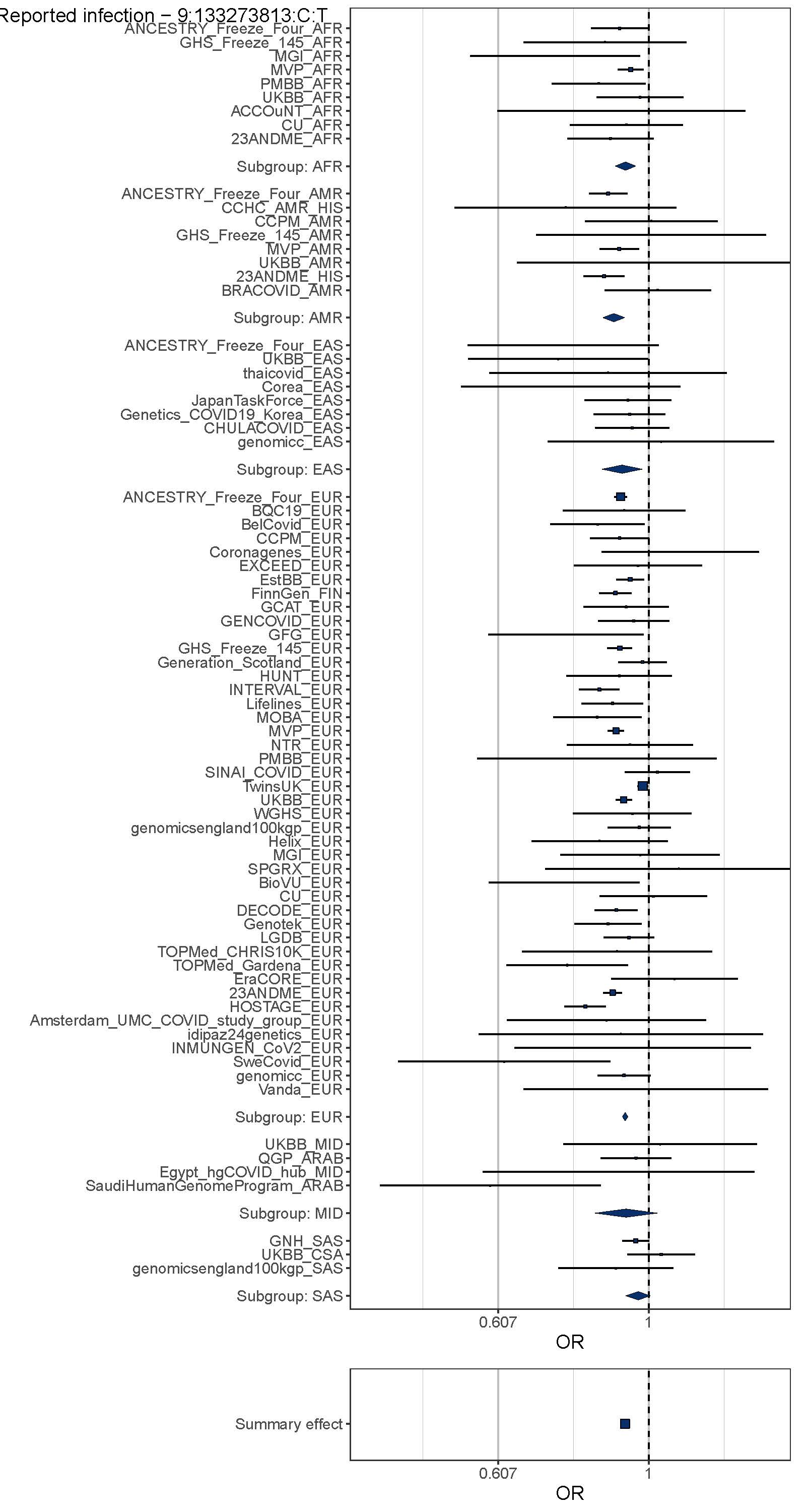

**Supplementary Figure 7**

**LocusZoom plots to visualize the meta-analysis results at the loci passing genome-wide significance**. For each genome-wide significant locus in three meta-analyses: meta-analysis of critical illness, hospitalization, and reported infection, we showed 1) a Manhattan plot of each locus where a color represents a weighted-average r2 value (see COVID-19 Host Genetics Initiative, 2021) to a lead variant (unadjusted P-values from the two-tailed inverse variance weighted meta-analysis); 2) r2 values to a lead variant across gnomAD v2 populations, i.e., African/African-American (AFR), Latino/Admixed American (AMR), Ashkenazi Jewish (ASJ), East Asian (EAS), Estonian (EST), Finnish (FIN), Non-Finish Europeans (NFE), North-Western Europeans (NWE), and Southern Europeans (SEU); 3) genes at a locus; and 4) genes prioritized by each gene prioritization metric where a size of circles represents a rank in each metric. Note that the COVID-19 lead variants were chosen across all the meta-analyses (Supplementary Table 2) and were not necessarily a variant with the most significant P-value from each inverse variance weighted meta-analysis.

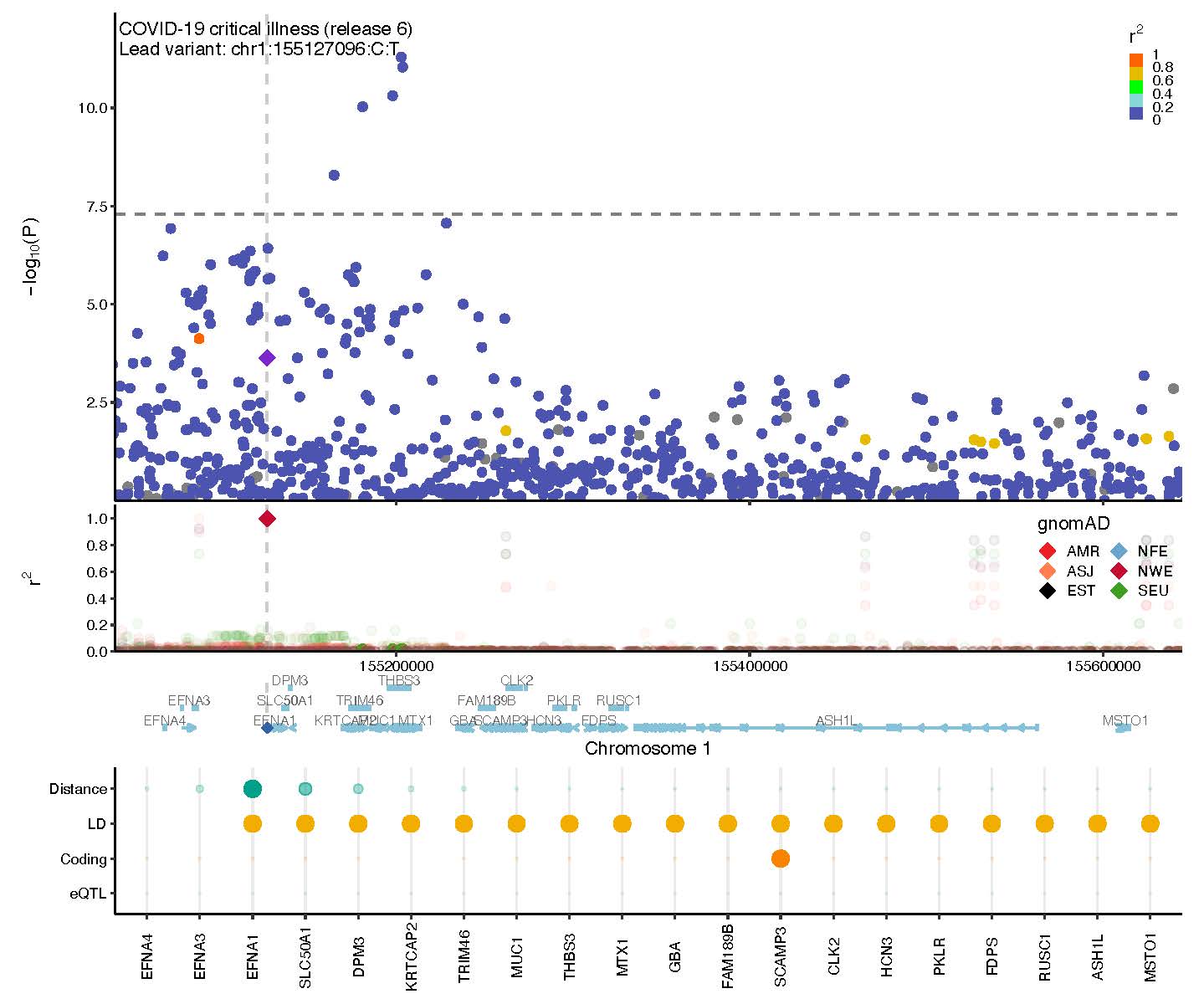

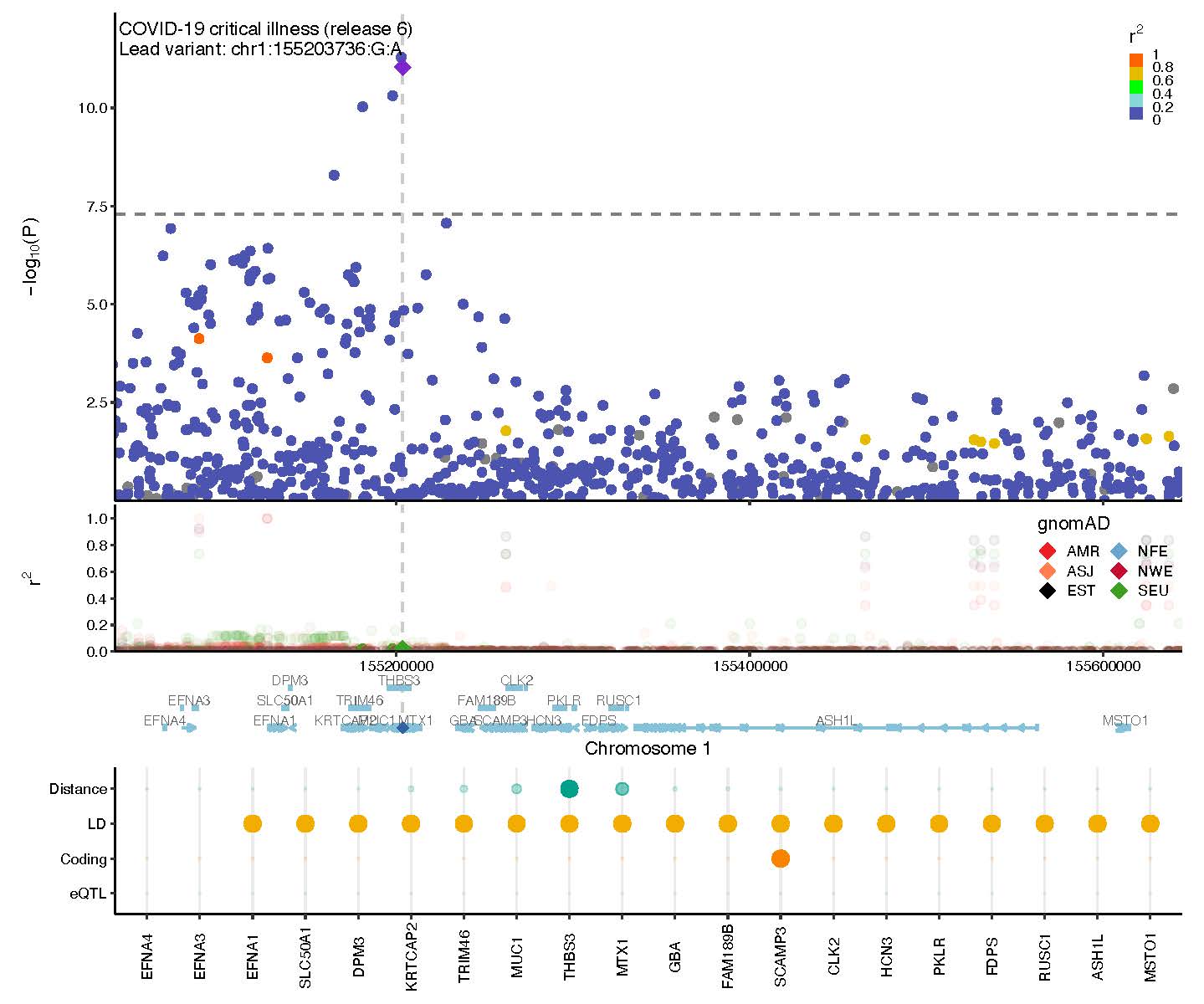

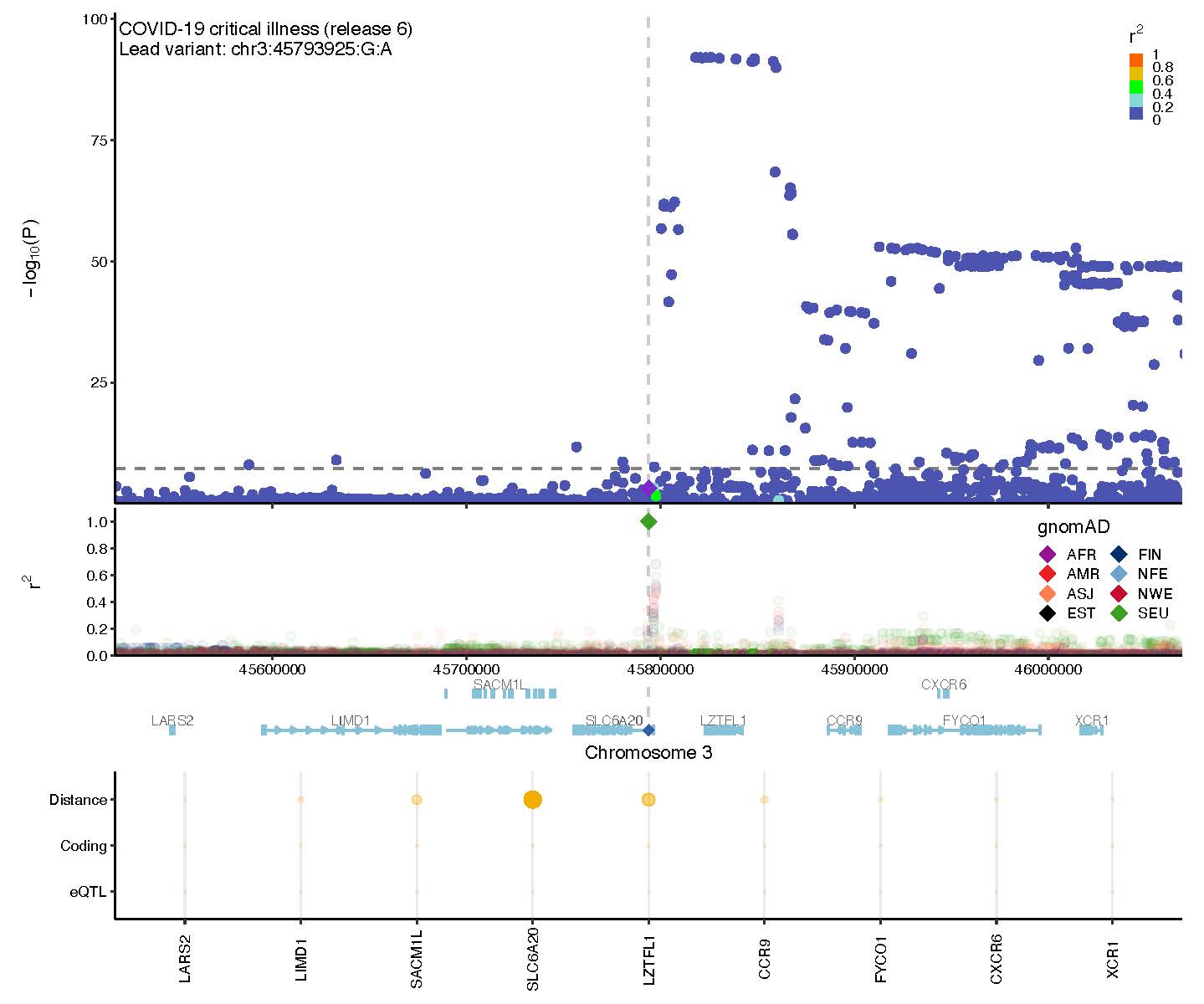

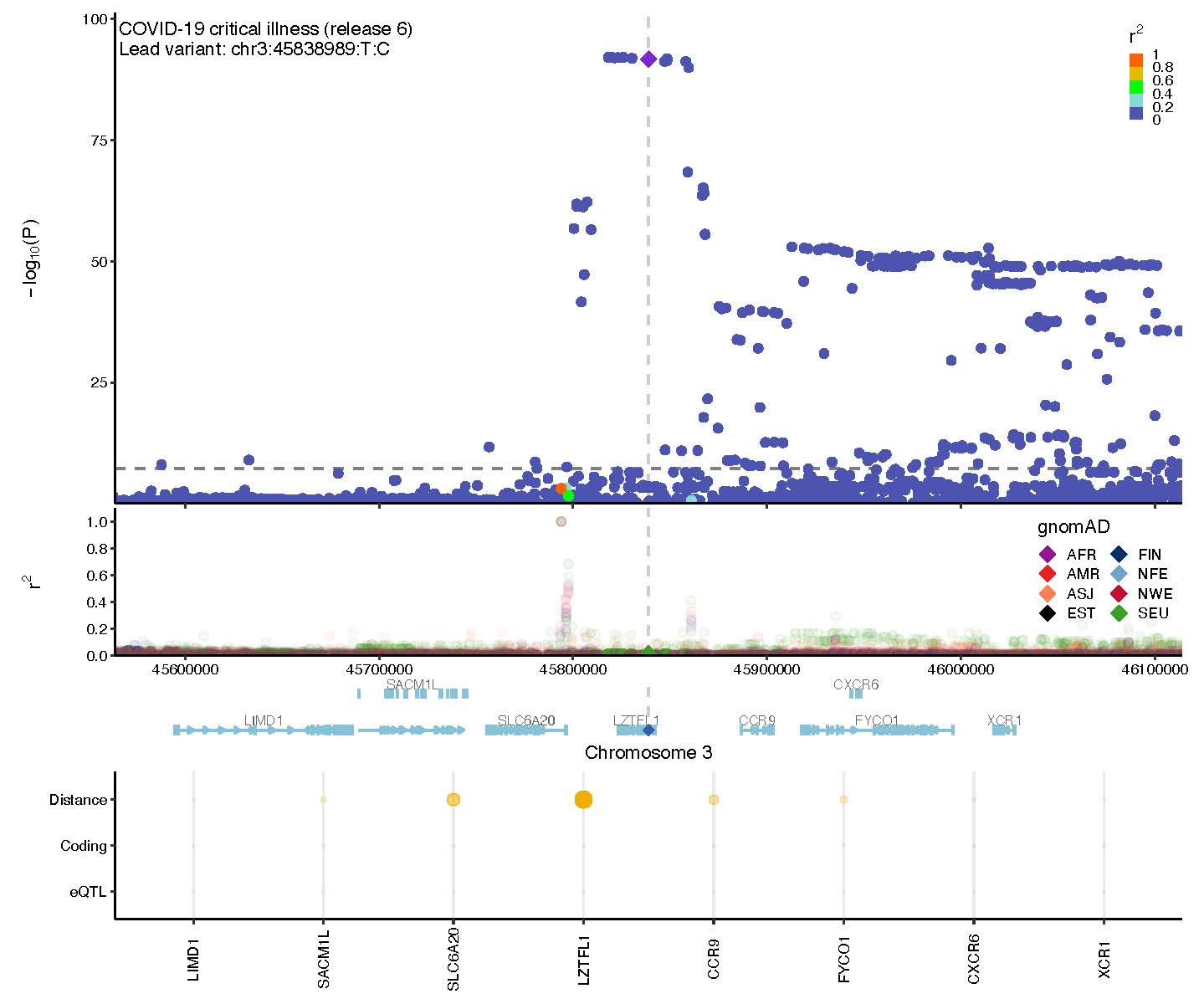

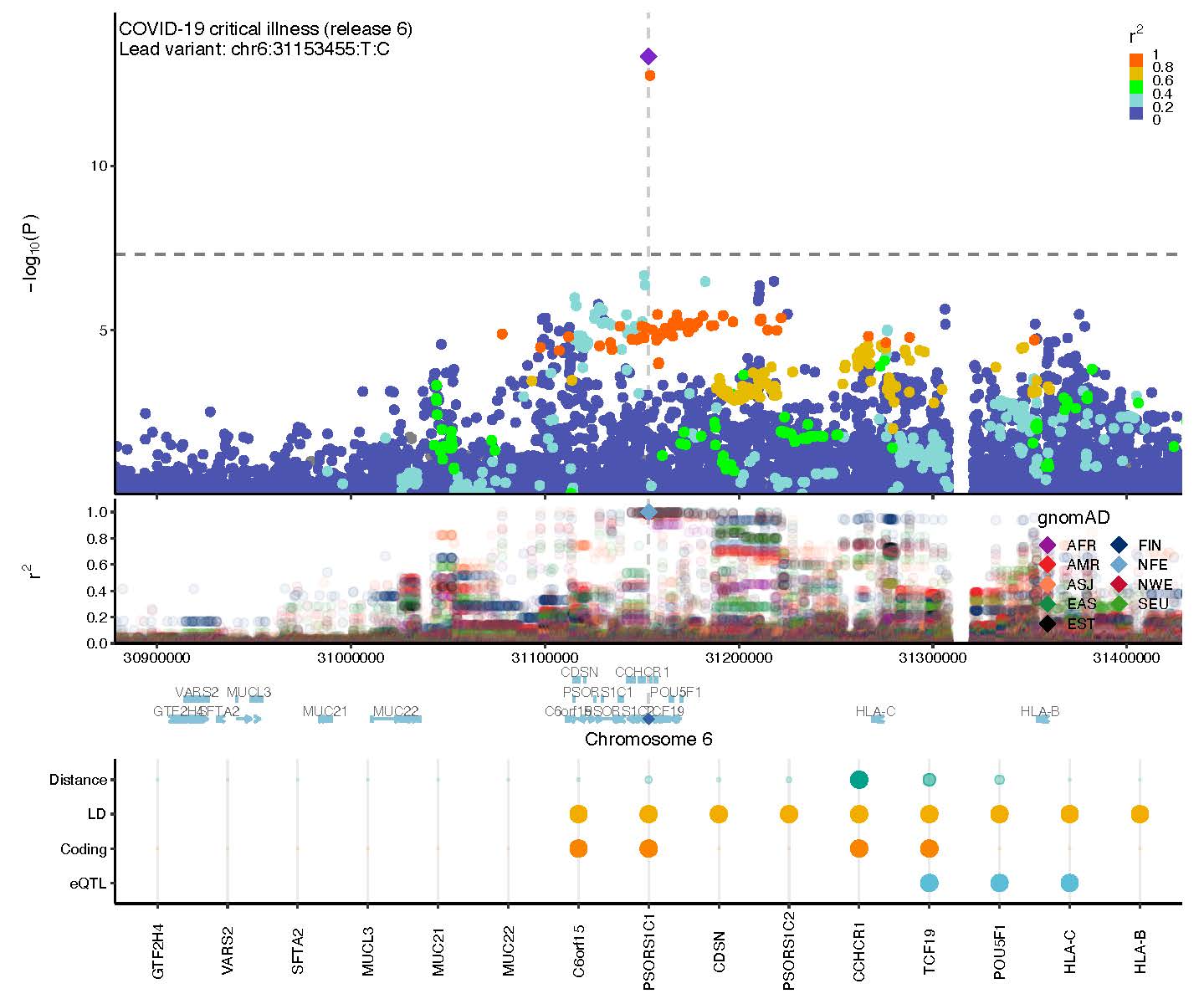

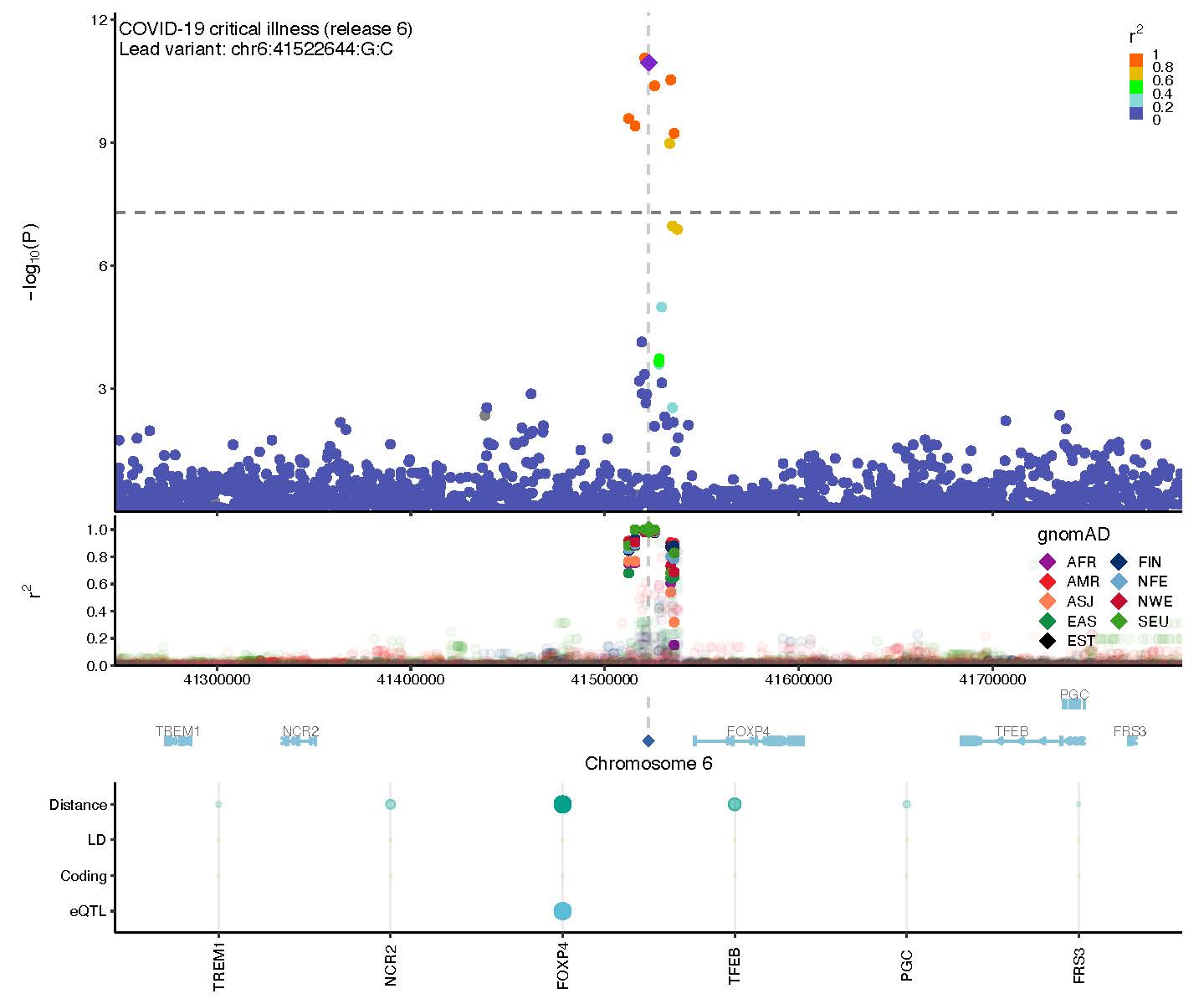

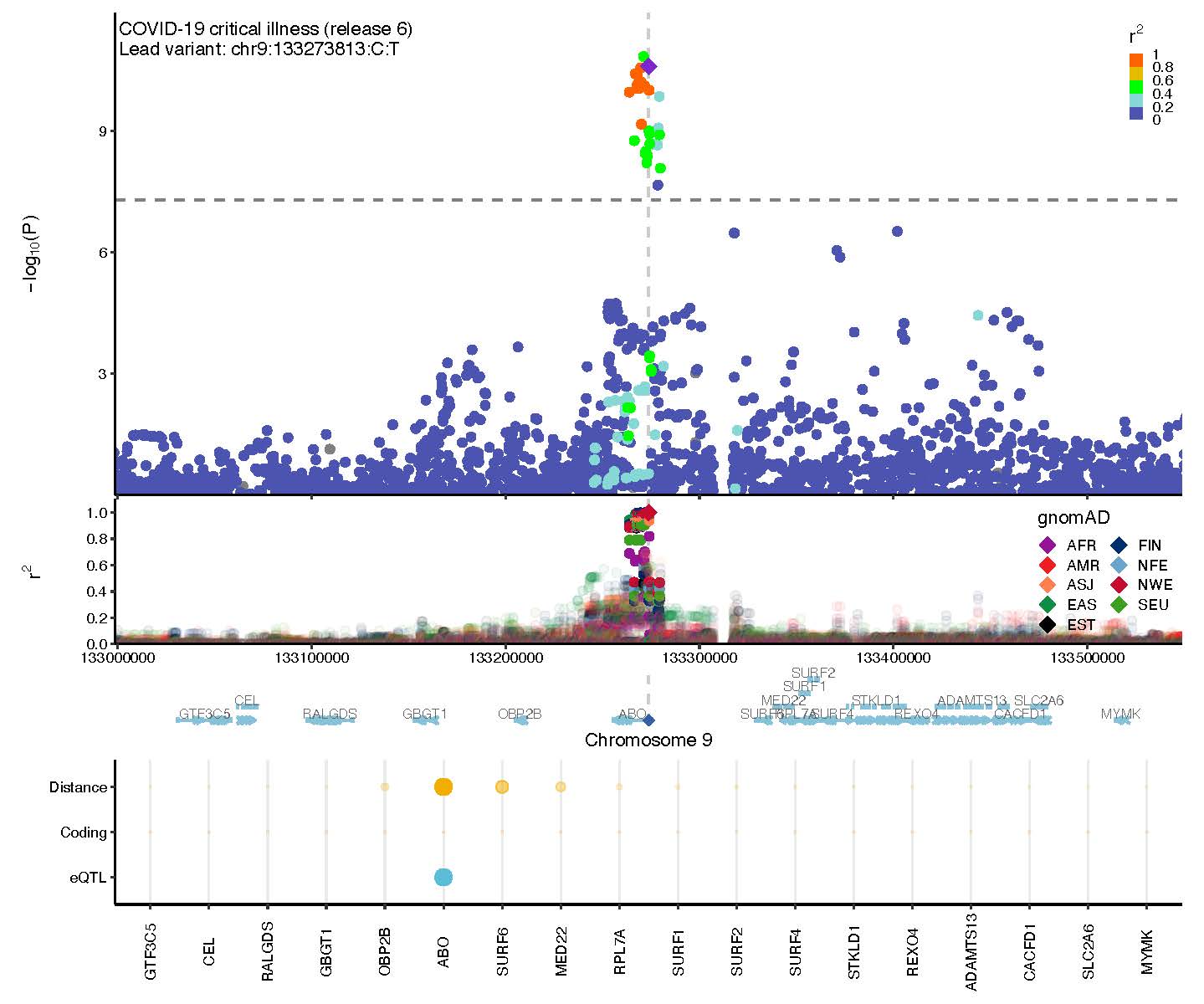

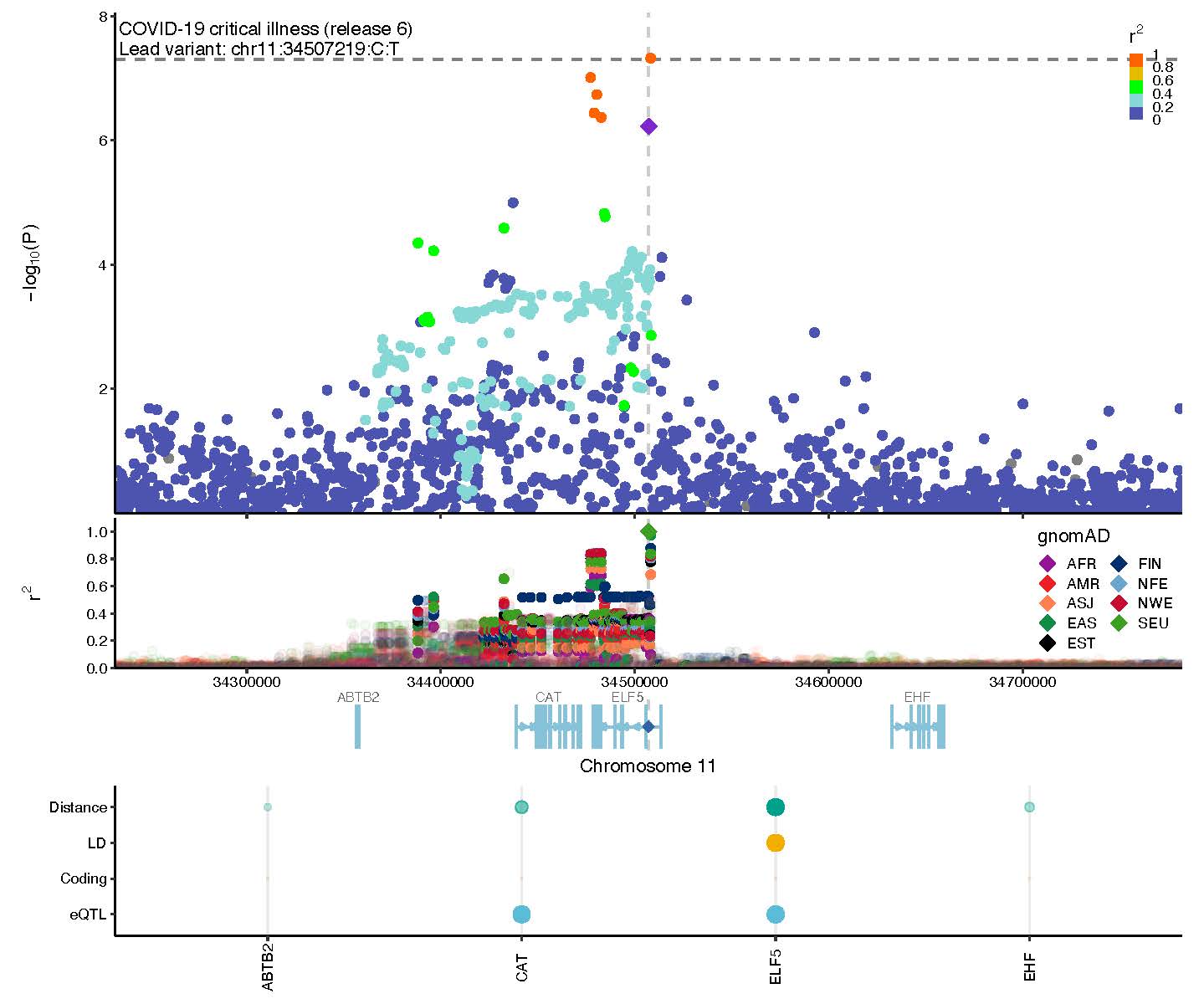

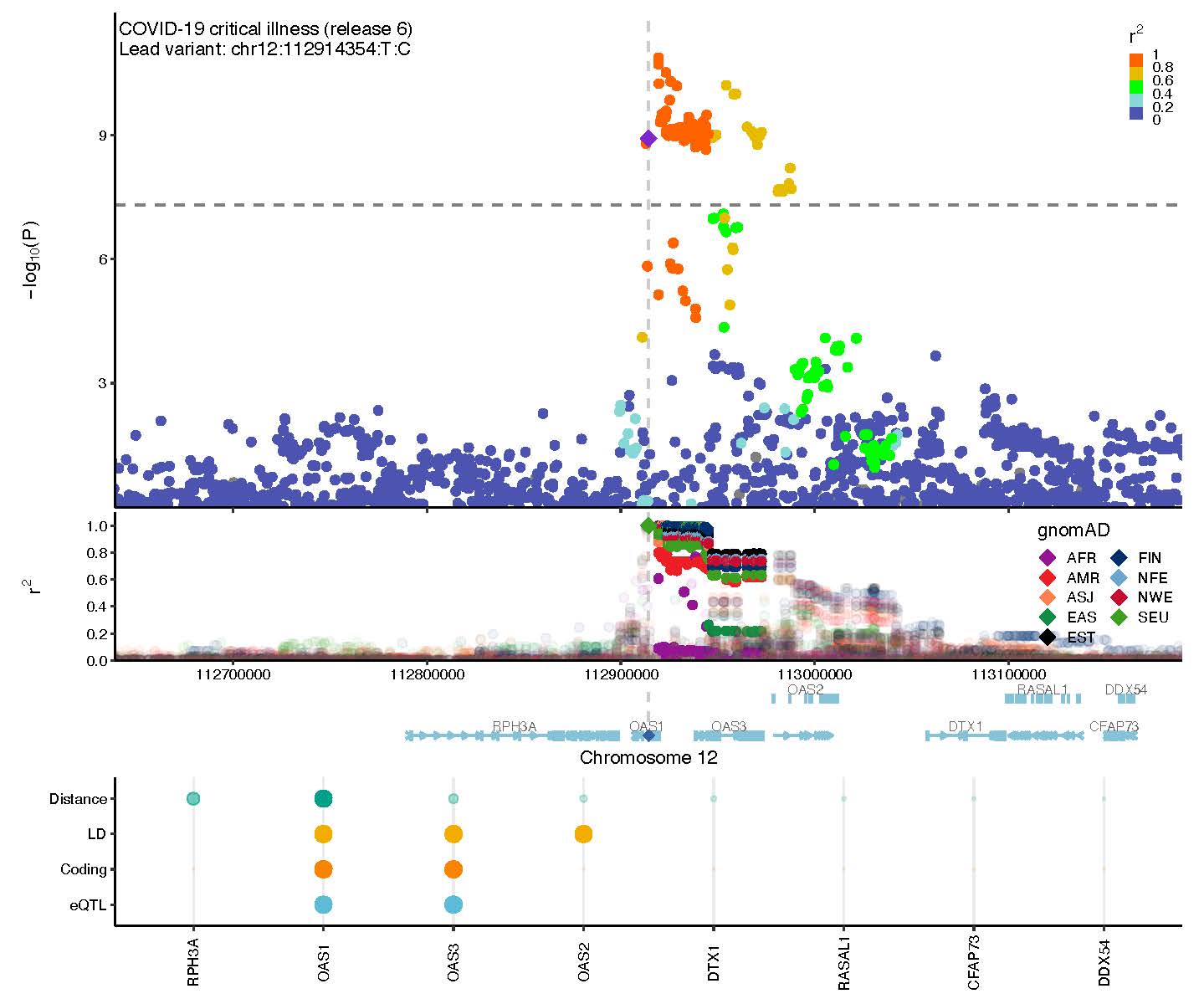

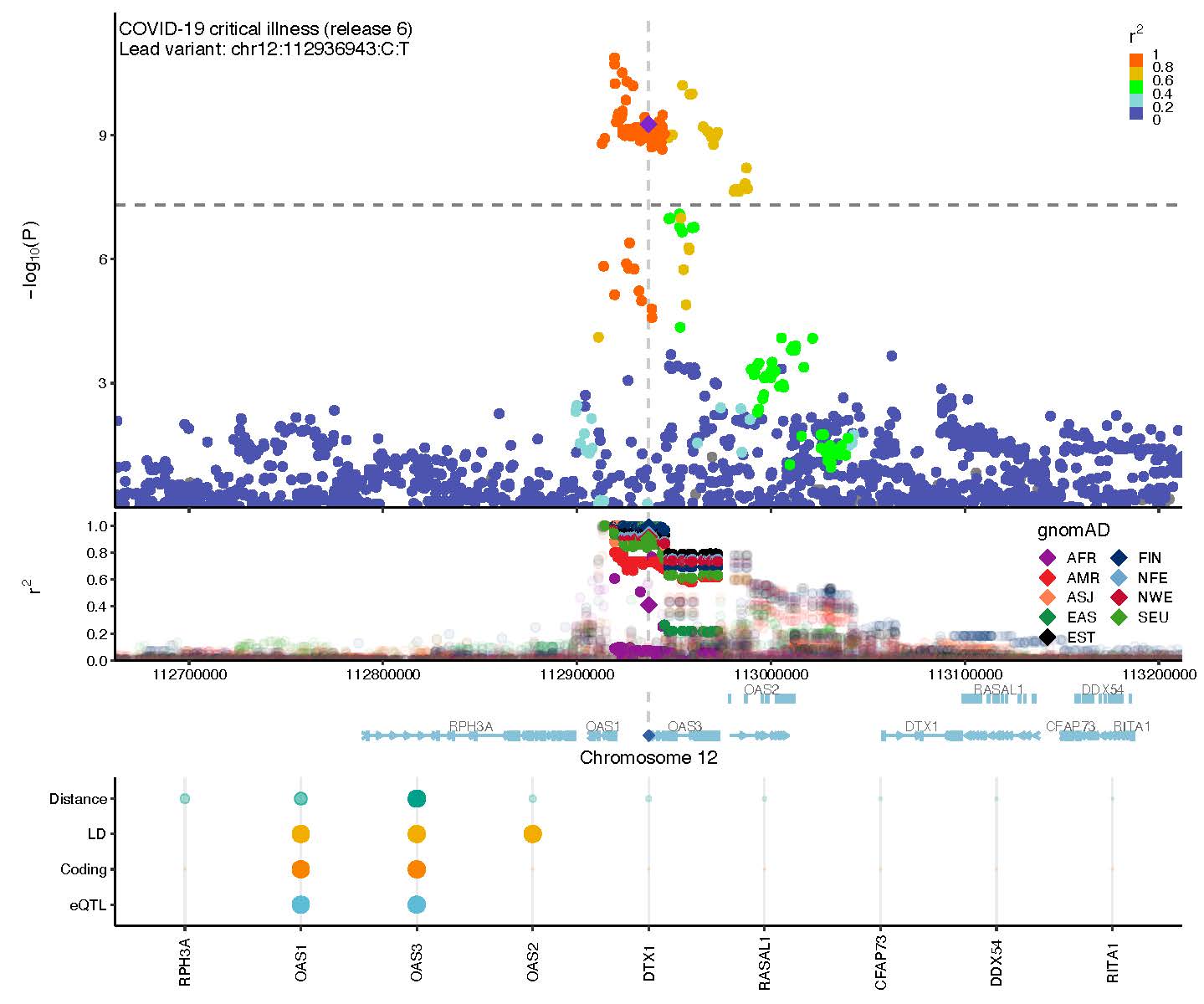

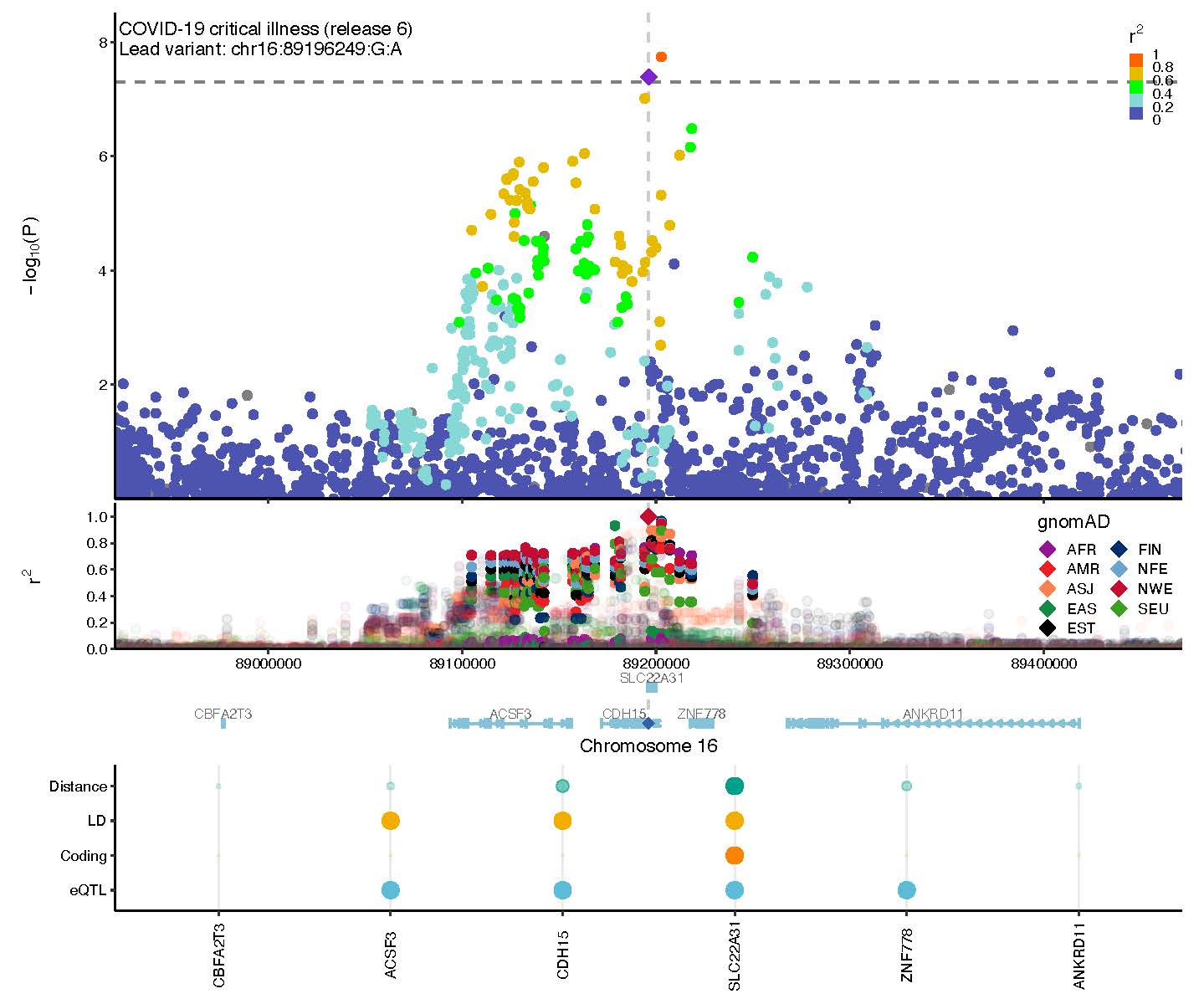

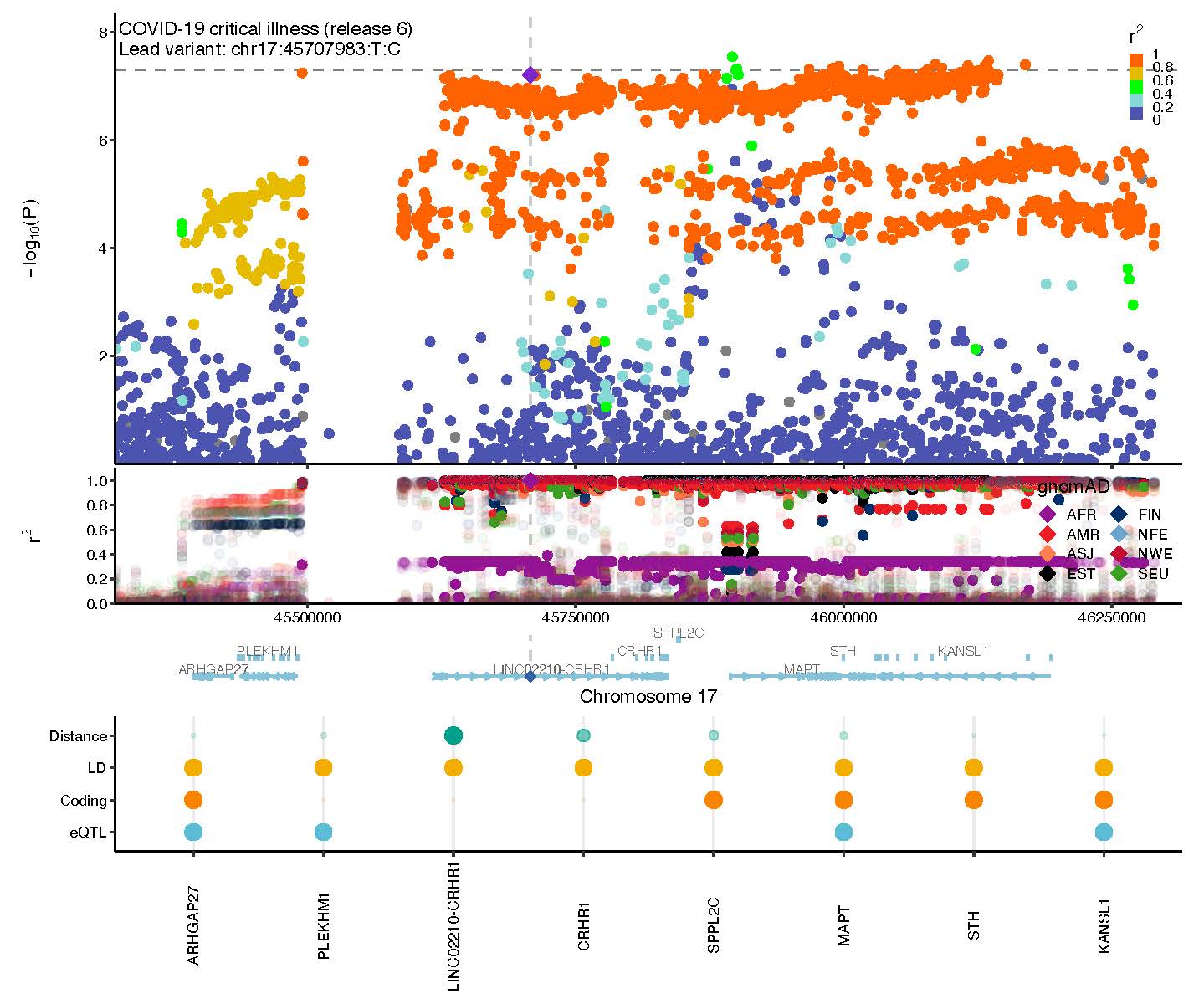

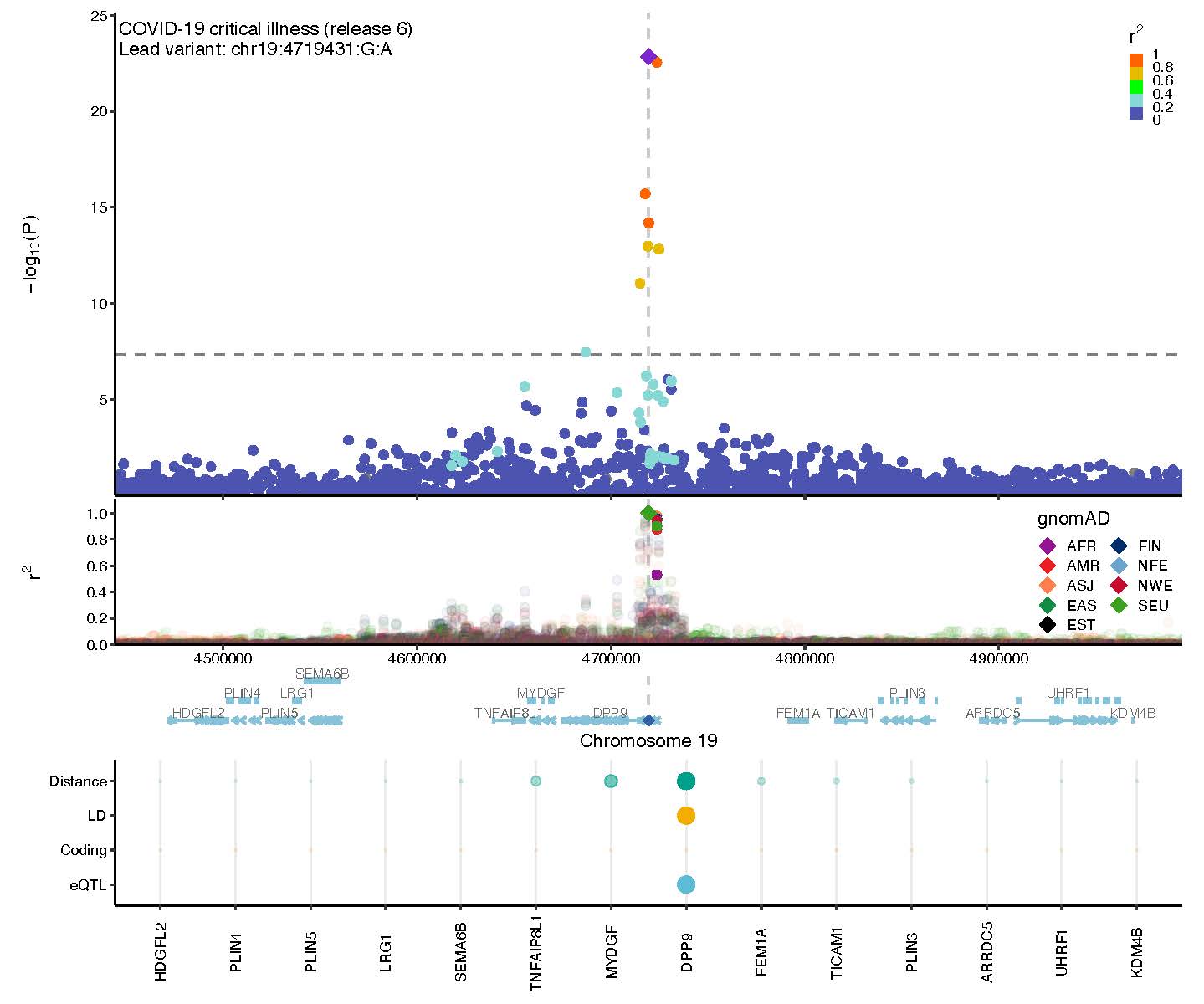

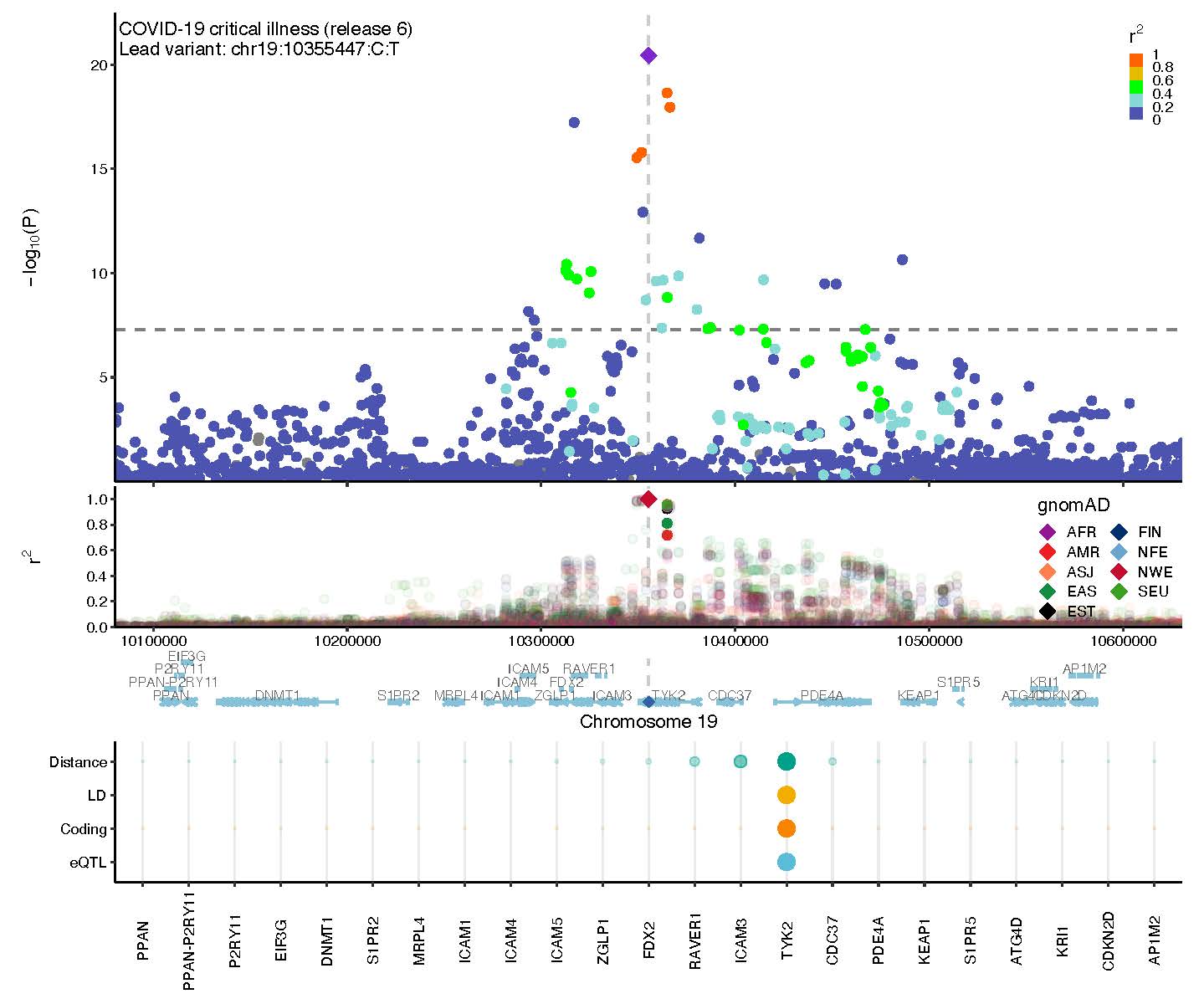

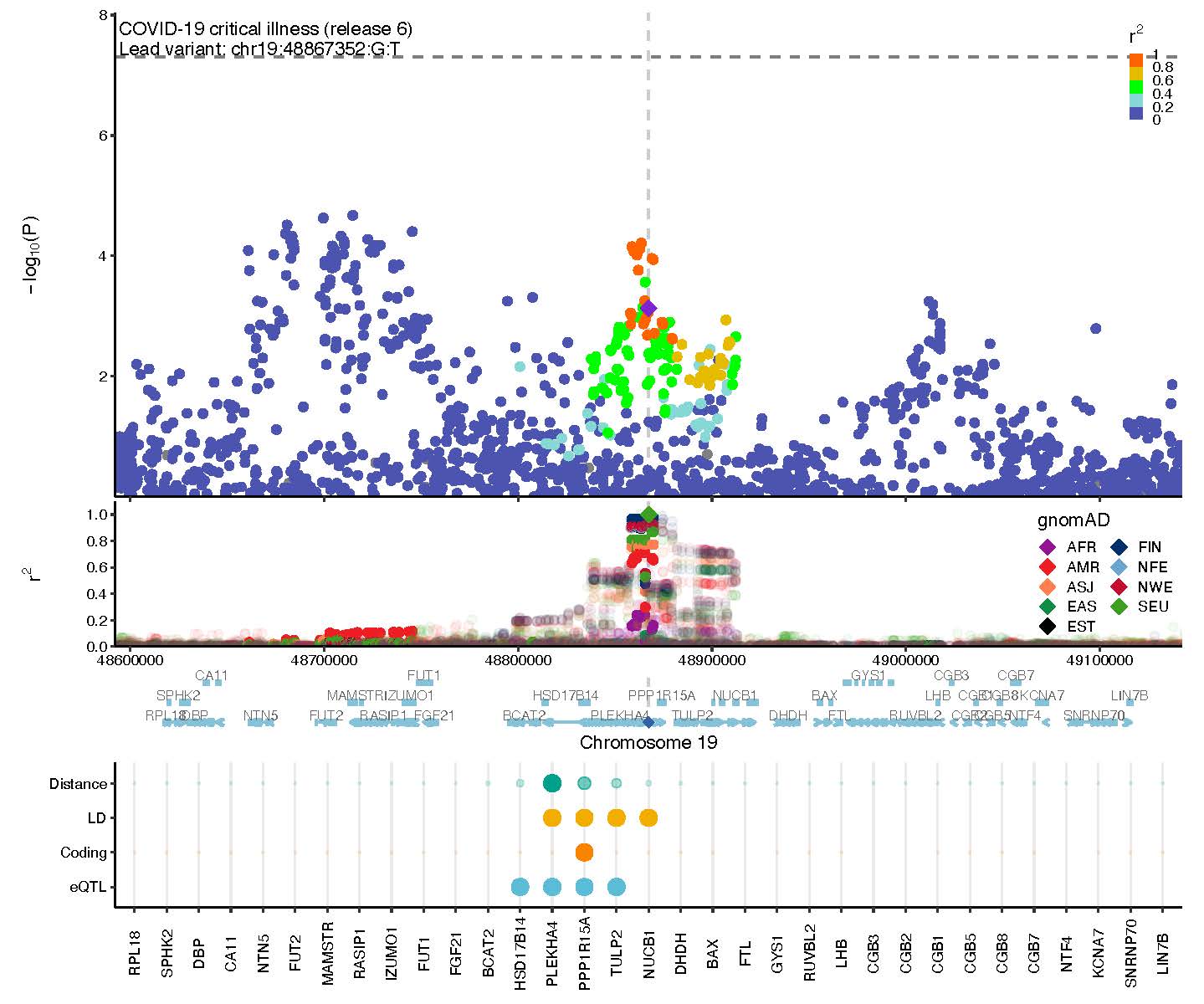

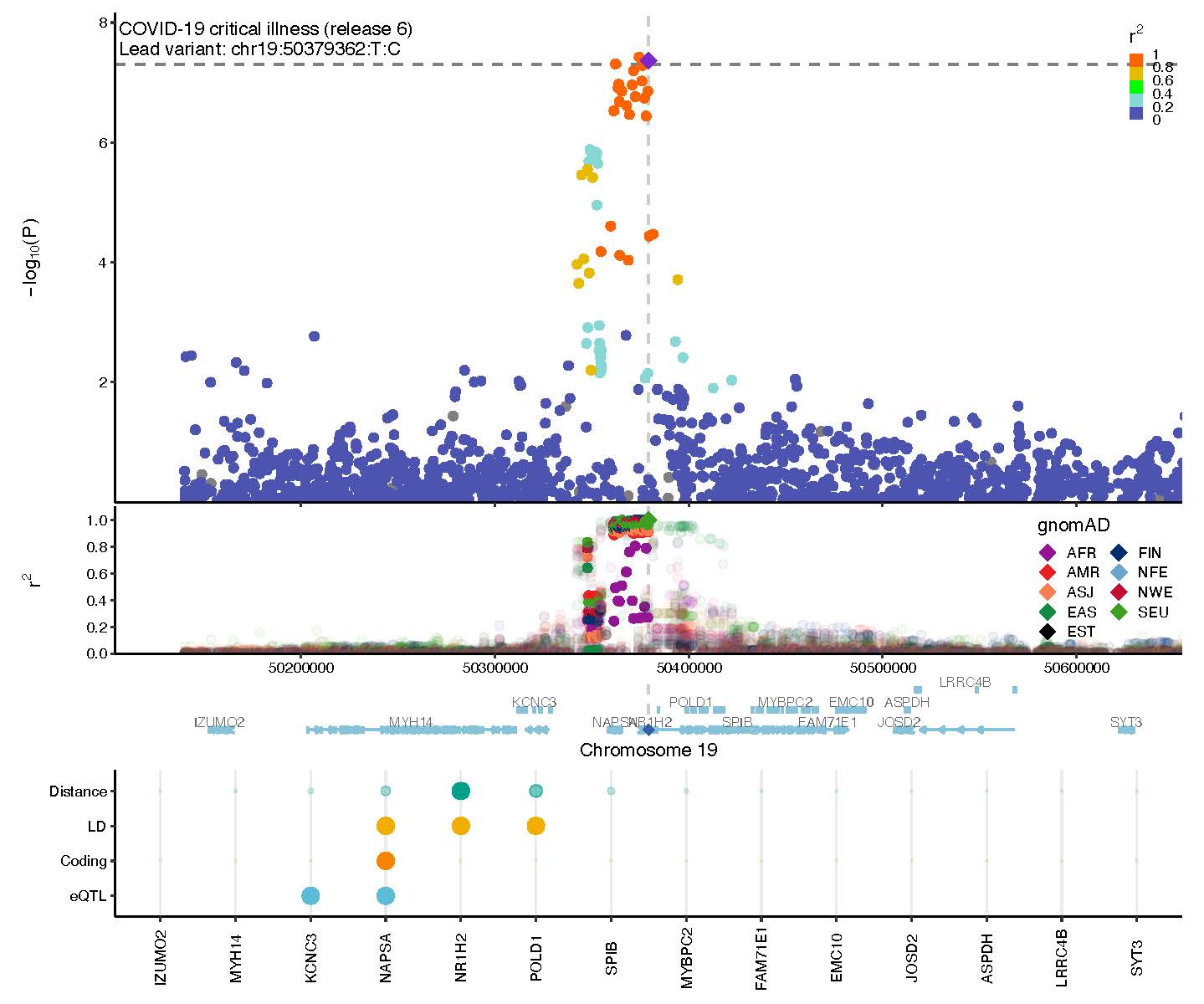

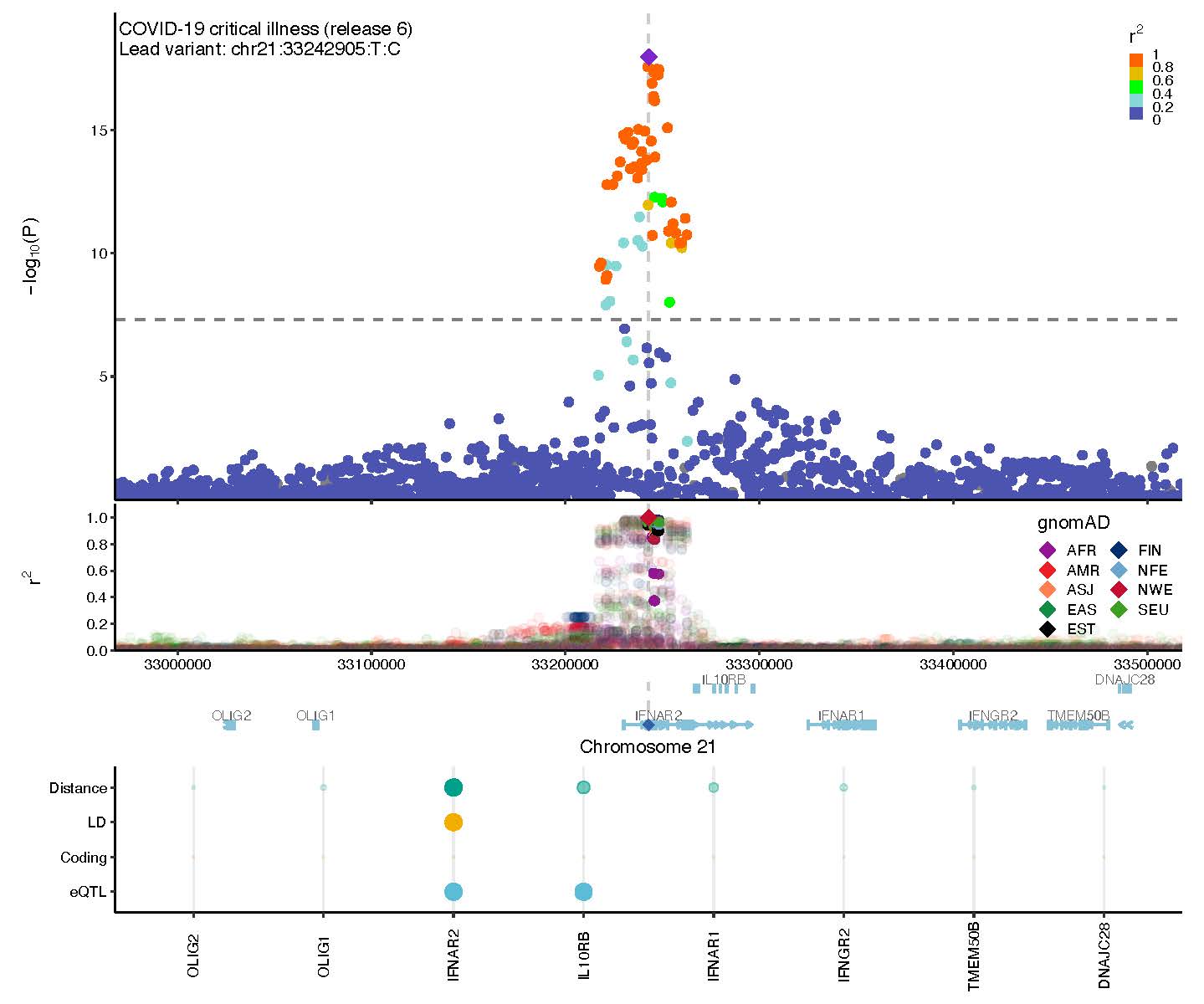

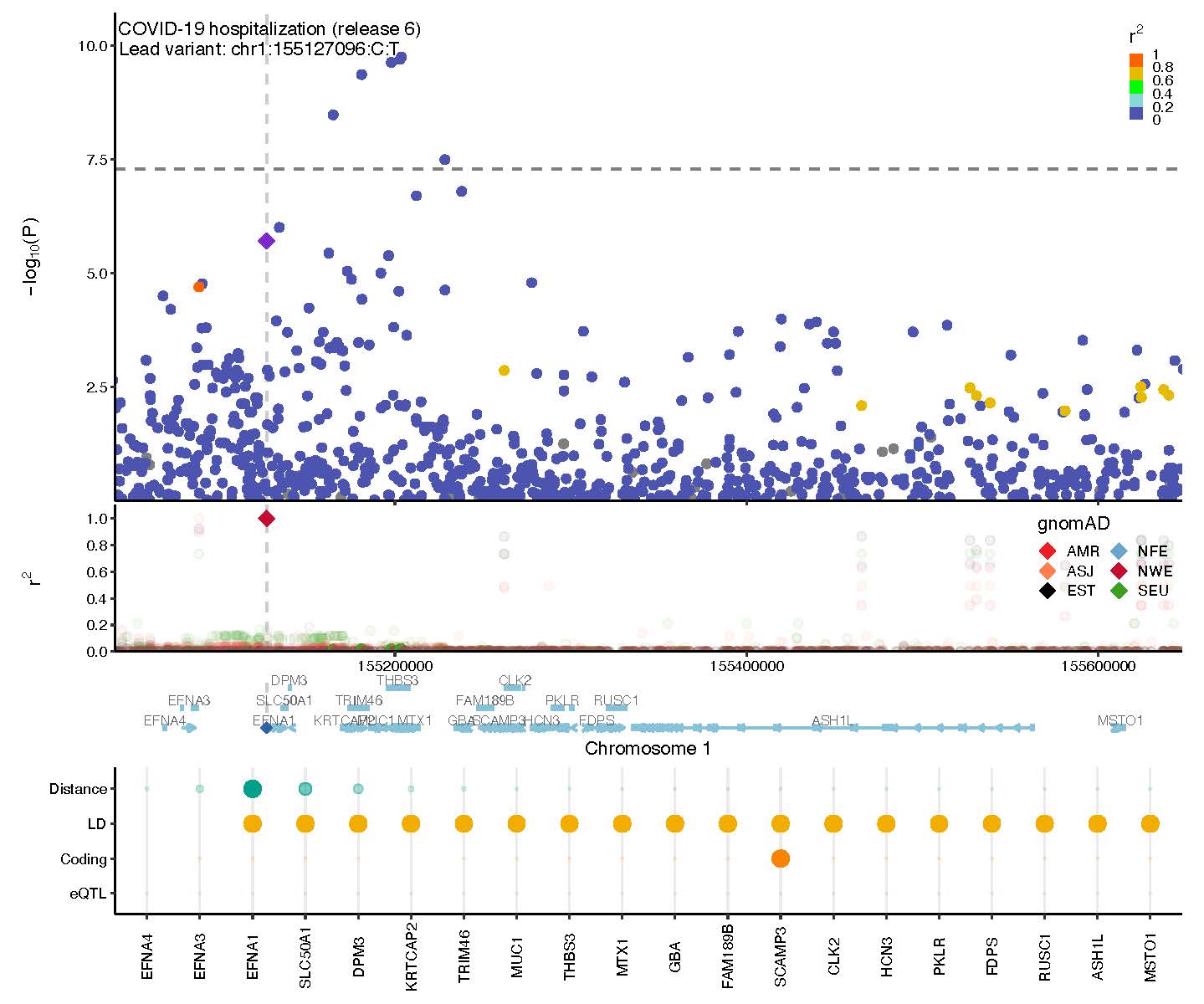

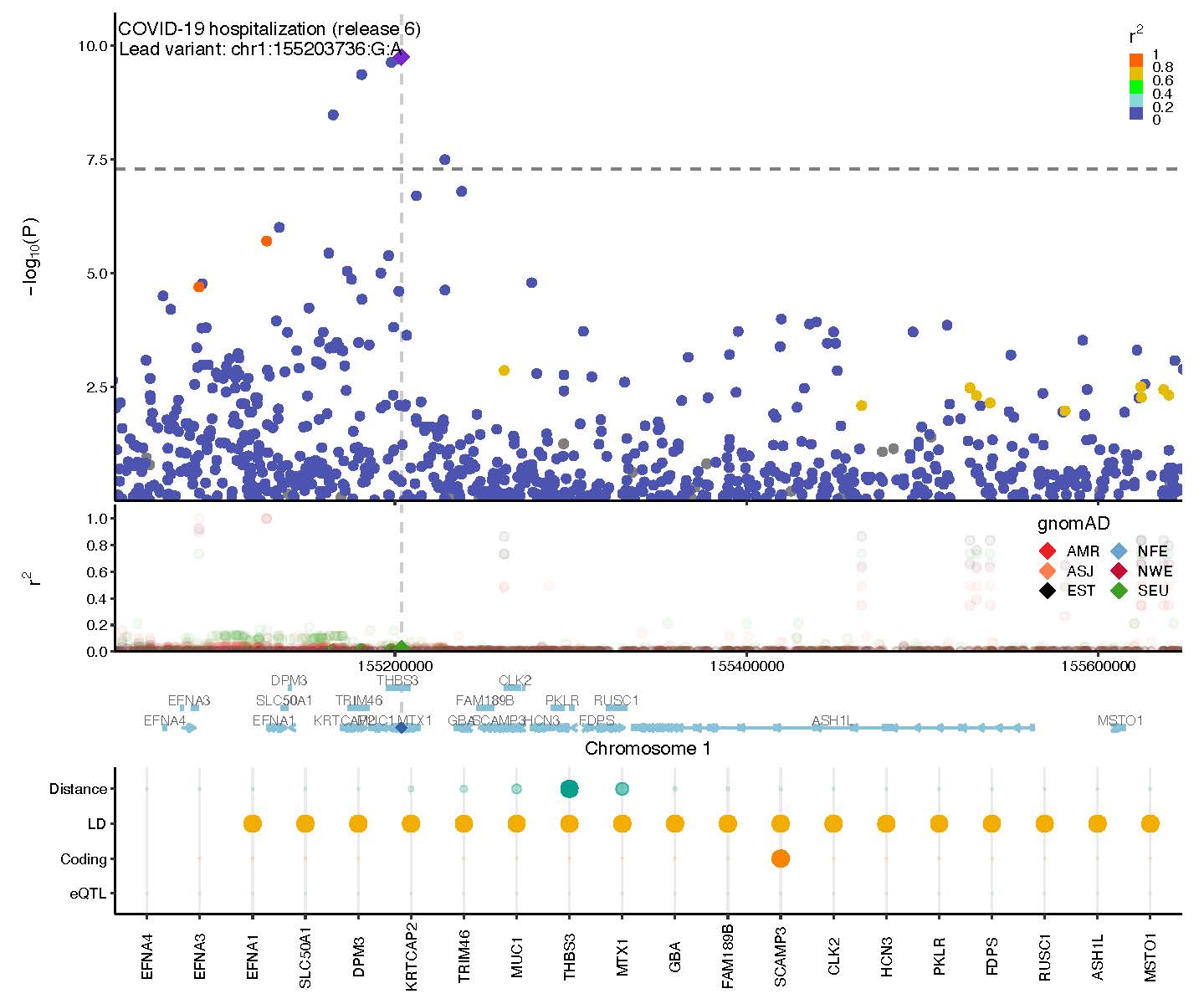

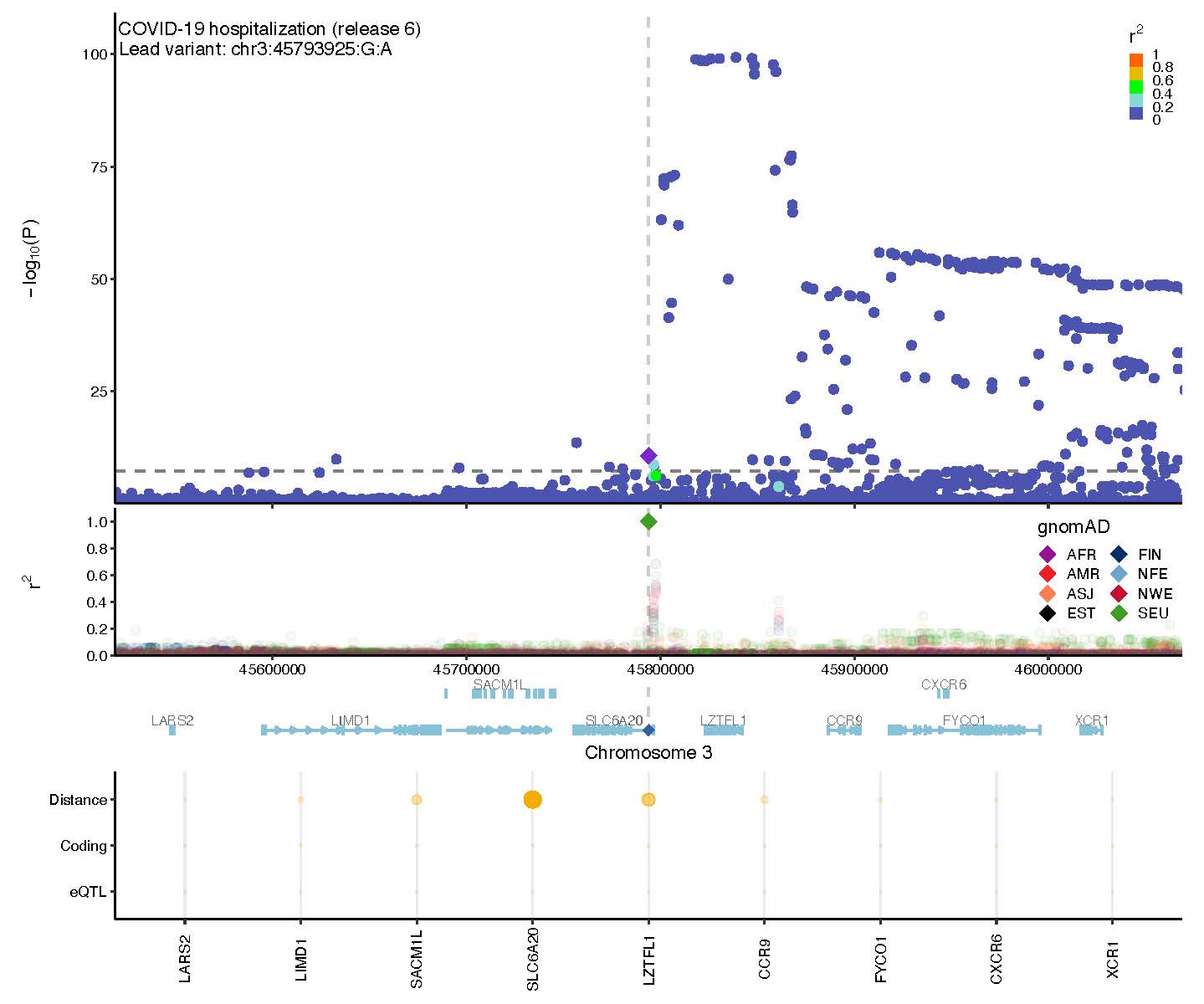

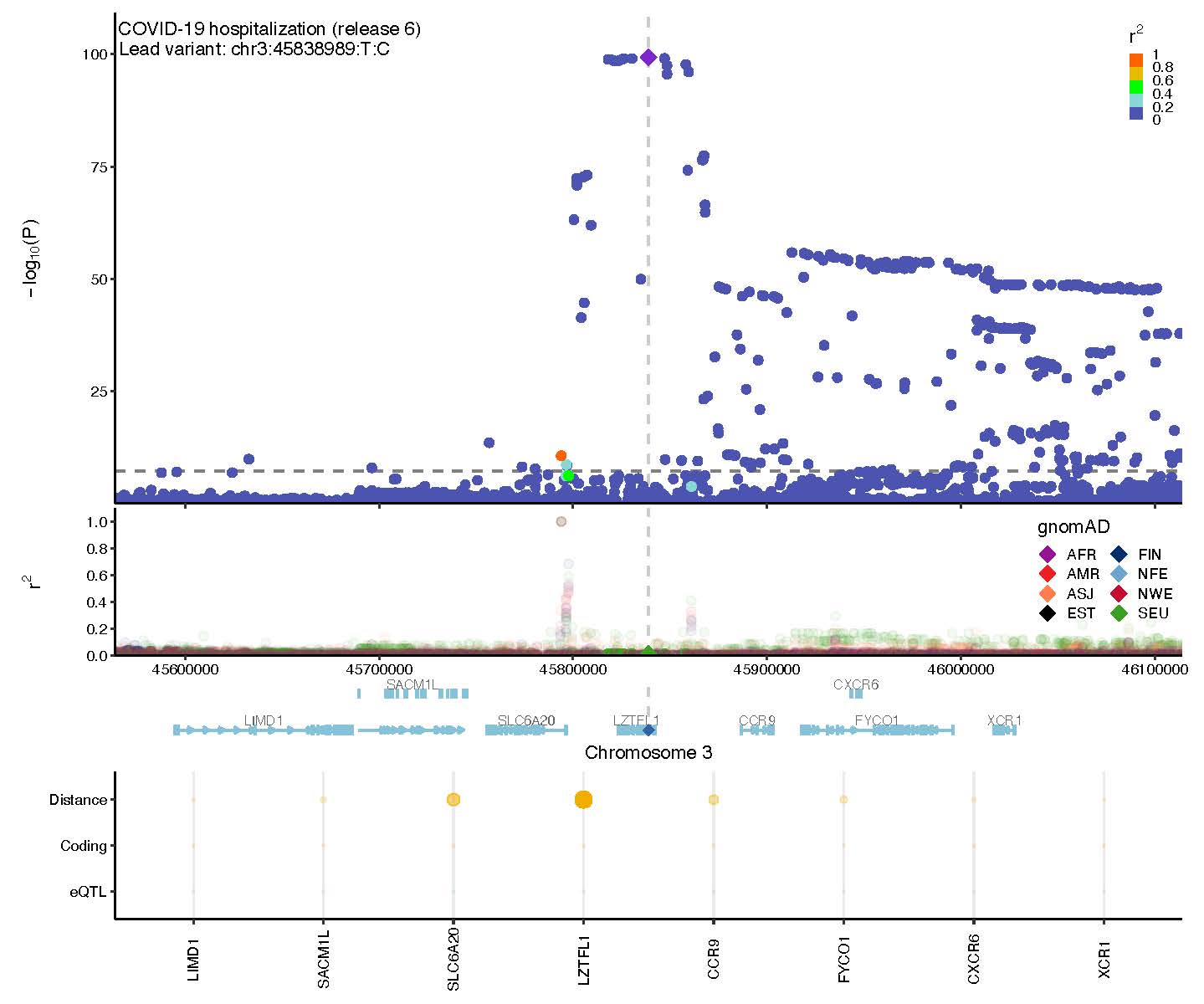

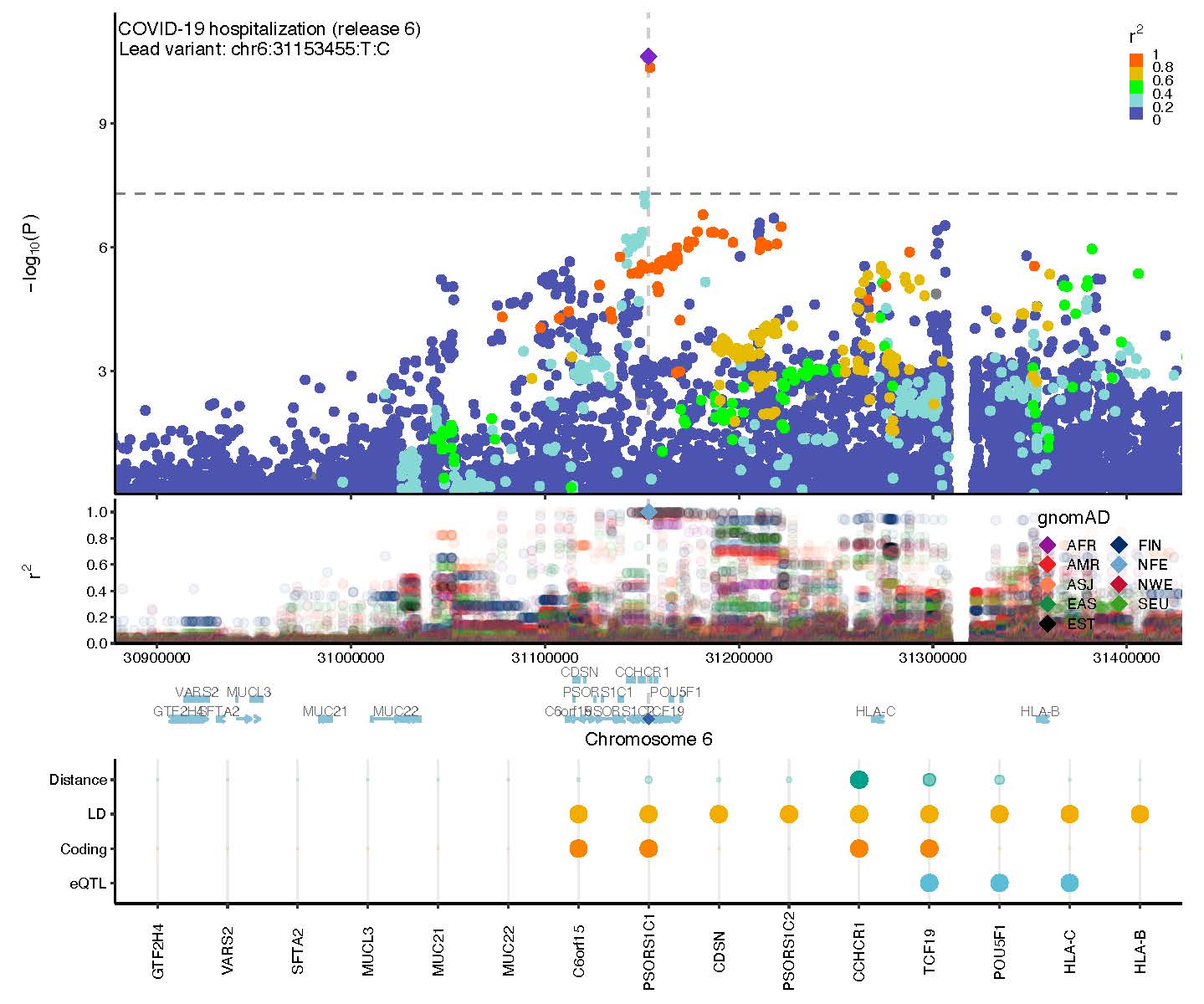

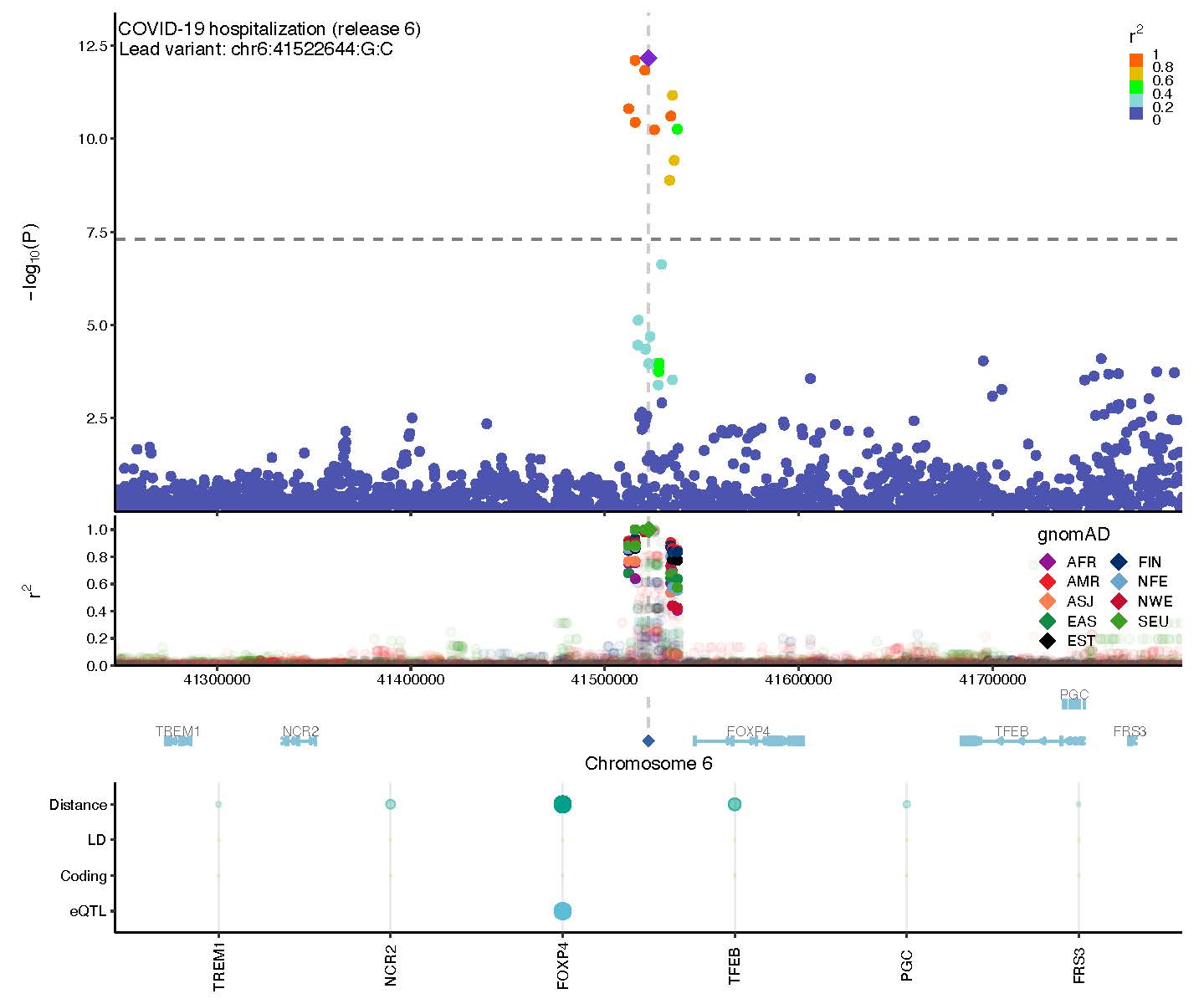

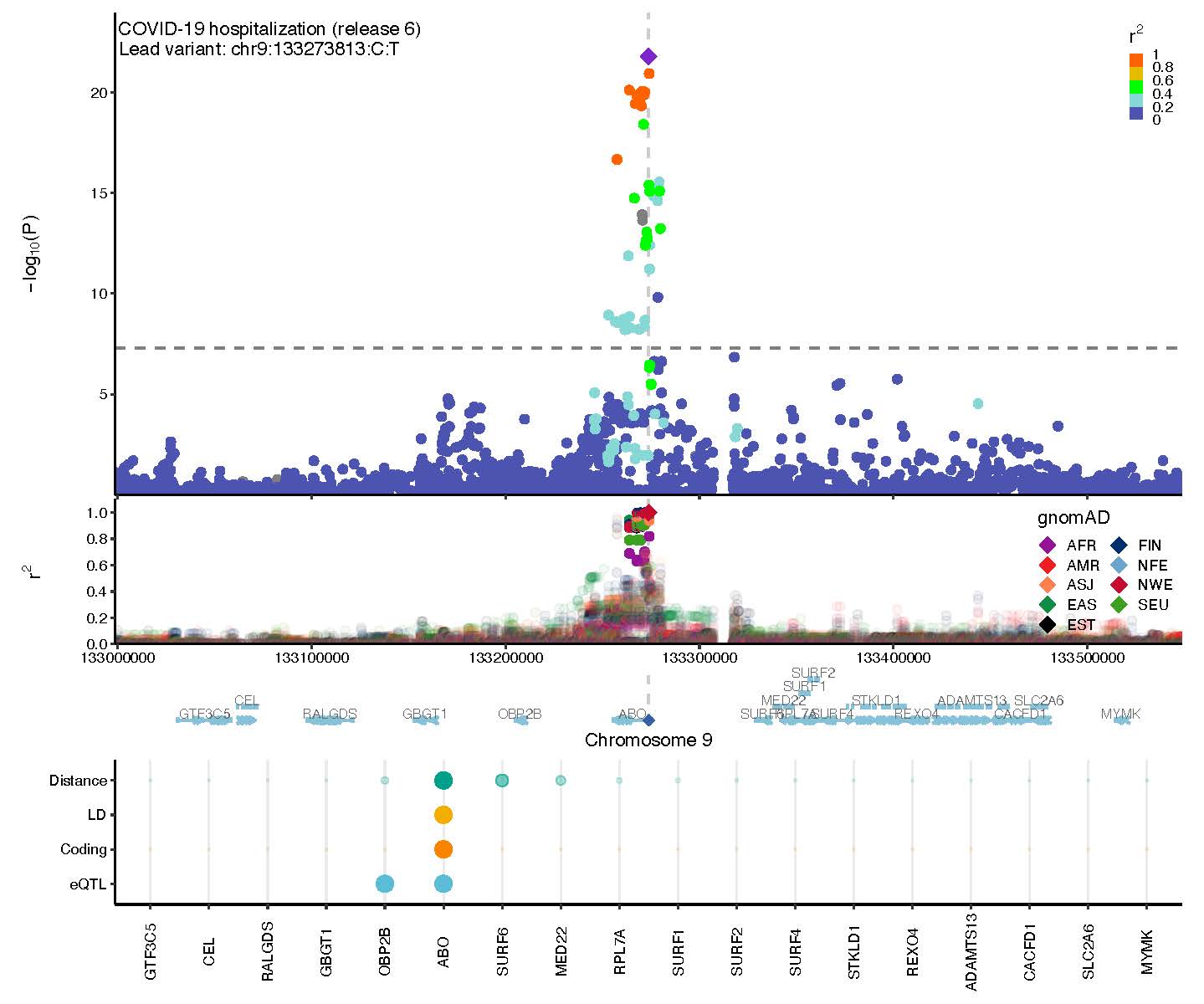

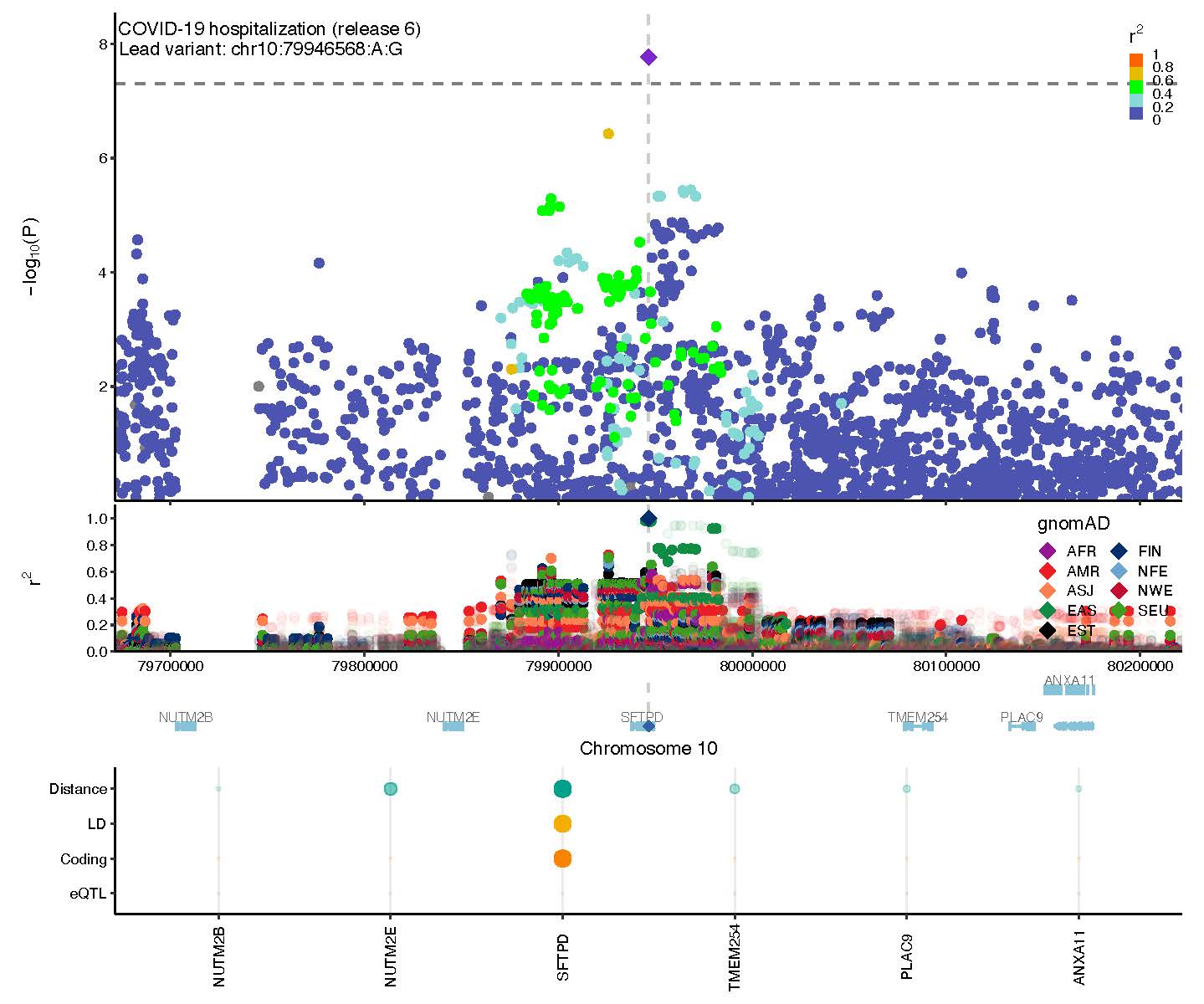

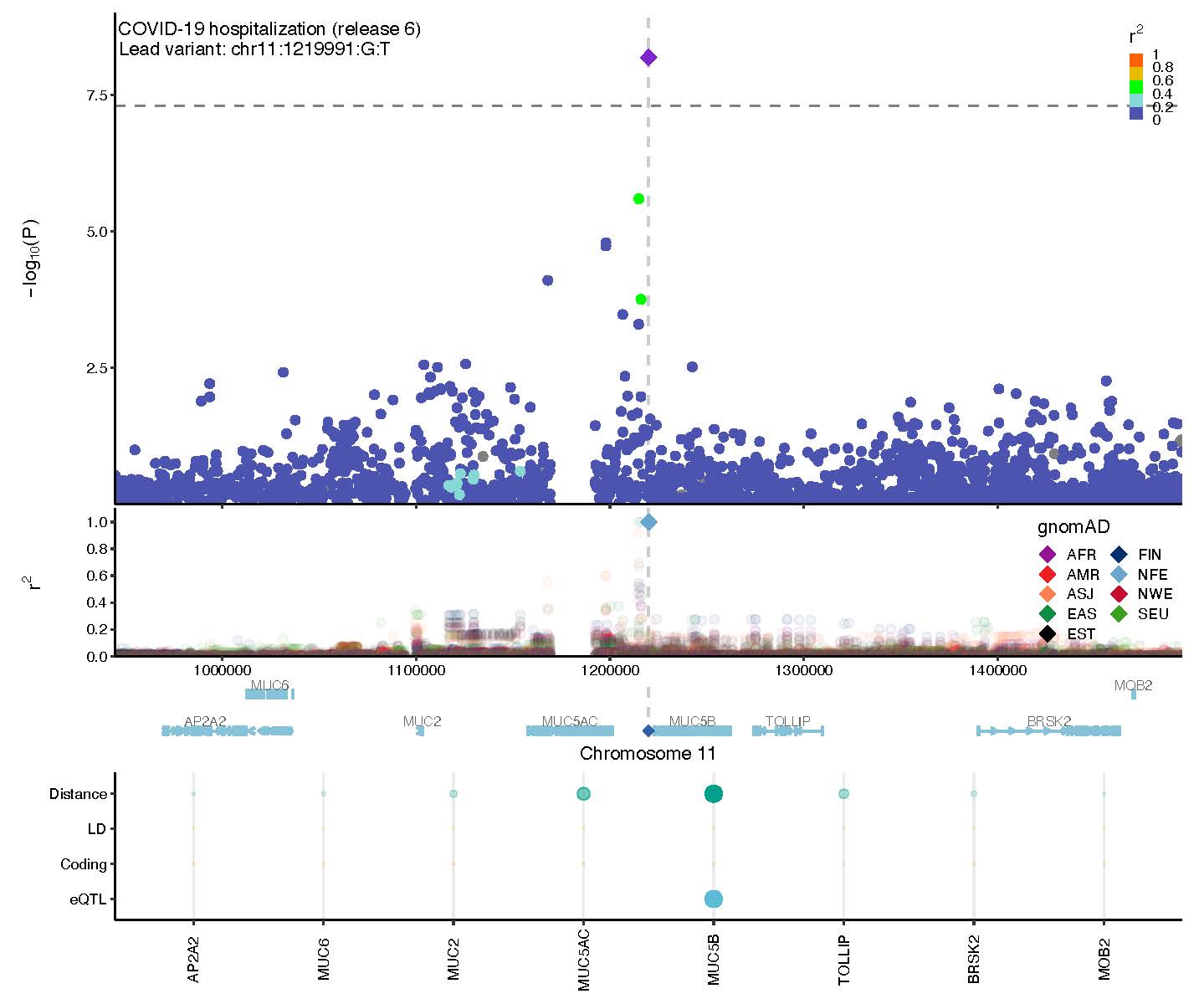

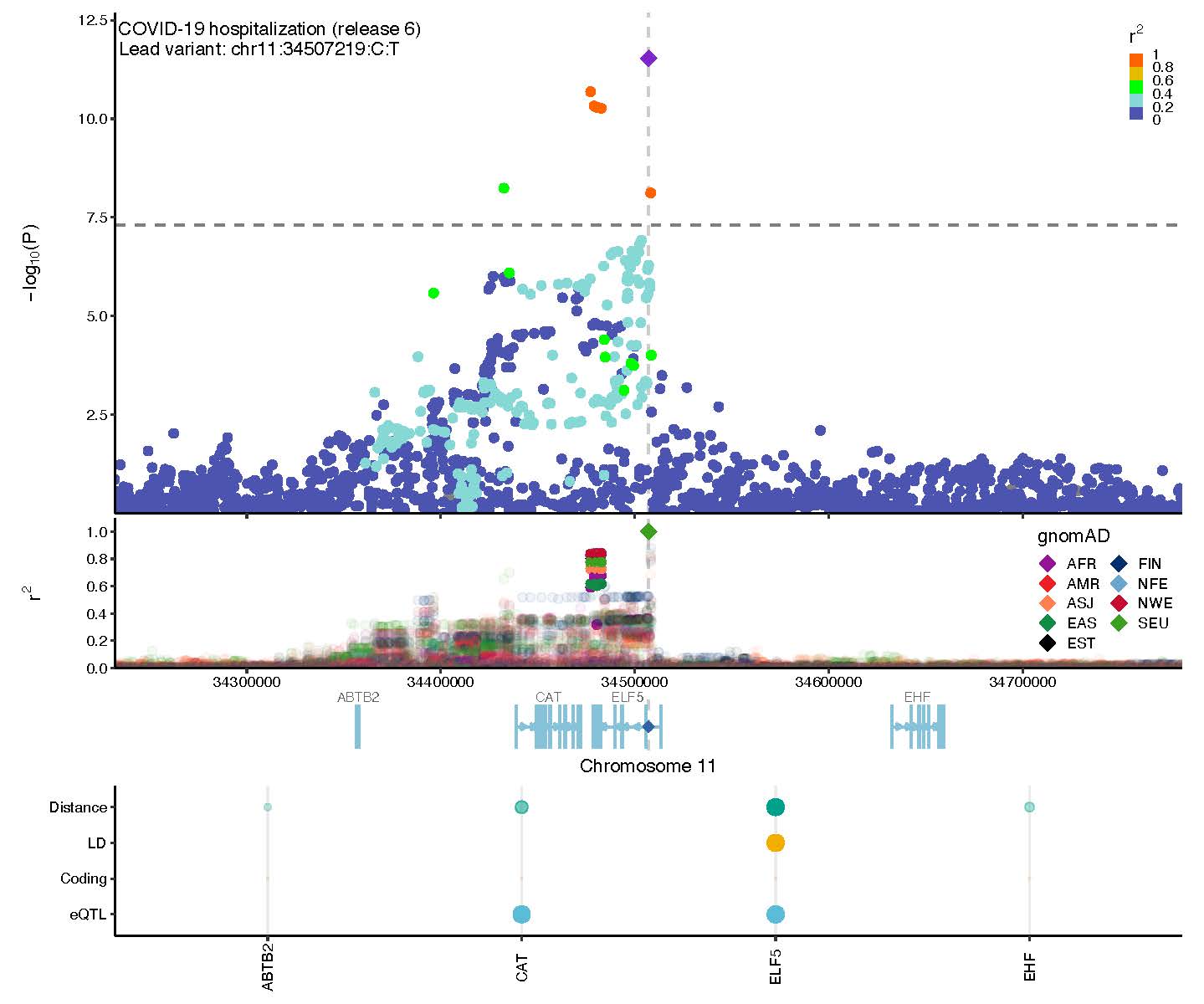

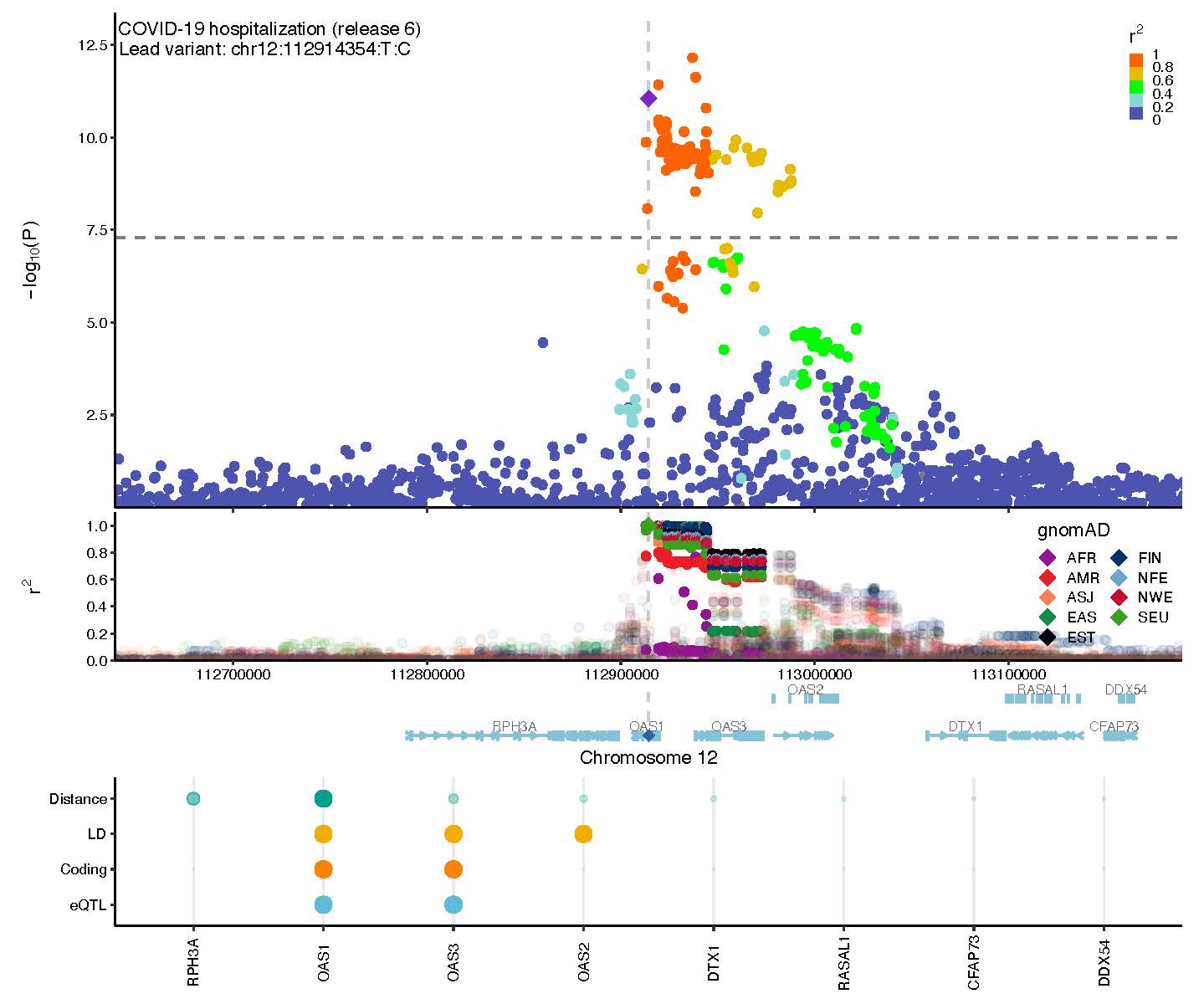

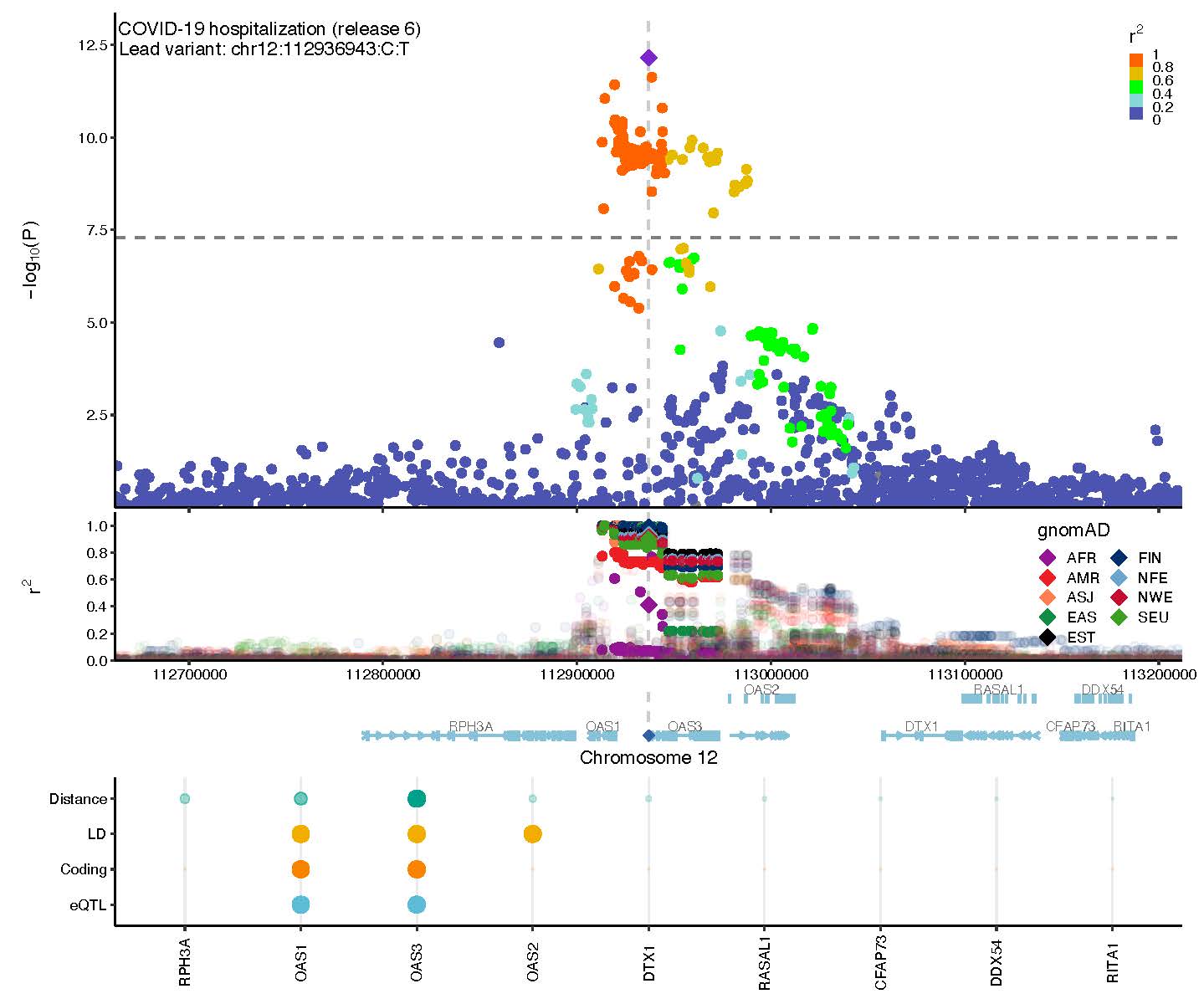

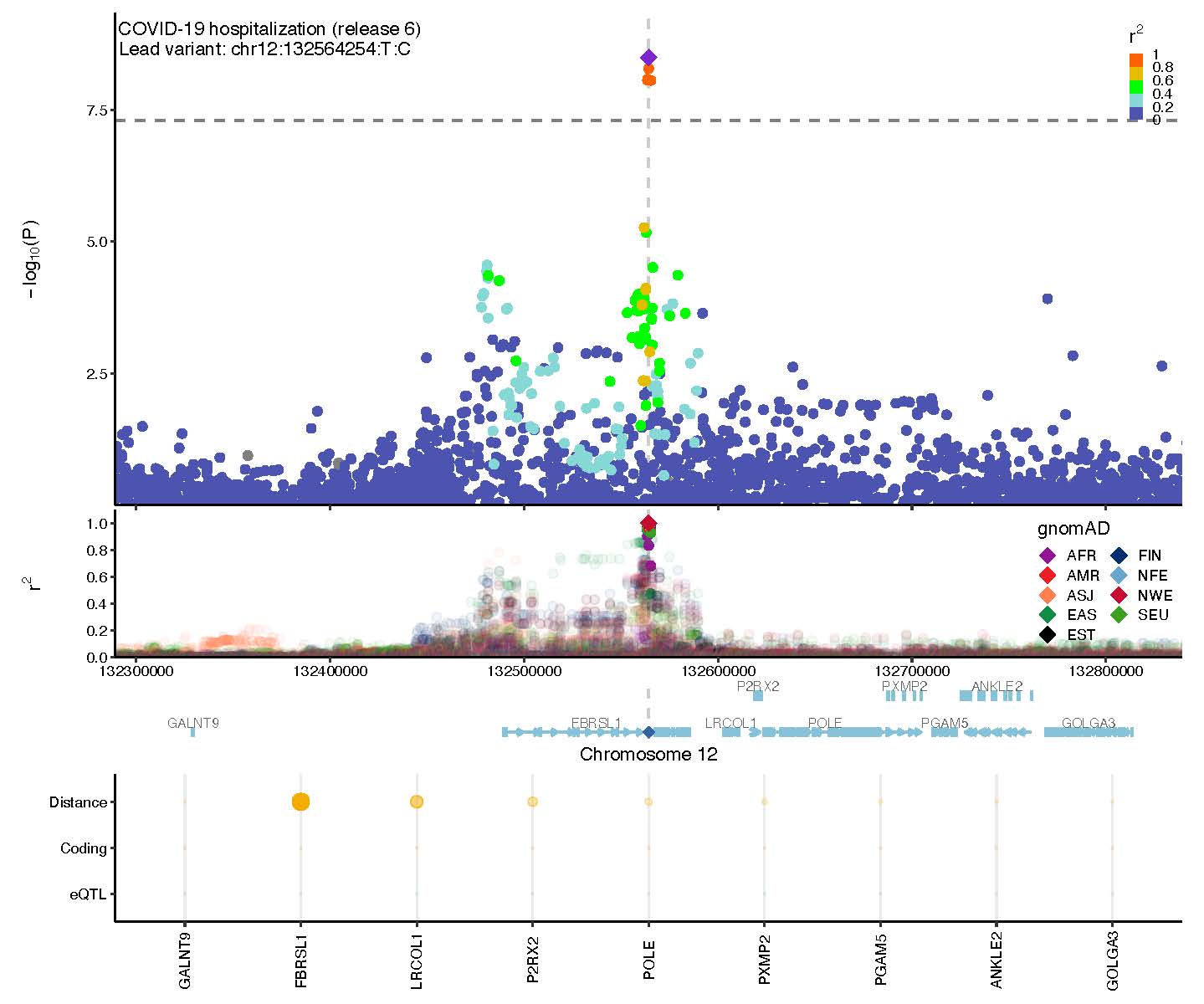

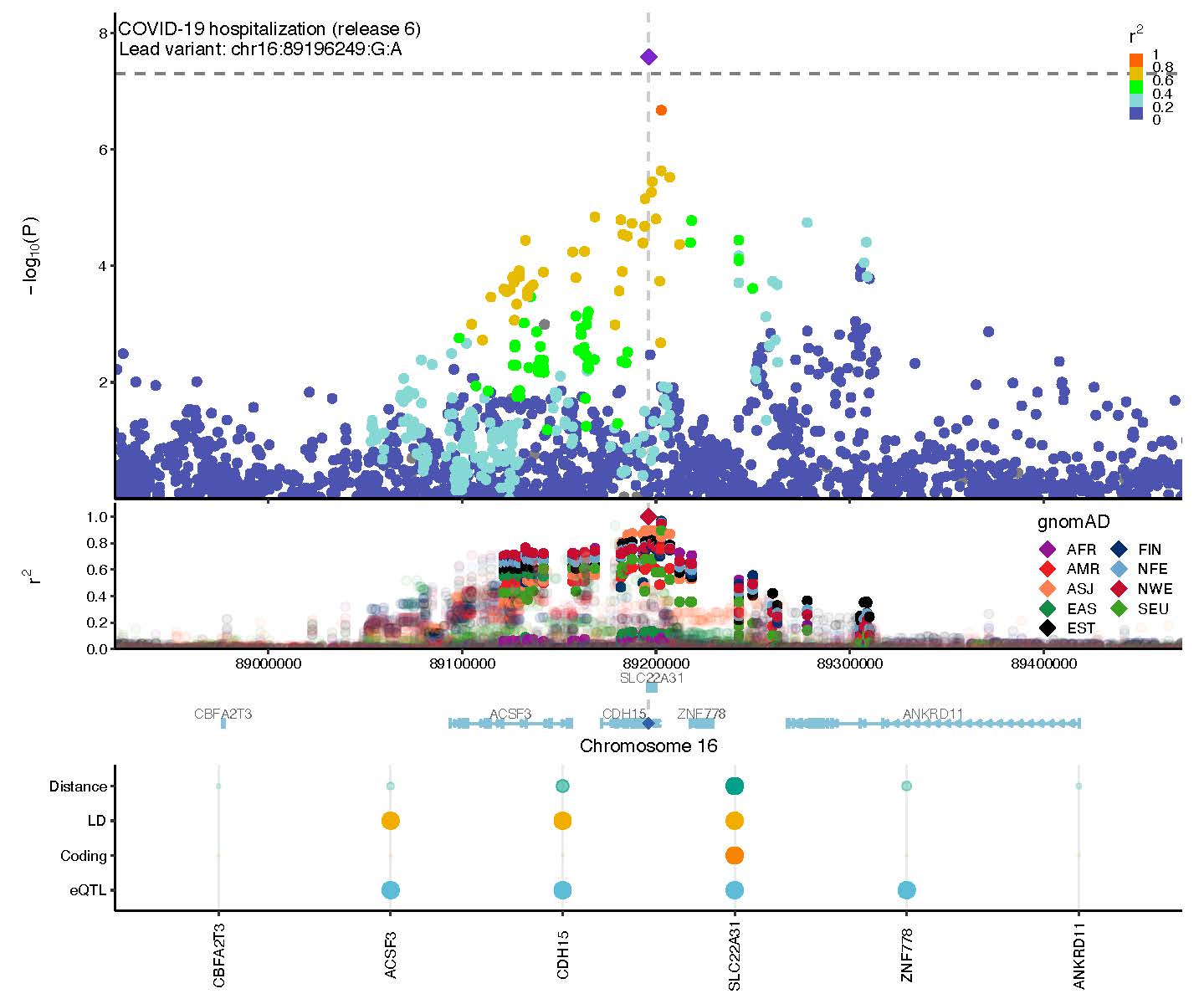

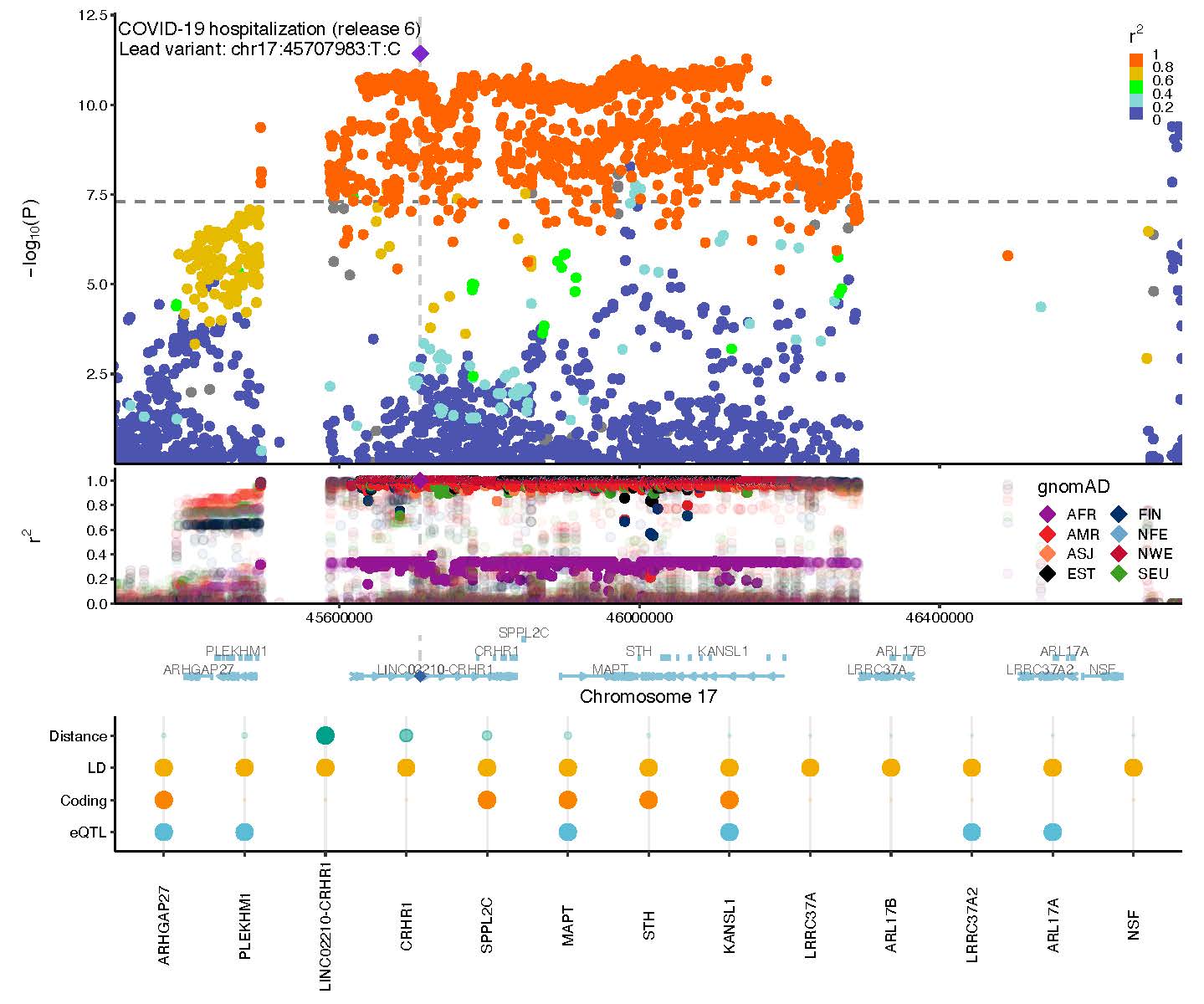

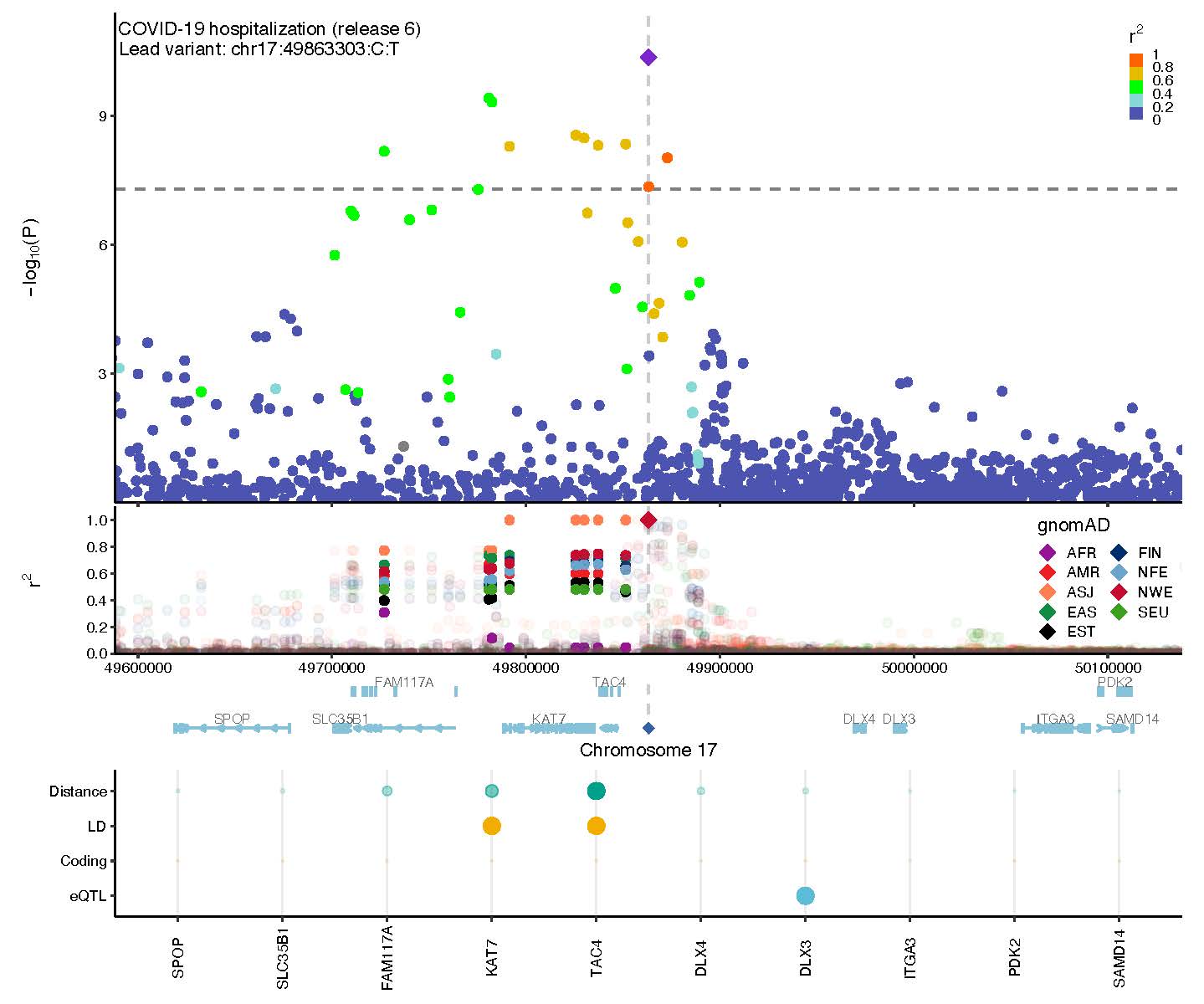

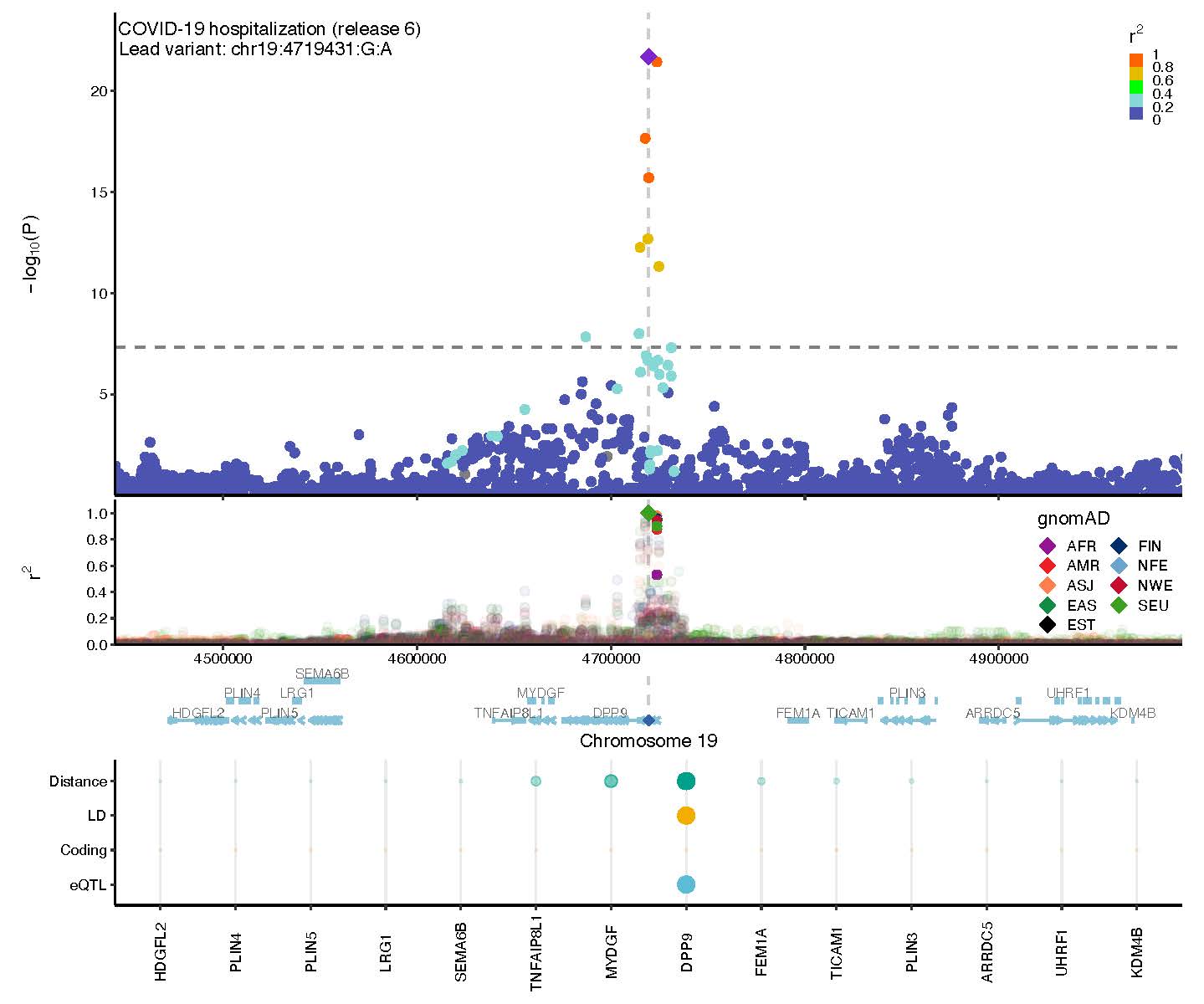

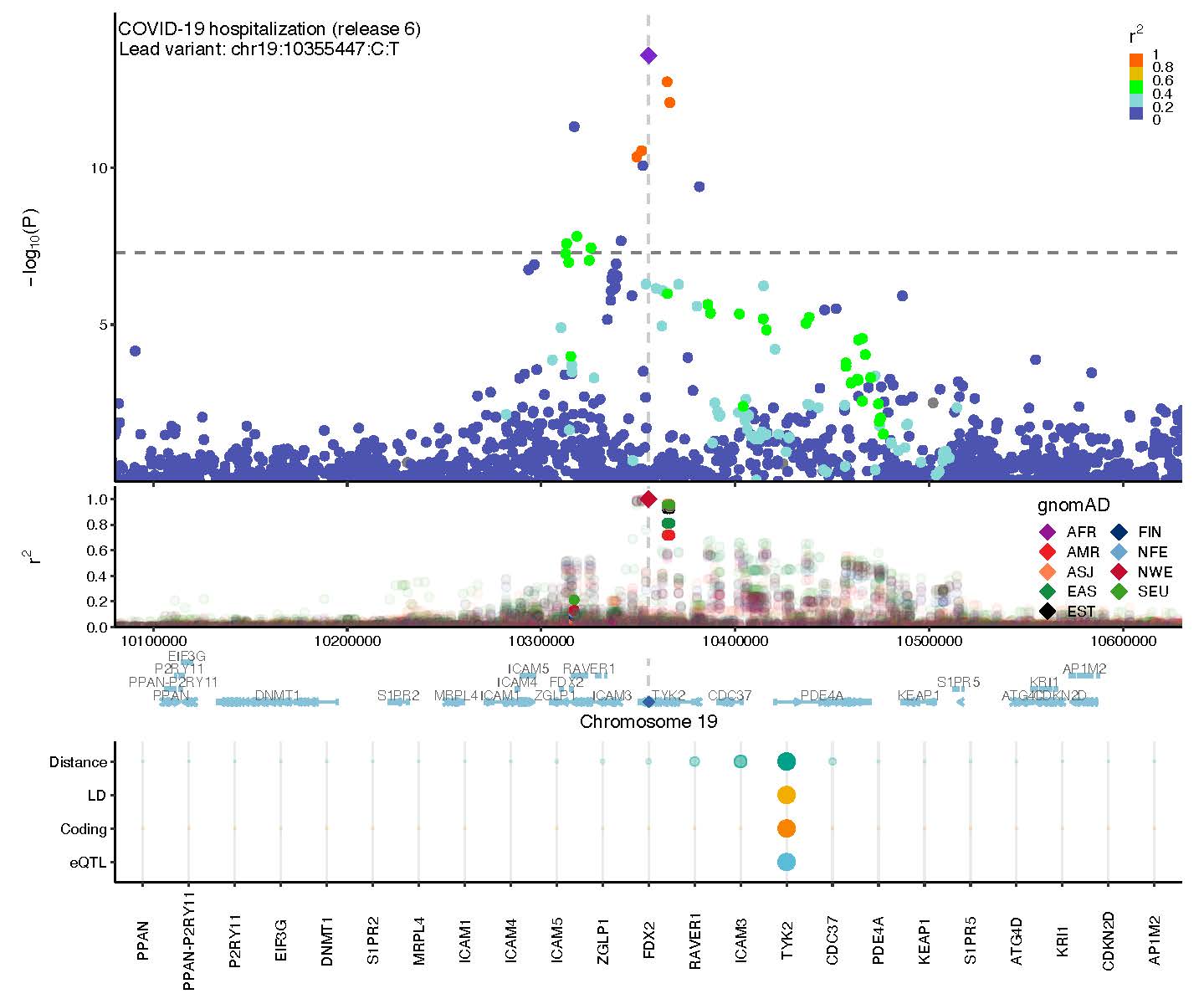

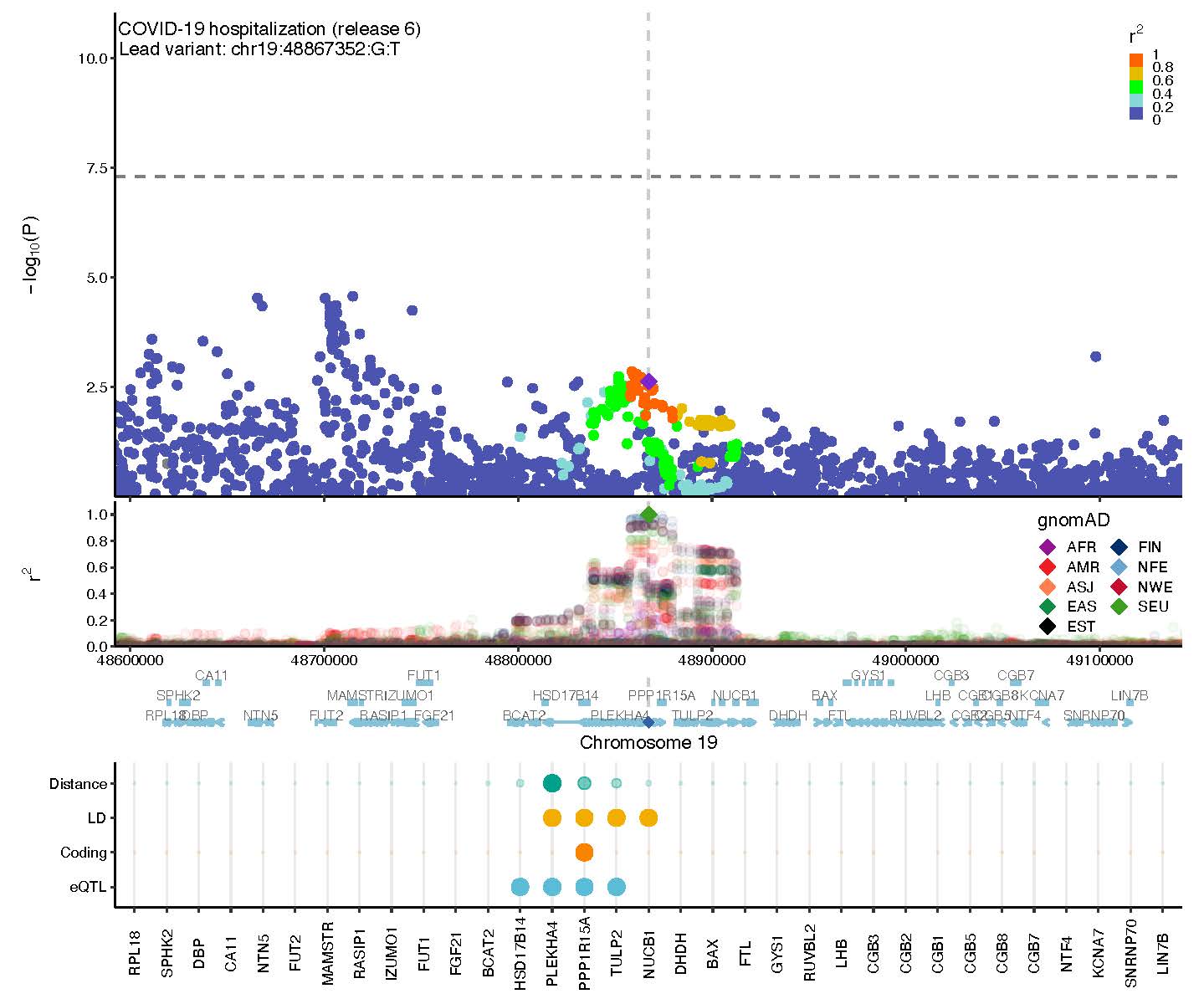

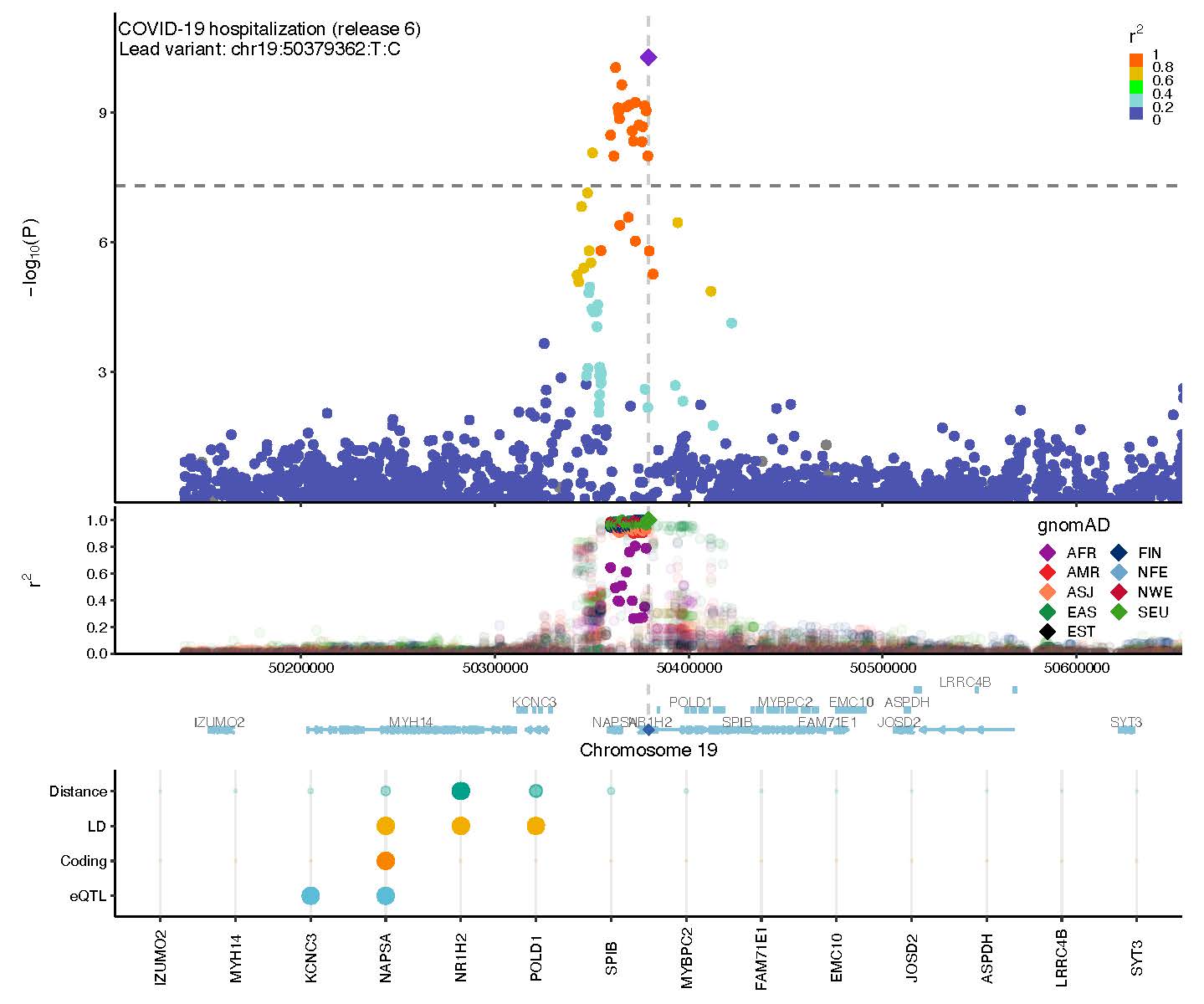

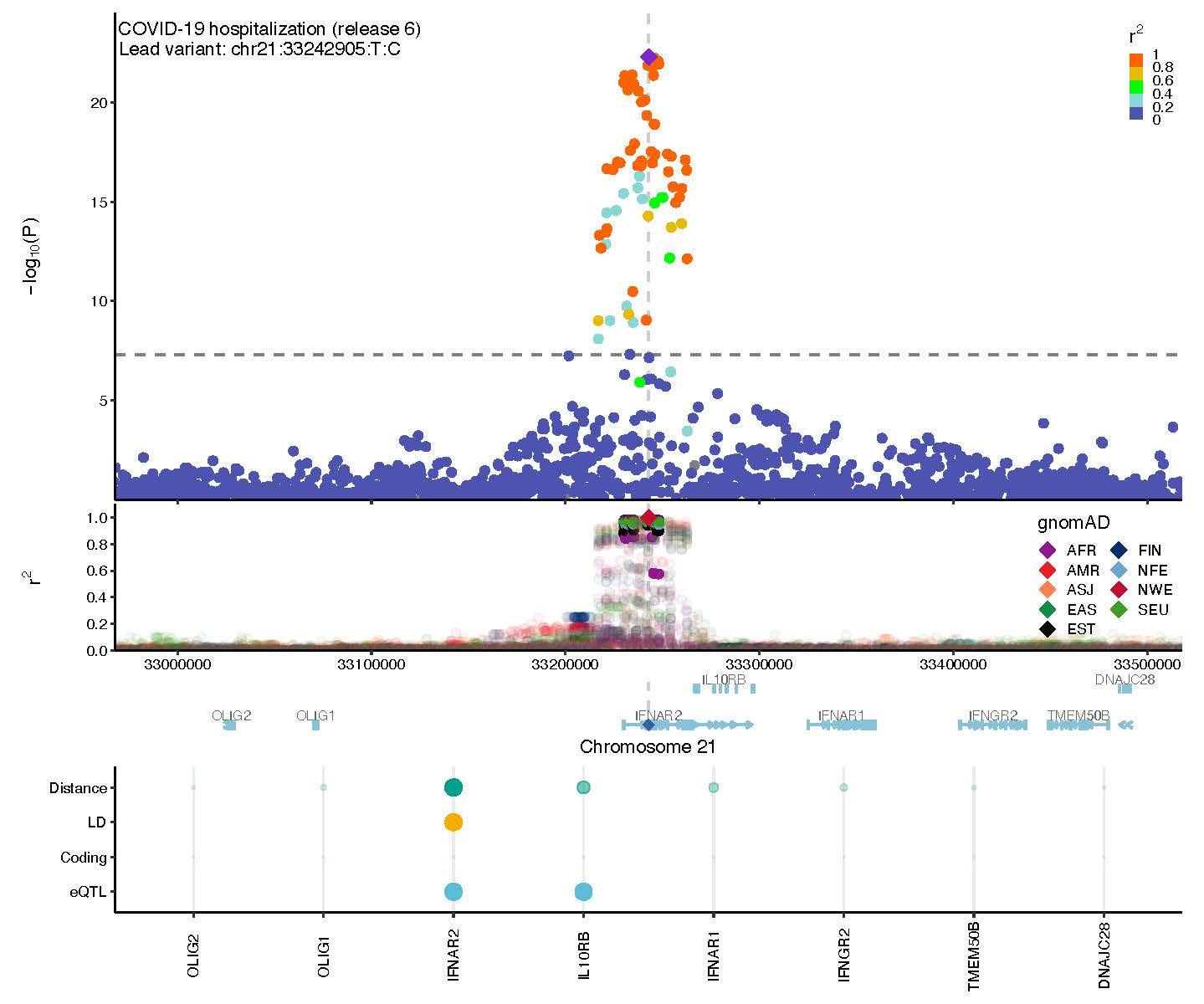

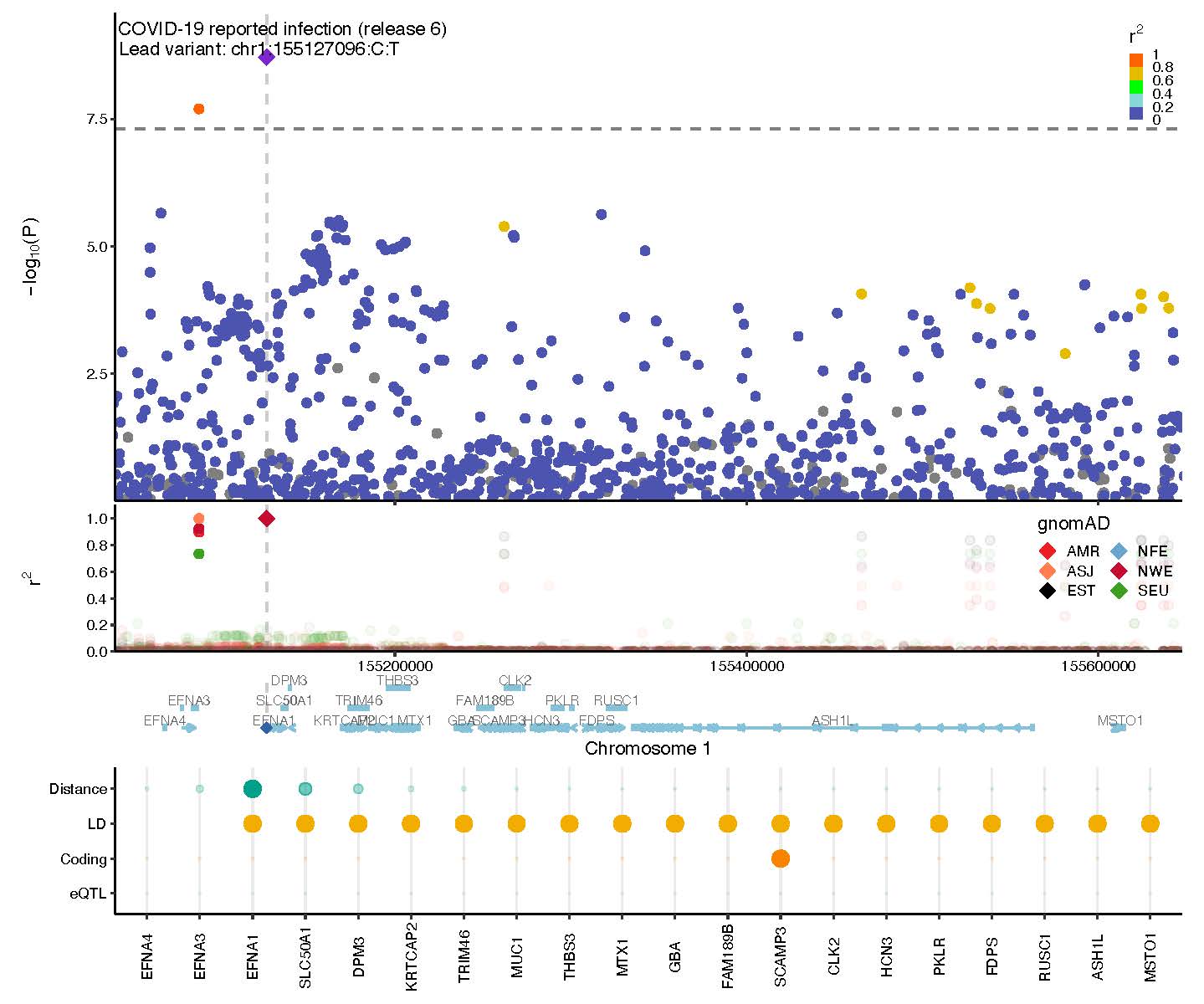

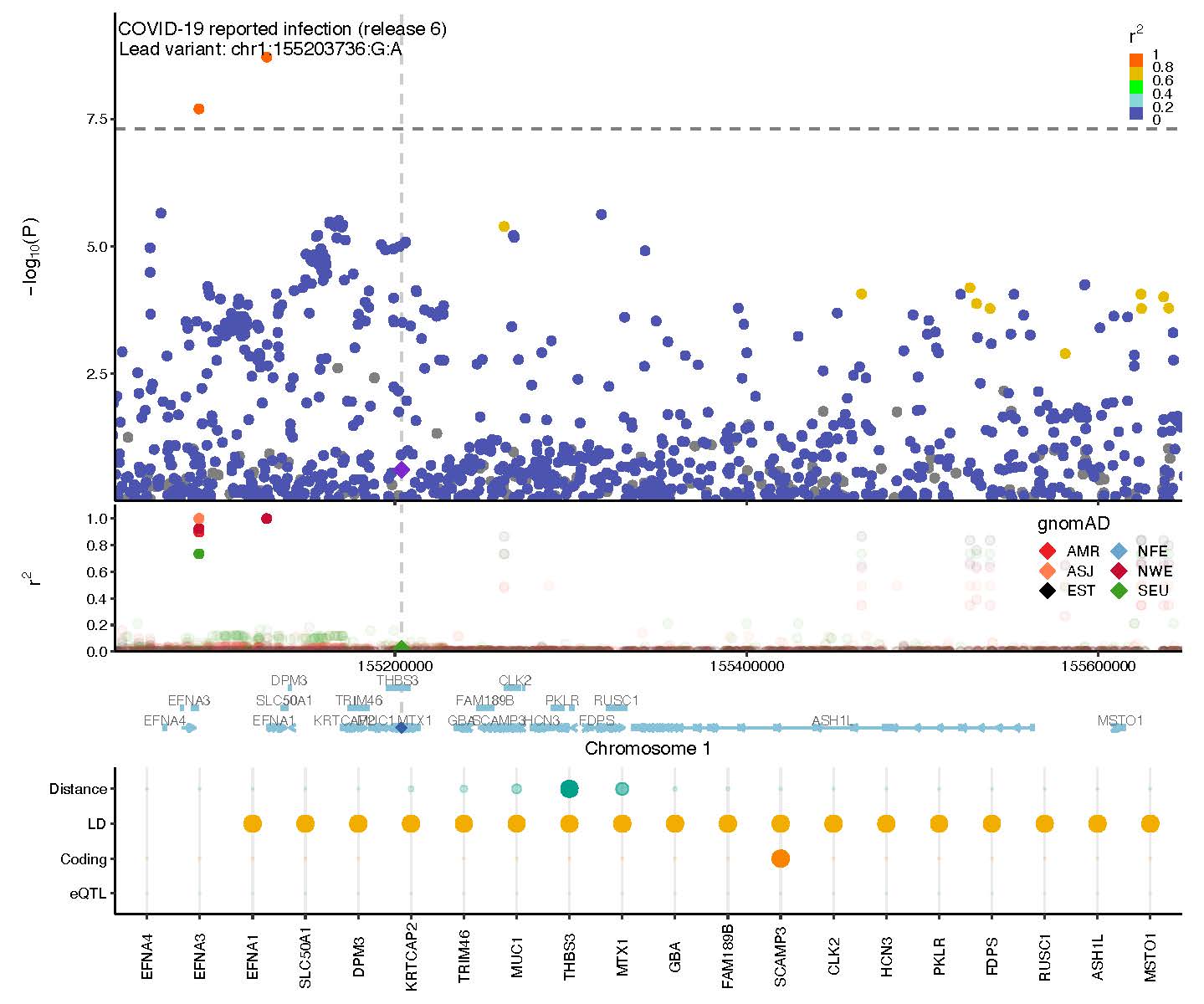

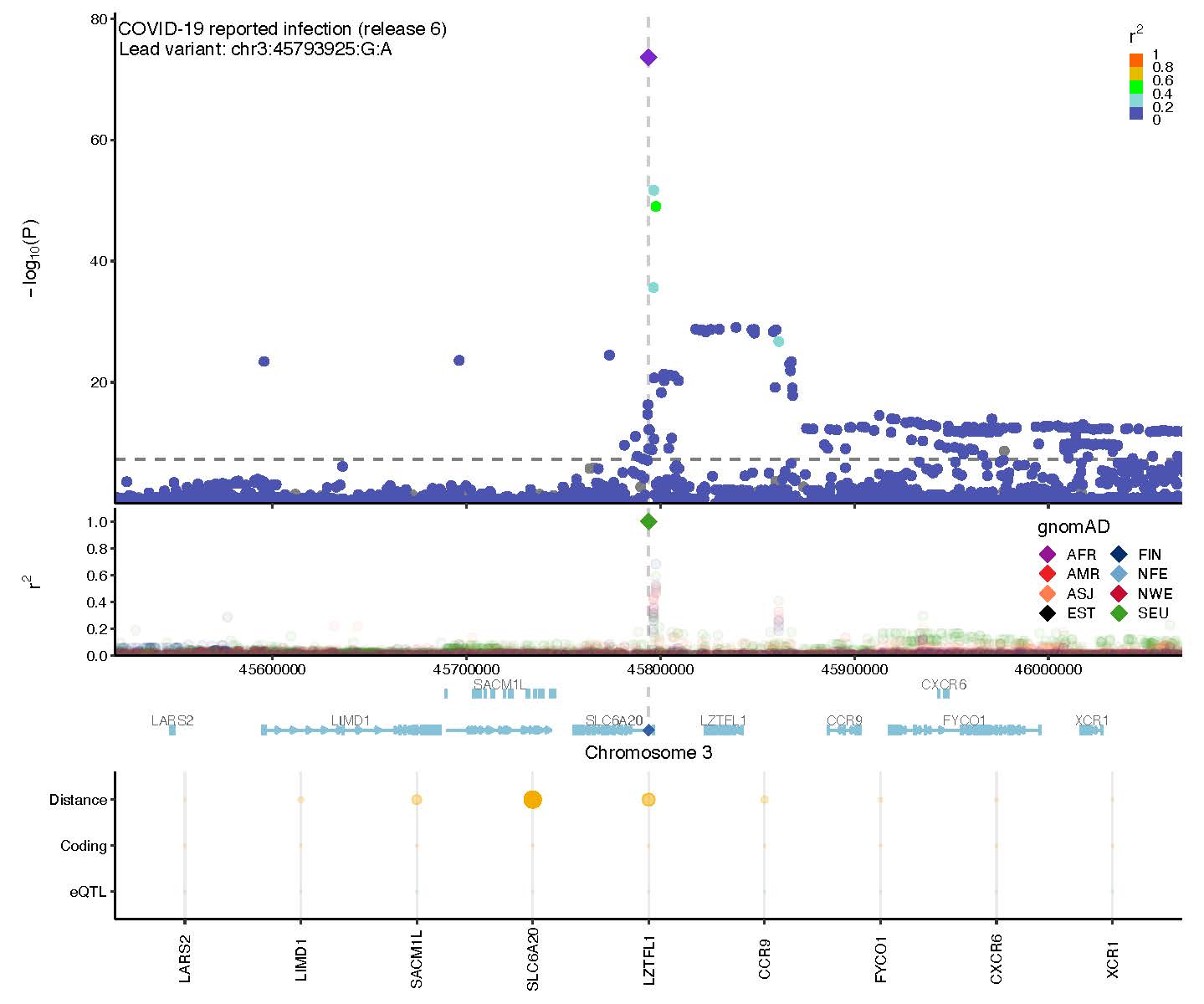

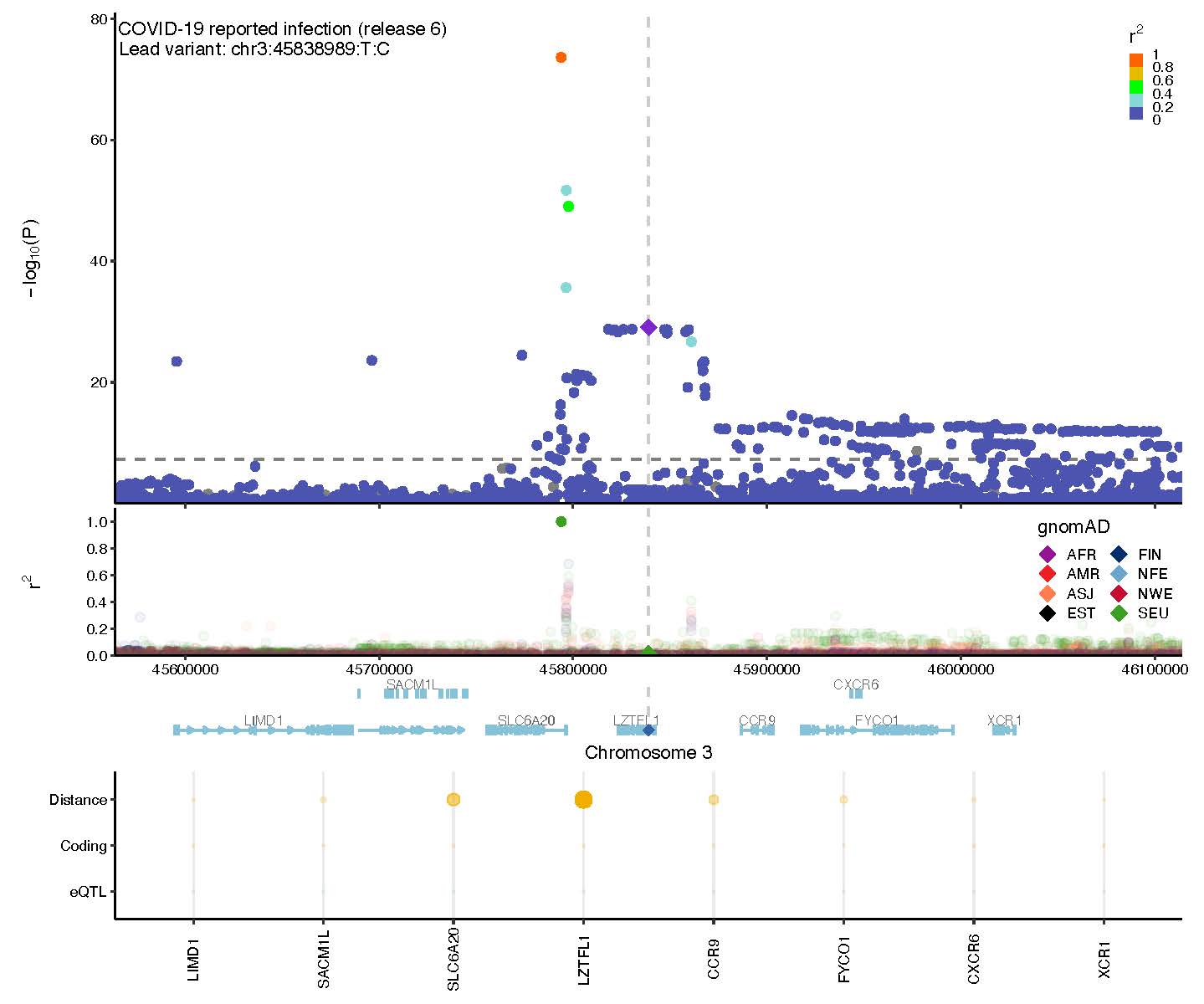

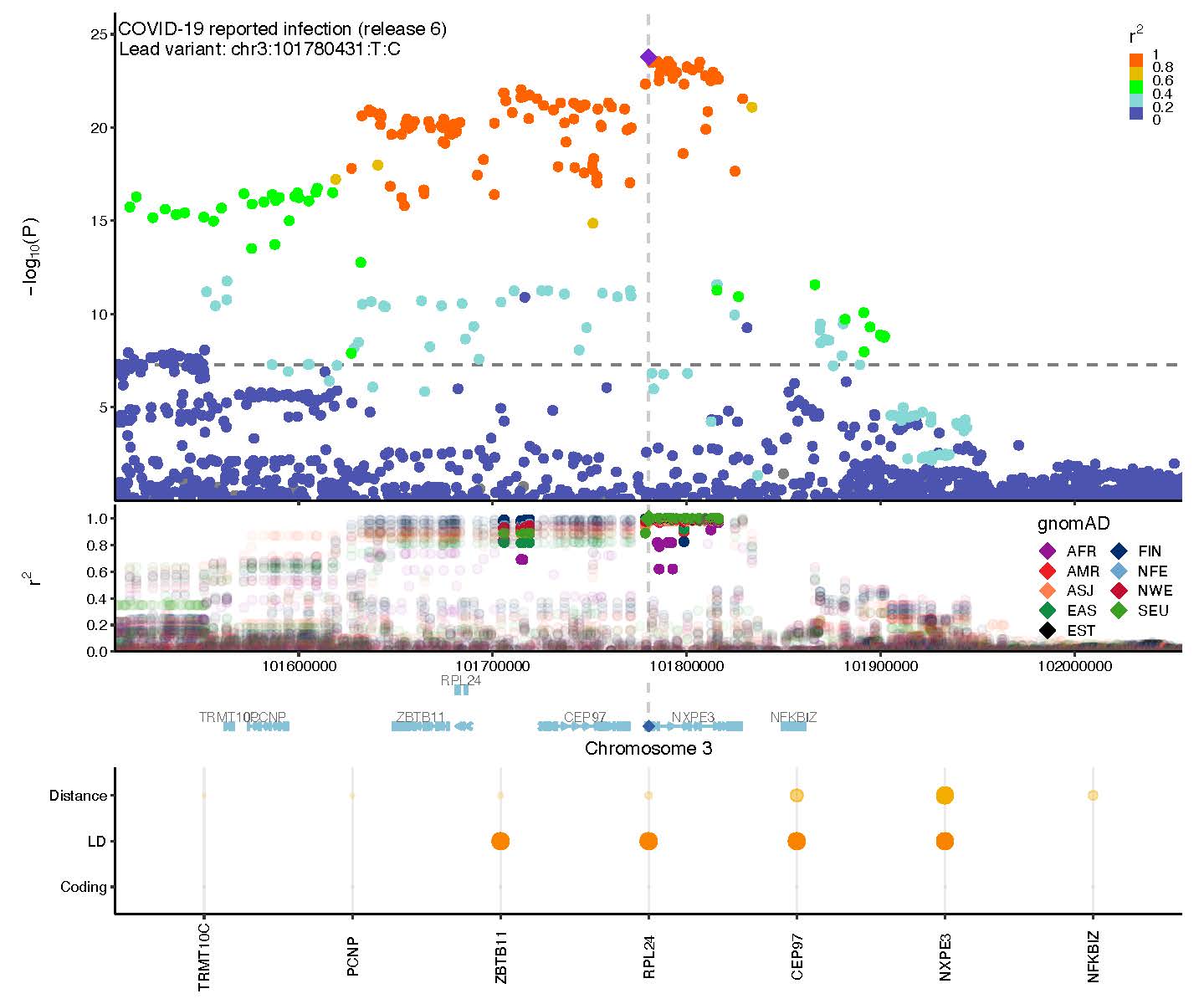

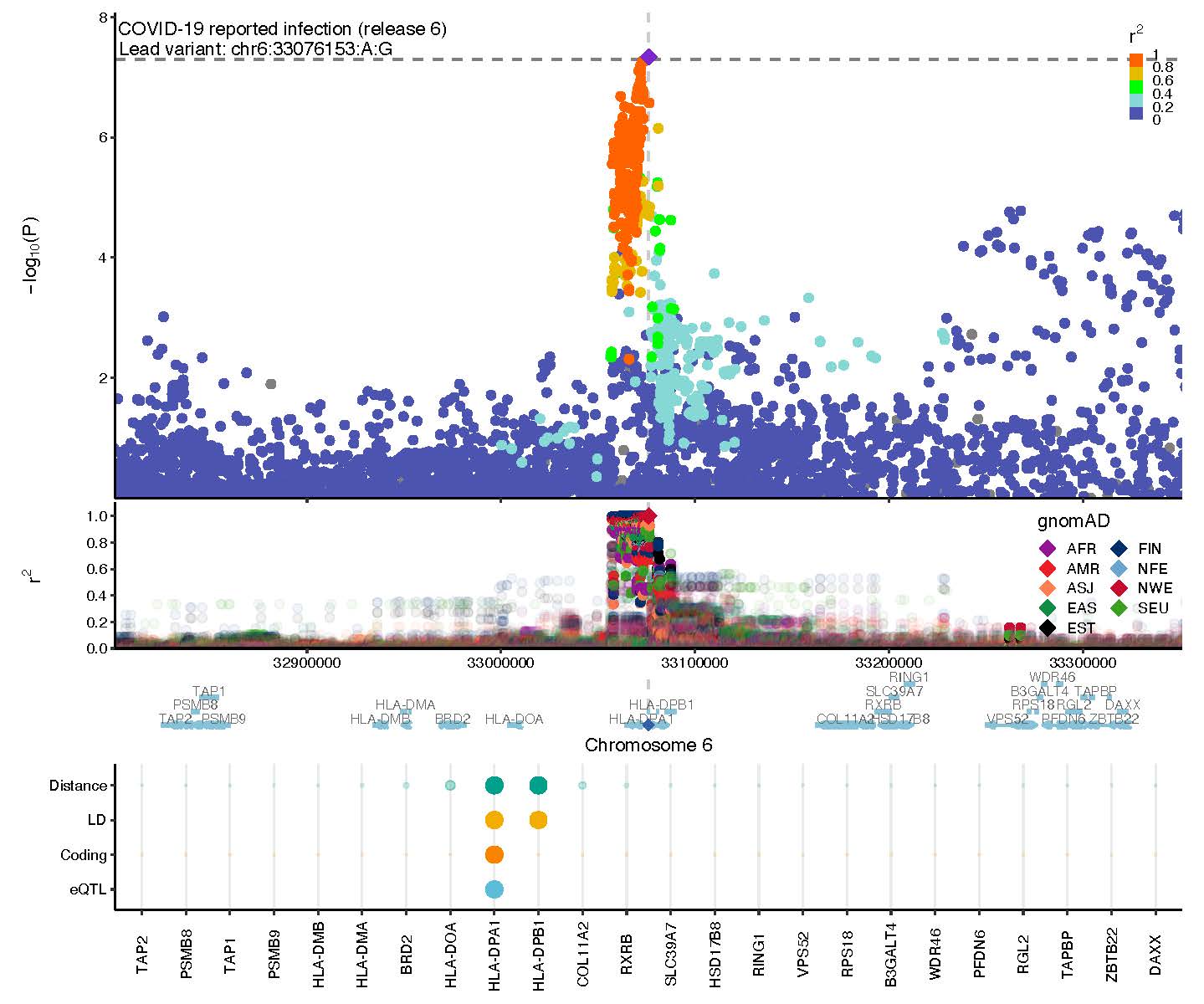

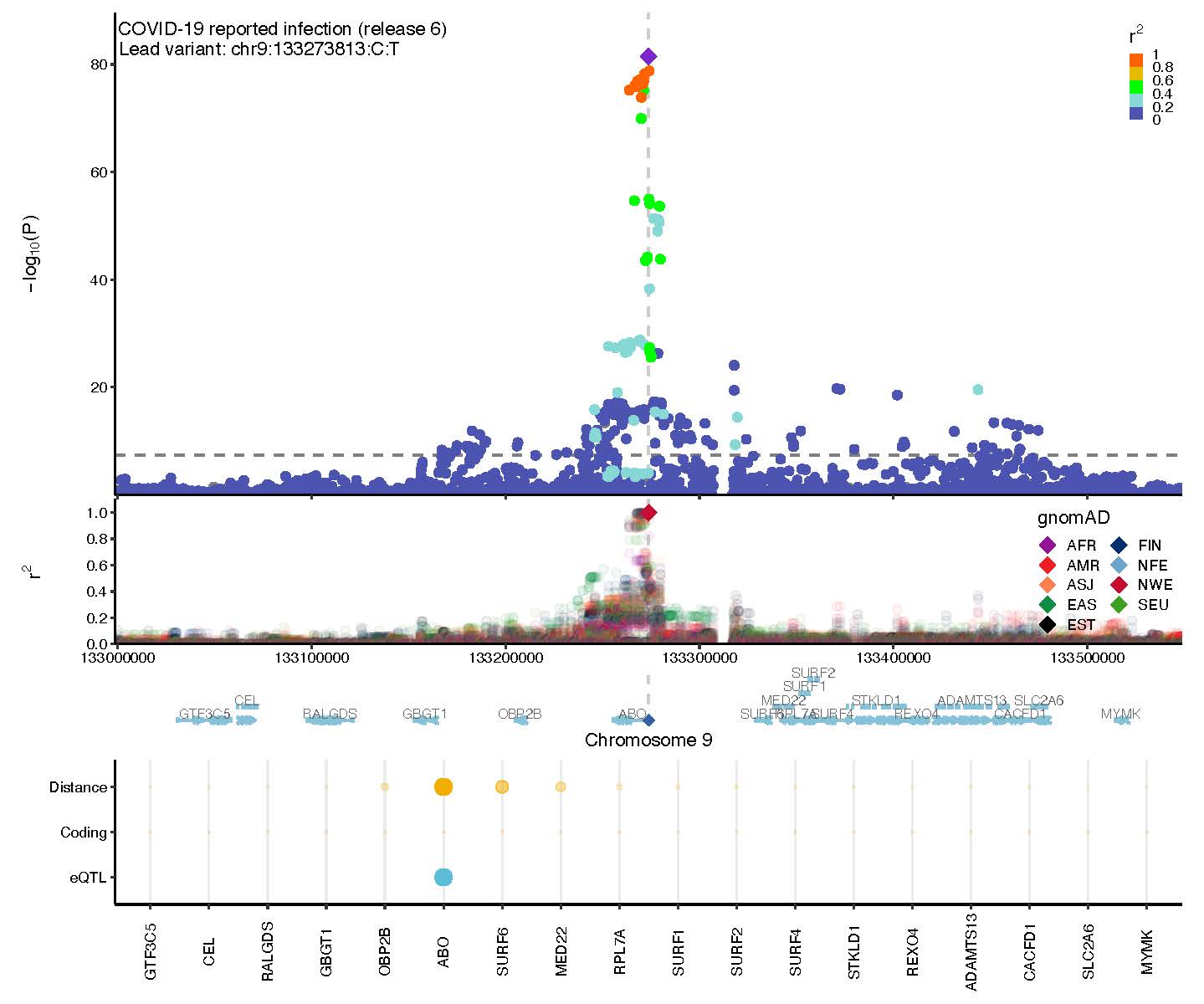

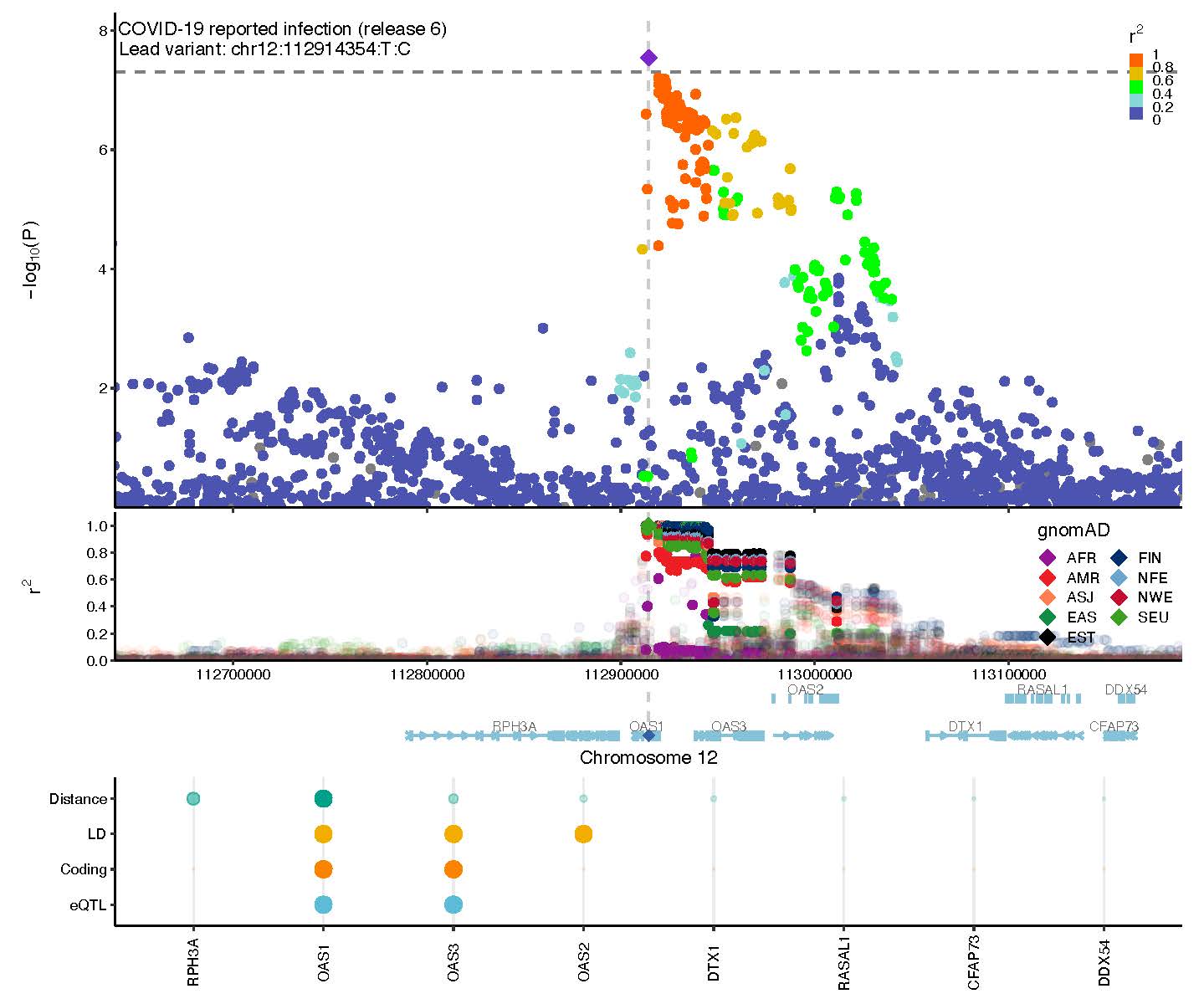

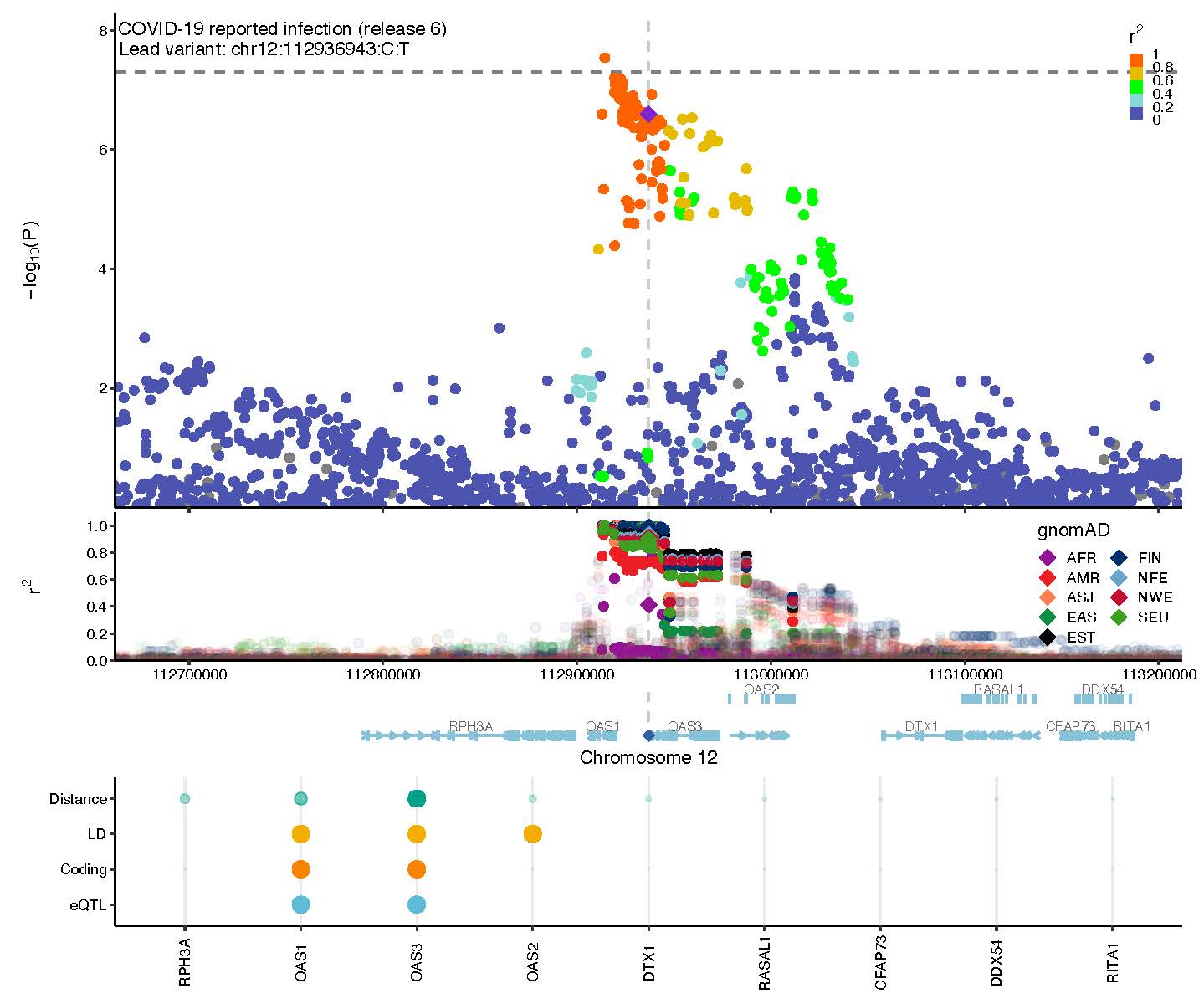

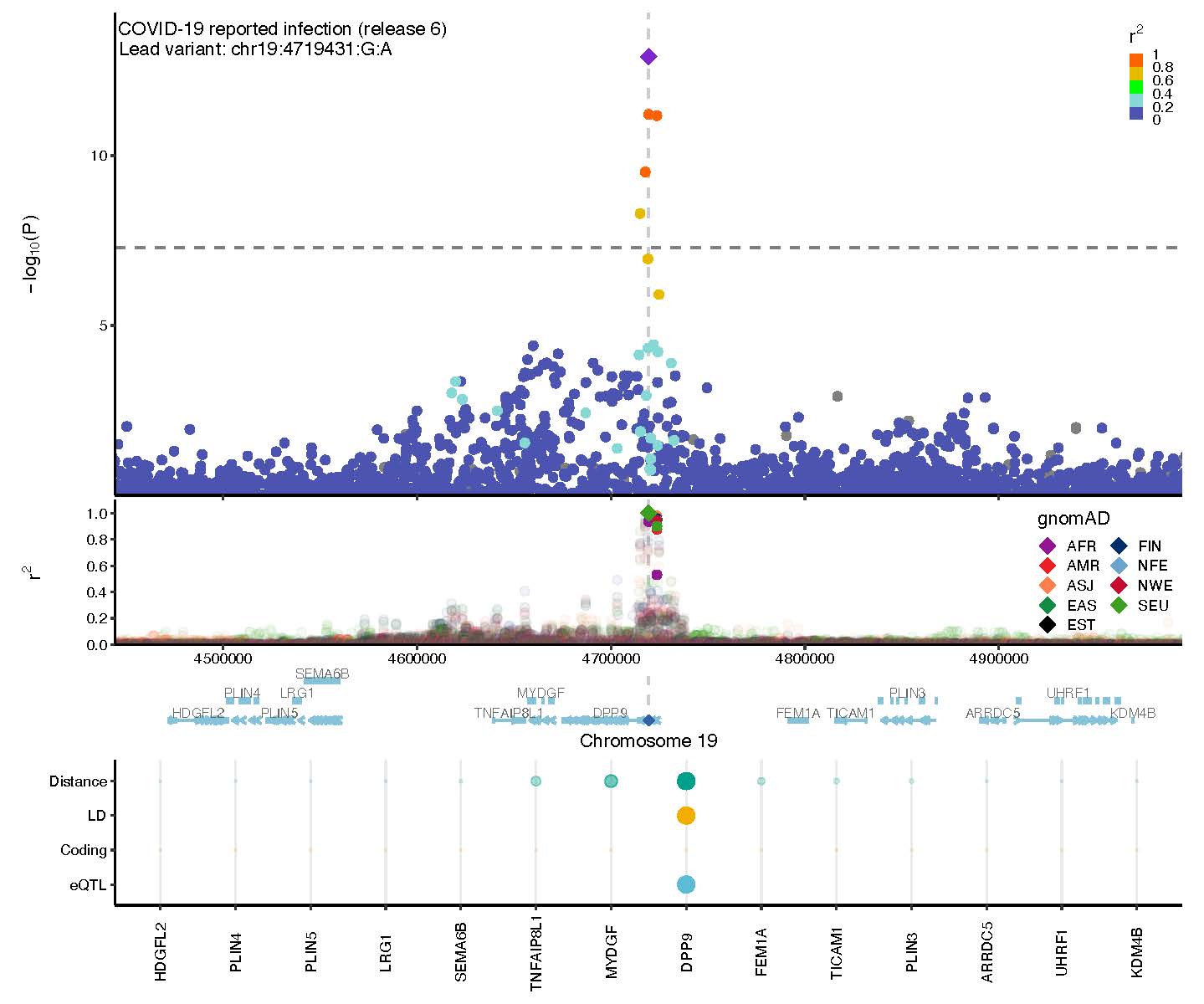

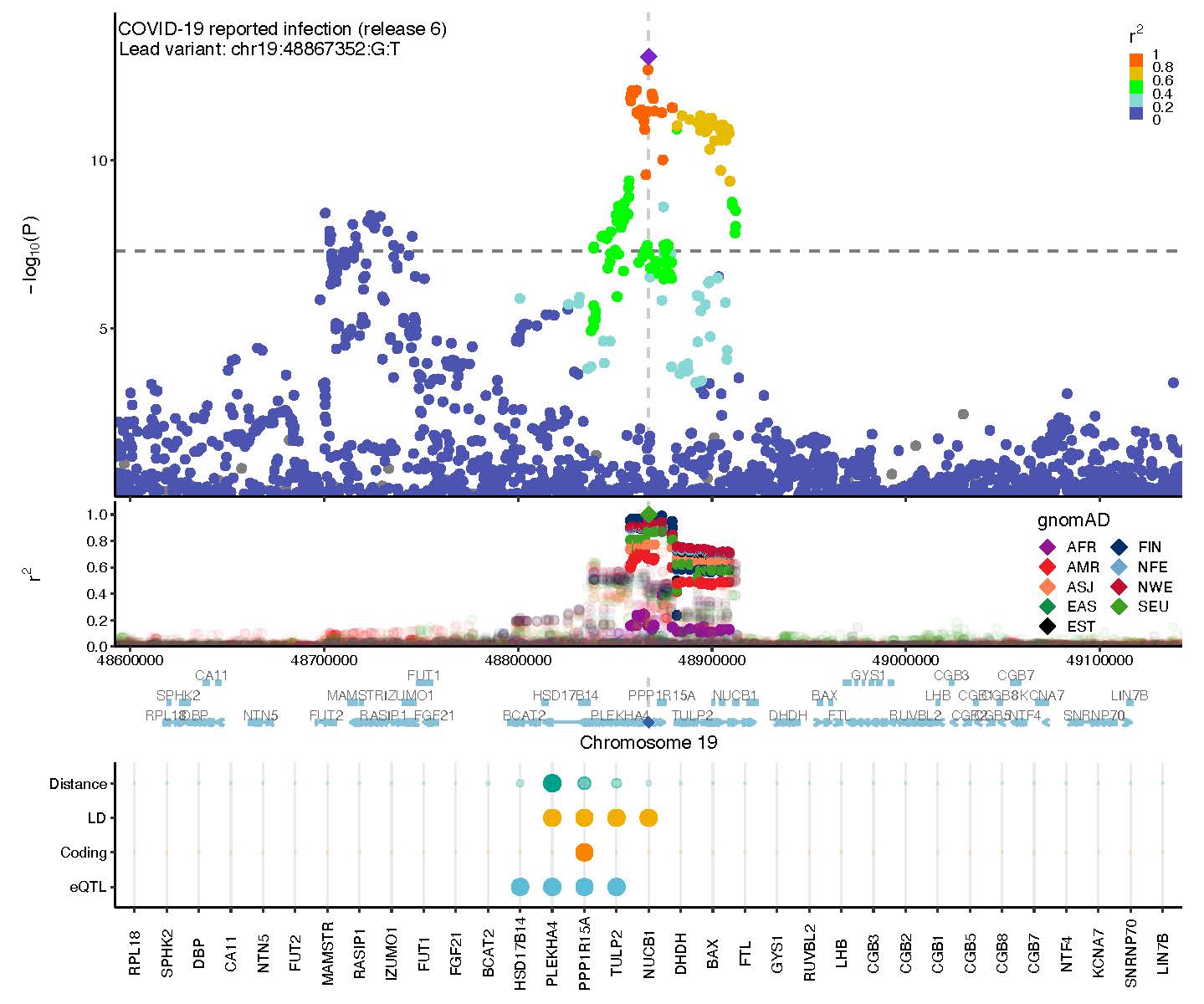

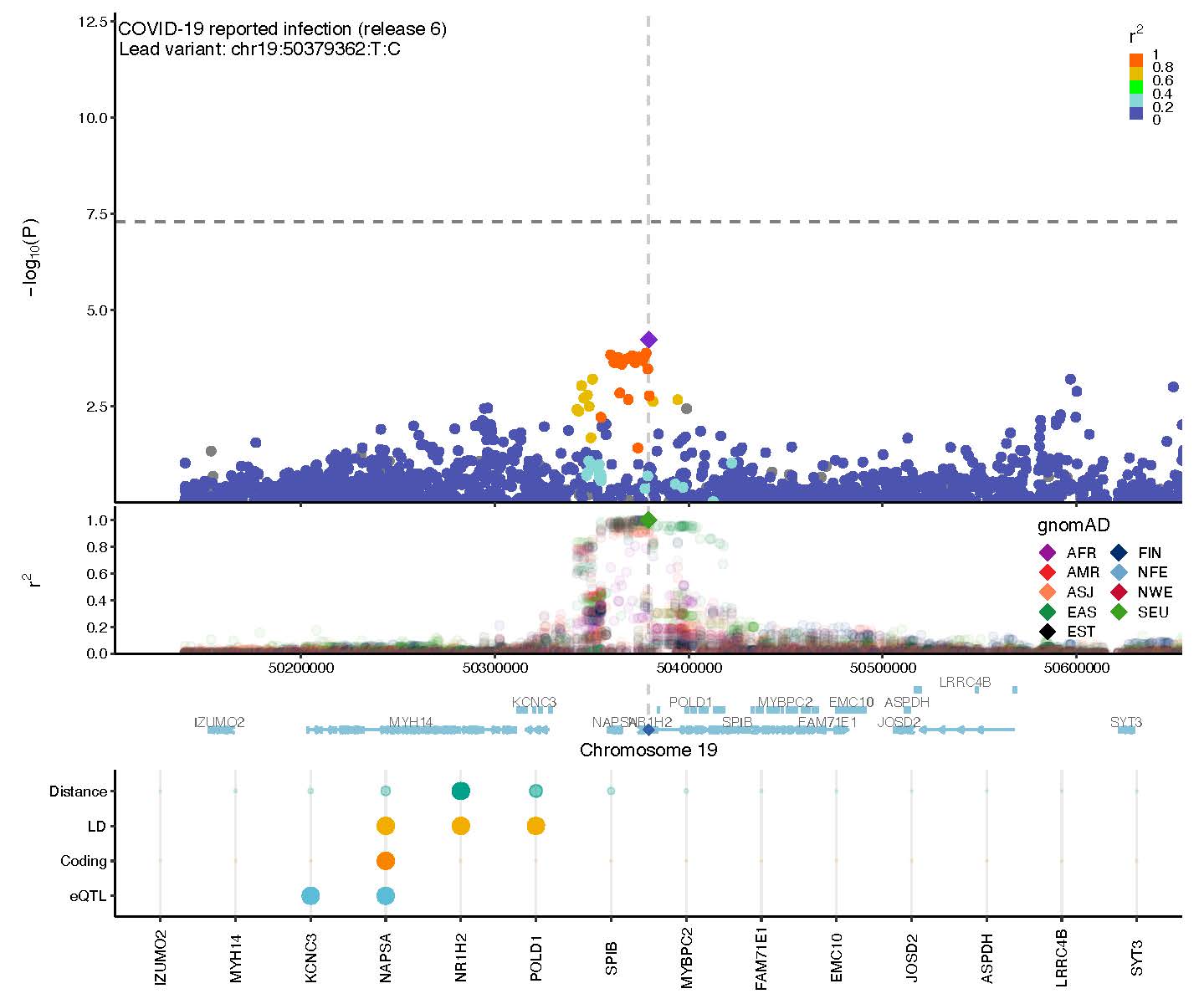

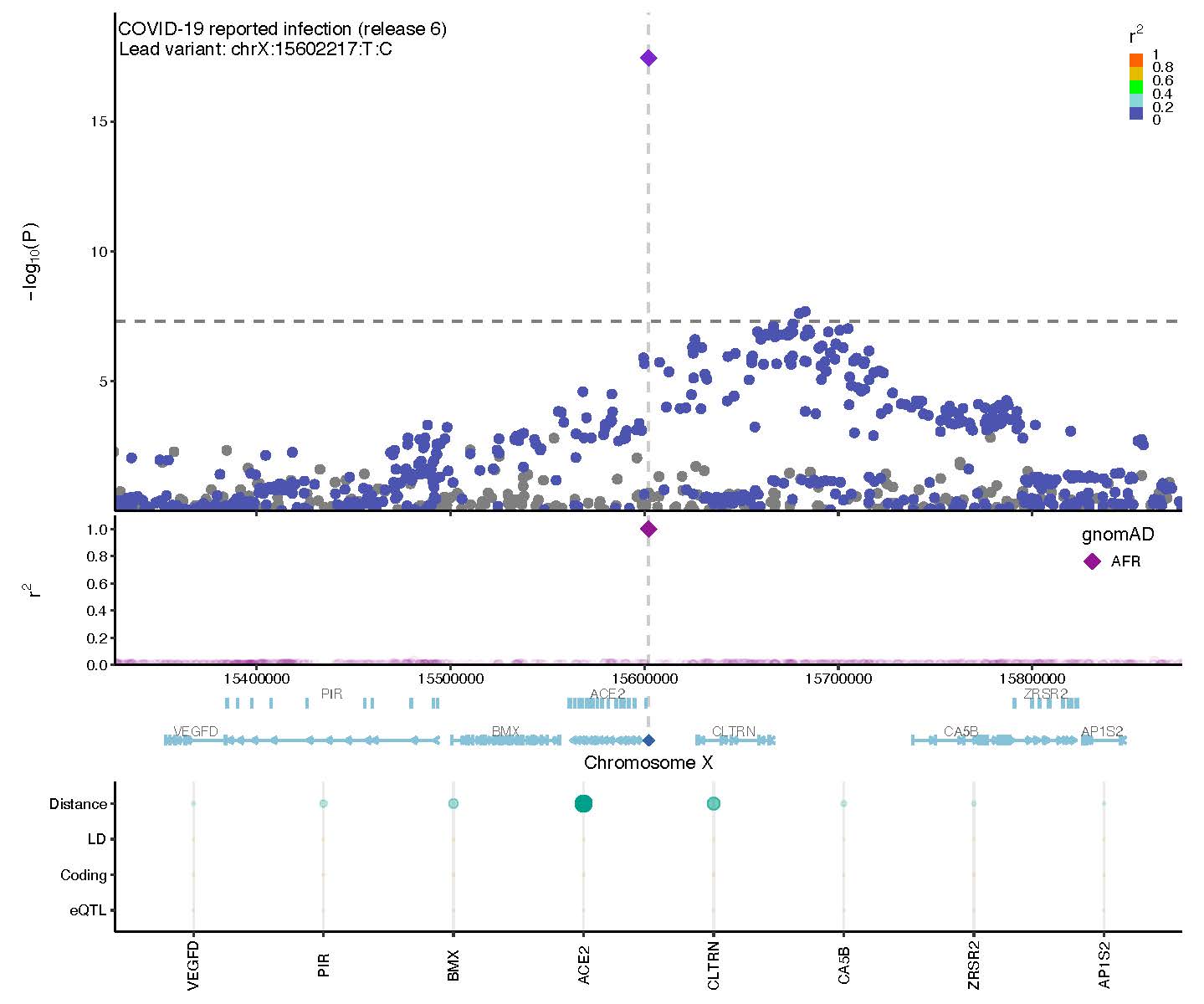

**Supplementary Figure 8**

**Genetic correlations and Mendelian randomization causal estimates between 38 traits and COVID-19 critical illness, hospitalization, and SARS-CoV-2 reported infection.** Larger squares correspond to more significant P-values, with genetic correlations or MR causal estimates significantly different from zero at a P < 0.05 shown as a full-sized square. Genetic correlations or causal estimates that are significantly different from zero at a false discovery rate (FDR) of 5% are marked with an asterisk. Two-sided *P*-values were calculated using LDSC for genetic correlations and Inverse variance weighted analysis for MR.

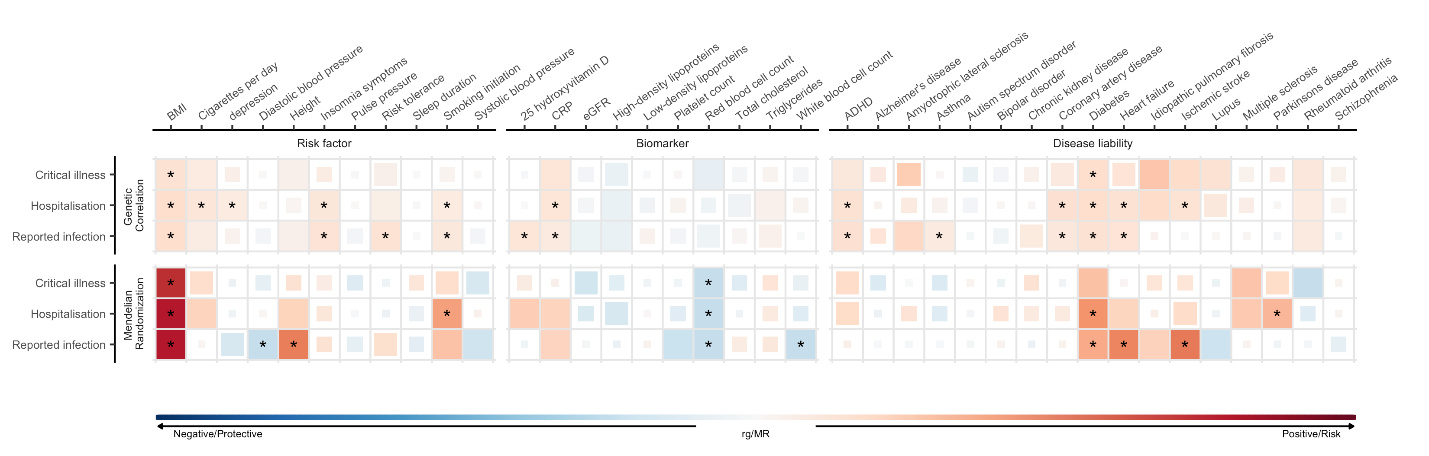
